# HECTOR: multimodal deep learning predicts recurrence risk in endometrial cancer

**DOI:** 10.1101/2023.11.27.23298994

**Authors:** Sarah Fremond-Volinsky, Nanda Horeweg, Sonali Andani, Jurriaan Barkey Wolf, Maxime W Lafarge, Cor de Kroon, Gitte Ørtoft, Estrid Høgdall, Jouke Dijkstra, Jan J Jobsen, Ludy CHW Lutgens, Melanie E Powell, Linda R Mileshkin, Helen Mackay, Alexandra Leary, Dionyssios Katsaros, Hans W Nijman, Stephanie M de Boer, Remi A Nout, Marco de Bruyn, David Church, Vincent THBM Smit, Carien L Creutzberg, Viktor H Koelzer, Tjalling Bosse

## Abstract

Predicting distant recurrence of endometrial cancer (EC) is crucial for personalized adjuvant treatment. The current gold standard of combined pathological and molecular profiling is costly, hampering implementation. We developed HECTOR (**H**istopathology-based **E**ndometrial **C**ancer **T**ailored **O**utcome **R**isk), a multimodal deep learning prognostic model using hematoxylin-and-eosin-stained whole-slide-images and tumor stage as input, on 1,912 patients from seven EC cohorts including the PORTEC-1/-2/-3 randomized trials. HECTOR demonstrated C-indices in internal (*n* = 353) and external (*n* = 151) test sets of 0.788 and 0.816 respectively, outperforming the current gold-standard, and identified patients with markedly different outcomes (10-year distant recurrence-free probabilities of 97.0%, 77.7% and 58.1% for HECTOR low, intermediate and high risk groups). HECTOR also predicted adjuvant chemotherapy benefit better than current methods. Morphological and genomic feature extraction identified correlates of HECTOR risk groups, some with therapeutic potential. HECTOR improves on the current gold-standard and may help delivery of personalized treatment in EC.

## Introduction

Endometrial cancer (EC) is the most common gynecological malignancy in high-income countries and is increasing in incidence^1^. While most women with localized disease are cured by surgery, 10-20% develop distant recurrence which is typically incurable. Adjuvant chemotherapy can reduce this risk, at the expense of toxicity^2,3^. Thus, current guidelines recommend such treatment based on a combination of clinicopathological risk factors (e.g. histologic subtype, grade lymphovascular space invasion (LVSI), FIGO tumor stage) and, if available, the molecular classification of EC. The latter identifies patients with favorable and unfavorable outcomes defined by *POLE* mutation (*POLE*mut) or p53 abnormality (p53abn) respectively, and intermediate outcomes characterized by mismatch repair deficiency (MMRd) or no specific molecular profile (NSMP)^4–7^. However, in practice this remains challenging due to high-interobserver variability of histopathological factors, costs and turn-around-times of molecular testing, and lack of consensus on combining these risk factors. In addition, histological slides contain lots of visual information, some with prognostic potential^8^, that is only partly captured in the grading and tumor histotyping by pathologists.

Deep learning (DL) models, including those using digitized hematoxylin-and-eosin-stained (H&E) tumor slides have shown great promise in prediction of molecular alterations^9–11^, cell composition^12^, and prognosis^13–18^, outperforming standard pathologist-based assessment. This is particularly true of the latest generation of self-supervised learning and whole slide image (WSI) prediction DL models which use attention-based networks^19^, graphs^13,17^, or vision transformers^20,21^ to provide more granular and interpretable image representation. Additionally, multimodal DL models for prognosis prediction are promising to outperform unimodal approaches that solely rely on the morphological information provided by H&E WSIs^14^. We previously developed a DL model, im4MEC, to accurately predict the molecular EC classification from tumor H&E WSIs, and showed that image-based molecular classes predicted prognosis^9^. Others have used DL models to predict EC recurrence^22^ or overall survival^13,14,16,17,23^, but these have relied on more detailed tumor profiling, such as multiplex immunofluorescence staining^22^ or the combination of H&E WSIs with genomic and/or transcriptomic data^14^, neither of which are deliverable in clinical practice at present. Thus, there remains a pressing unmet need for a method that can predict EC distant recurrence from input data generated as part of routine clinical diagnostics.

Here, we report the development and evaluation of HECTOR (**H**istopathology-based **E**ndometrial **C**ancer **T**ailored **O**utcome **R**isk) (Fig. 1) – a multimodal DL model to predict EC distant recurrence from H&E WSI and tumor stage – across seven EC cohorts including three large randomized trials^2,24–28^.

**Fig. 1:**
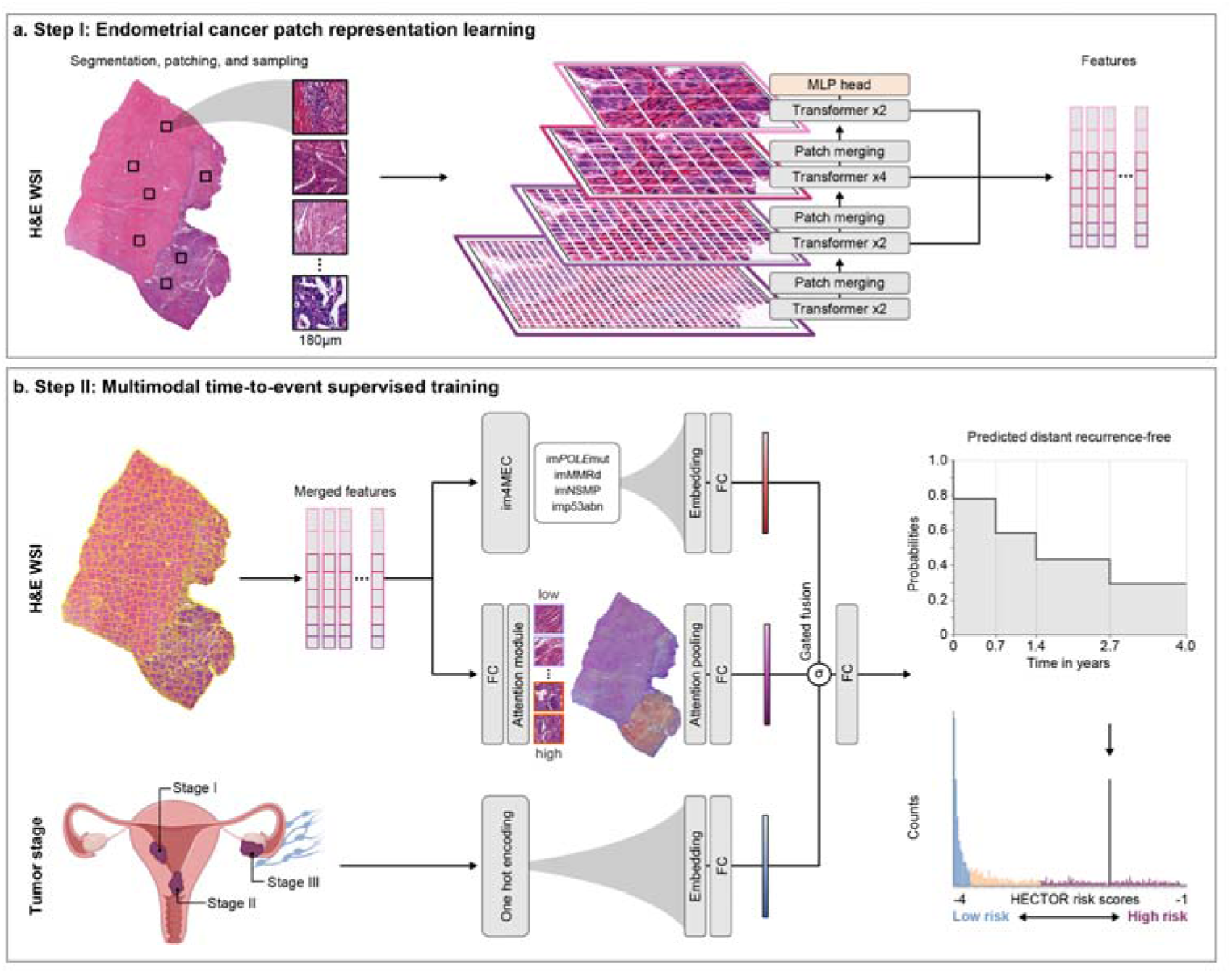
Overview of HECTOR. **a**, Tissue was segmented from the H&E whole slide image (WSI) of endometrial cancer (EC) and subsequently patched at 180μm. A multi-stage vision transformer^45^ was trained with self-supervised learning by randomly sampling patches from WSIs of 1,862 patients, excluding any patients of the internal and external test sets. Patch-level features are extracted from the last eight transformer blocks. **b,** HECTOR takes the H&E WSI and the (FIGO 2009) tumor stage I-III category^31^ as inputs. Extracted patch-level features are spatially and semantically averaged. The patch features are passed into both attention-based multiple instance learning module model, and im4MEC (with all layers frozen), a deep learning model predicting the molecular class from H&E WSI as image-based (im)*POLE*mut, imMMRd, imNSMP or imp53abn^9^. Both the tumor stage category and image-based molecular class are fed through the embedding layers. Gating-based attention is applied on the resulting three embeddings^14,32^, followed by a Kronecker product for fusion. The negative log-likelihood was used to predict the distant recurrence-free probabilities function over discrete time^46^. Risk scores were defined as the integrated predicted survival probabilities. MLP = multilayer perceptron; *POLE*mut = *POLE* mutated; MMRd = mismatch repair deficient; p53abn = p53 abnormal; NSMP = No specific molecular profile; im = image-based; FC = fully connected layer.

## Results

### Endometrial cancer cohorts

HECTOR is a two-step DL model wherein the first step consists of self-supervised tumor image representation learning, and the second of the distant recurrence prediction task (Fig. 1).

To train and validate the distant recurrence prediction task of HECTOR, we collected and curated H&E-stained WSIs and comprehensive clinicopathological datasets, molecular and clinical distant recurrence data for 1,912 patients with tumor stage (2009 FIGO) I-III EC across seven cohorts, including the PORTEC-1, –2 and –3 randomized trials^2,24–28^ (Extended Data Fig. 1; study CONSORT diagram shown as Supplementary Fig. 1,2, Table 1,2). Of these, the patients from the Leiden cohort (*n* = 151 patients) were held out as an external test set. The remaining patients were divided randomly into a 20% held-out internal test set (*n* = 353) and 80% training set (*n* = 1,408) where five-fold cross-validation was performed. The median duration of follow up in the training set, internal test set and external test was 7.8, 8.4 and 2.9 years respectively, during which 246 (17.5%), 62 (17.6%), and 24 (15.9%) patients had distant recurrence. Importantly, patients who underwent chemotherapy, predominantly the experimental treatment-arm of the PORTEC-3 randomized trial (*n* = 224), were excluded from training as this treatment influences distant recurrence^2,3^ (Extended Data Fig. 1). These PORTEC-3 patients were however used for downstream analysis of treatment benefit by HECTOR.

To train HECTOR’s self-supervised learning step (which requires a large imaging dataset without outcome data), we enriched the training set with TCGA-UCEC^29^ (The Cancer Genome Atlas Uterine Corpus Endometrial Carcinoma) WSIs and the WSIs that were excluded for the distant recurrence task due to FIGO stage IV or missing outcome (*n* = 1,862; Methods).

Altogether, including the two training steps and the downstream analyses, this study comprised tumor data of 2,590 patients.

### HECTOR design and performance

To design HECTOR and obtain the most performant DL model for prediction of distant recurrence based on the highest Concordance-index^30^ (C-index), we conducted ablation studies on the five-fold cross-validation (Supplementary Table 3). HECTOR’s first step comprises a vision transformer for patch-level self-supervised learning representation (Fig. 1a). HECTOR’s second step is a novel multimodal three-arm architecture to predict distant recurrence-free probabilities (Fig. 1b). The three-arm architecture fuses prognostic information from the H&E-stained WSI, the image-based (im) molecular class as predicted by im4MEC directly from the H&E-WSI^9^, and the tumor stage (2009 FIGO I-III)^31^. To do this, we combined attention-based multiple instance learning with a novel method which used embedding layers to map the discrete risk factors (the image-based molecular class and tumor stage) to a higher-dimensional continuous vector space, with the importance of each factor controlled by gating-based attention^14,32^. Further details are provided in Methods.

HECTOR demonstrated a mean C-index of 0.795±0.031 on five-fold cross-validation. Notably, the addition of the image-based molecular class arm as predicted by im4MEC to the H&E WSI (referred to as two-arm or one-arm model respectively) boosted performance from 0.775±0.031 to 0.782±0.026 with no need for extra input data. Adding the tumor stage (FIGO I-III) further improved the C-index to 0.795±0.031 yielding the final architecture of HECTOR (Fig. 2a). The cumulative area under the receiver operating curve^33^ and Integrated Brier Score^34^ are reported in Supplementary Table 4. We also observed that HECTOR concentrated high attention to fewer regions while ignoring large parts of the H&E WSI compared to a model relying on the H&E WSI (Extended Data Fig. 2).

**Fig. 2:**
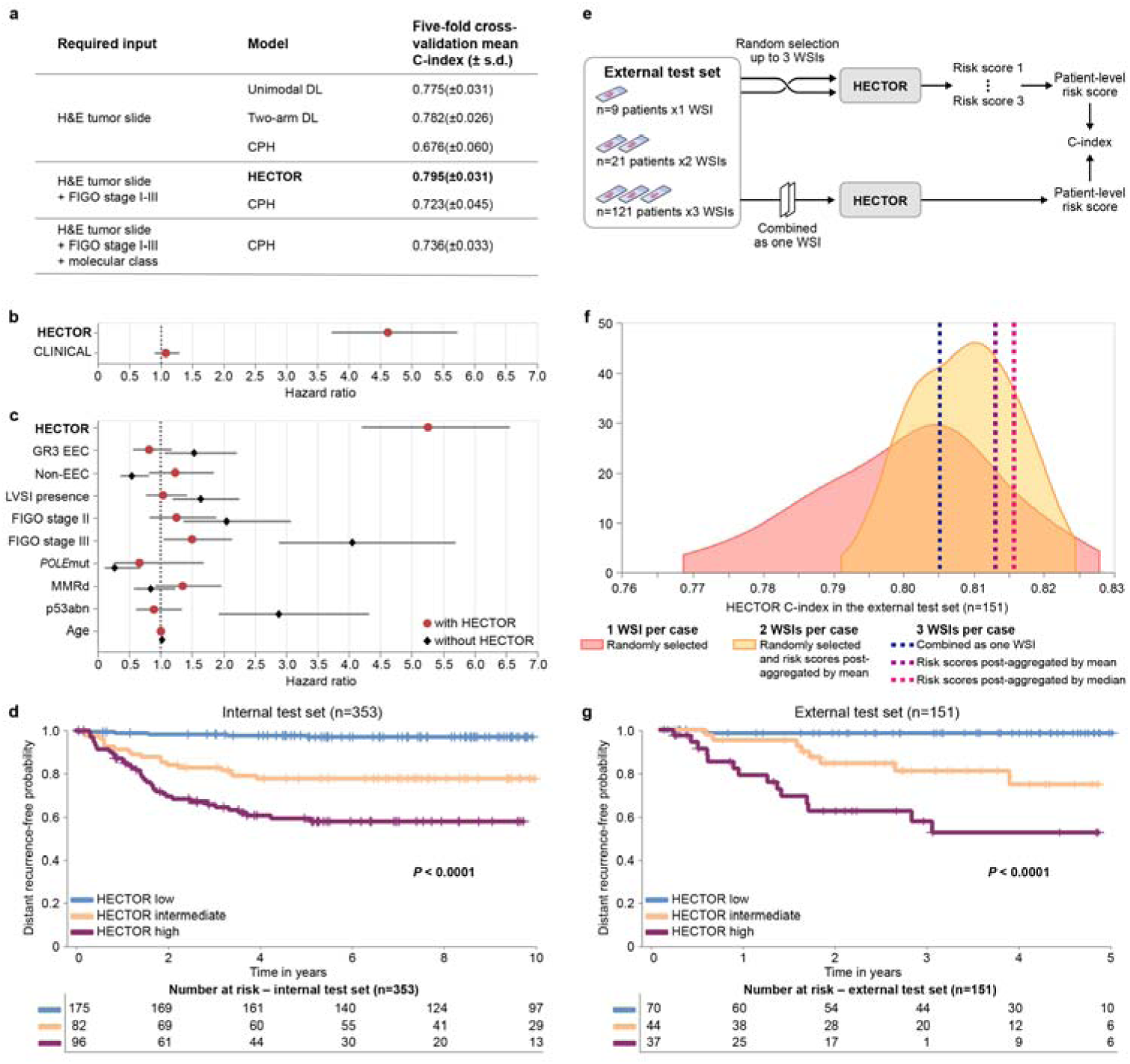
Performance of HECTOR. **a**, Comparison of HECTOR performance using the C-index with alternative deep learning-based unimodal and multimodal models and Cox Proportional Hazard models. **b**, Comparison of prognostic values between HECTOR and clinicopathological and molecular risk factors combined into one risk score in a multivariable analysis. **c**, Residual prognostic value of all established clinicopathological and molecular risk factors when using HECTOR predicted risk scores in a multivariable analysis. **d,** 10-year distant recurrence-free probabilities analysis using the Kaplan-Meier method by HECTOR risk groups in the internal test set and log-rank test *P* values. **e,** Experiments conducted in the external test set with the input of multiple WSIs. **f**, C-index of HECTOR in the external test set using one to three WSIs per patient. **g**, Five-year distant recurrence-free probabilities analysis using the Kaplan-Meier method by HECTOR risk groups when using up to three WSIs (post-aggregated by median) in the external test set and log-rank test *P* values. G3 = grade 3; EEC = endometrioid endometrial cancer; *POLE*mut = *POLE* mutated; MMRd = mismatch repair deficient; p53abn = p53 abnormal; LVSI = lymphovascular space invasion. TCGA-UCEC = The Cancer Genome Atlas Uterine Corpus Endometrial Cancer. EBRT = External beam radiotherapy, VBT = Vaginal brachytherapy. MIL = Multiple Instance learning. LVSI = lymphovascular space invasion; s.d. = standard deviation; H&E = hematoxylin-and-eosin; WSI = whole slide image.

On the unseen internal test set, HECTOR obtained a C-index of 0.788.

Univariable analysis, using a Cox proportional hazard model with continuous HECTOR risk score as the independent variable and time to distant recurrence as the dependent variable, showed strong prognostic value of HECTOR risk score as a continuous variable (hazard ratio (HR) = 4.92; 95% confidence interval (CI): 4.20-5.76; *P* = 9.72e-87).

To aid clinical interpretation, we defined categorical HECTOR risk groups at quartiles of the continuous risk scores in the training set. We combined groups from the first two quartiles as these had very similar clinical outcomes in the training set (distant recurrence-free probabilities of 98.1% and 95.8% respectively; Supplementary Fig. 3). We then tested the prognostic value of these groups on the internal test set. 10-year distant recurrence-free probabilities for HECTOR low (*n* = 175), intermediate (*n* = 82), and high (*n* = 96) risk groups were 97.0% (95% CI: 0.930-0.988), 77.7% (95% CI: 0.670-0.854) and 58.1% (95% CI: 0.469-0.677) respectively (log-rank *P* < 0.001; Fig. 2d). Corresponding HR for HECTOR high and intermediate risk groups using the HECTOR low risk group as the reference were 26.18 (95% CI: 16.26-42.15; *P* = 3.55e-41) and 4.00 (95% CI: 2.29-6.97; *P* = 1.04e-06), respectively.

### Comparison with current prognostic gold standard

We compared DL-based risk scores (that is the one-, two-arm and HECTOR model) with the current standards for EC prognostication comprising clinicopathological risk factors and the molecular EC classification on the five-fold cross-validation (Fig 2a.). For this, we first compared C-indices by type of input required: (i) a ‘base’ Cox model including variables defined by pathologist using H&E images alone (histologic subtype, grade and LVSI); (ii) the base model plus tumor stage; (iii) the base model plus tumor stage and molecular EC class. In the five-fold cross-validation, given H&E-based input data, the one– and two-arm model discrimination was superior to the base Cox model (C-index = 0.676±0.060). HECTOR model discrimination was superior to the base Cox model plus tumor stage which used the same inputs (C-index = 0.723±0.045), and better or as good as the base Cox model plus tumor stage and molecular EC class (C-index = 0.736±0.033), which requires sequencing, immunohistochemistry, and expert pathology.

We further compared HECTOR prognostic value against current clinicopathological and molecular risk factors in multivariable analysis using HECTOR continuous risk scores as the independent variable. HECTOR retained prognostic value in multivariable models in which known risk factors (histological subtype, grade, LVSI, 2009 FIGO I-III tumor stage, age, molecular class) combined as one risk score (referred to as CLINICAL risk score), were not prognostic (HECTOR HR = 4.62 (95% CI: 3.72-5.73; *P* = 5.02e-44) versus CLINICAL HR = 1.08 (95% CI: 0.90-1.30; *P* = 0.402)) (Fig. 2b). Similar multivariable analysis including risk factors as individual variables showed independent prognostic value of HECTOR (HR = 5.26; 95% CI: 4.21-6.56; *P* = 2.30e-48), with only FIGO tumor stage III disease retaining statistical significance (HR = 1.50; 95% CI: 1.05-2.14; *P* = 0.026) (Fig. 2c). Other known risk factors were no longer prognostic after inclusion of HECTOR risk score suggesting that these factors were captured by HECTOR. For instance, the *POLE*mut and p53abn molecular classes derived from ground-truth sequencing and immunohistochemistry respectively (HR = 0.66 (95% CI: 0.26-1.69; *P* = 0.384) and HR = 0.90 (95% CI: 0.61-1.34; *P* = 0.616) respectively) and histological factors such as LVSI (HR: 1.05 95% CI: 0.77-1.42, *P* = 0.776) would not be of additive prognostic value for the prediction of distant recurrence.Given current prognostic gold standards which would classify p53abn EC as high-risk tumors and MMRd and NSMP as intermediate-risk tumors with heterogeneous outcomes, we validated the capacity of HECTOR to refine prognosis within the MMRd, NSMP and p53abn molecular classes in the training and internal test sets. In particular, the HECTOR low risk group also identified about 5.3% (16 out of 300) of p53abn EC cases with excellent prognosis in the entire dataset (Supplementary Fig. 4). Along these lines, we estimated number of patients with markable different risk classification between HECTOR and the ESGO-ESTRO-ESP 2021 guidelines^4^ which combines clinicopathological and molecular factors (Supplementary Fig. 5). Among all patients with intermediate to high-risk tumors based on the guidelines (and no report of distant recurrence), 48.2% (552 cases out of 1,146) patients were predicted HECTOR low risk, and 16.9% (62 cases out of 366) among high-risk tumors only. Among all guideline-based low to high intermediate-risk tumors, 11.2% (131 out of 1,170) patients were predicted HECTOR high risk, and 4.9% (14 out of 287) when restricting to only low risk tumors.

### Performance on the external test set including multiple WSIs

We next evaluated HECTOR prognostic value and robustness in the real-world external test set. For this, we leveraged the fact that most cases in this cohort had multiple tumor containing H&E WSIs derived from different tissue blocks per patient (121 of 151 cases had three WSIs; 21 had two and 9 had one; Fig. 2e), enabling us to first evaluate external performance of HECTOR and subsequently test robustness to selection of H&E WSI. Initial evaluation using HECTOR score derived from multitude random selection of a single WSI per patient demonstrated model C-index of 0.802±0.013 for prediction of distant recurrence (Fig. 2f).

HECTOR performance and risk stratification was slightly improved by the addition of further WSIs, (taking per-patient HECTOR risk scores as either the mean or median scores across WSIs) with C-indices of 0.809±0.007 with up to two WSIs per patient, and 0.813 or 0.816 with up to three WSIs (Fig. 2f). A different method was tested wherein the WSIs were combined as one single input bag of images, yielding a C-index of 0.805. The five-year distant recurrence-free probabilities using the median of HECTOR risk scores per patient were 98.4% (95% CI: 0.891-0.998) in HECTOR low risk (*n* = 70), 74.8% (95% CI: 0.534-0.874) in HECTOR intermediate risk (*n* = 44), and 52.6% (95% CI: 0.323-0.694) in HECTOR high risk (*n* = 37), (log-rank *P* < 0.001) (Fig. 2g, Supplementary Fig. 6). We explored potential confounding by intratumoral heterogeneity causing different risk scores across individual patient WSIs (Supplementary Fig. 7-10, Notes p14). Specifically, 85 cases out of the 142 cases with more than one WSI had consistent HECTOR risk group predictions across the WSIs and only three cases with three WSIs had a different predicted HECTOR risk group for each WSI.

### Association with clinicopathological data and input contribution

DL prognostic models may provide information on the correlates or features which determine clinical outcome. Initial analysis of the internal test set by multiple linear regression (Fig. 3a,b) revealed that lower HECTOR risk scores were associated with established favorable risk factors of endometrioid (EEC) histological subtype, grade 1, and *POLE*mut EC, and higher HECTOR risk scores with unfavorable factors including non-EEC histological subtypes, grade 3, FIGO stage III, LVSI, p53abn EC, estrogen receptor (ER) negativity, and L1CAM positivity (Supplementary Table 6-8, Fig. 11). MMRd EC, grade 2 and FIGO stage II were spread throughout the risk score axis and were not significant.

**Fig. 3:**
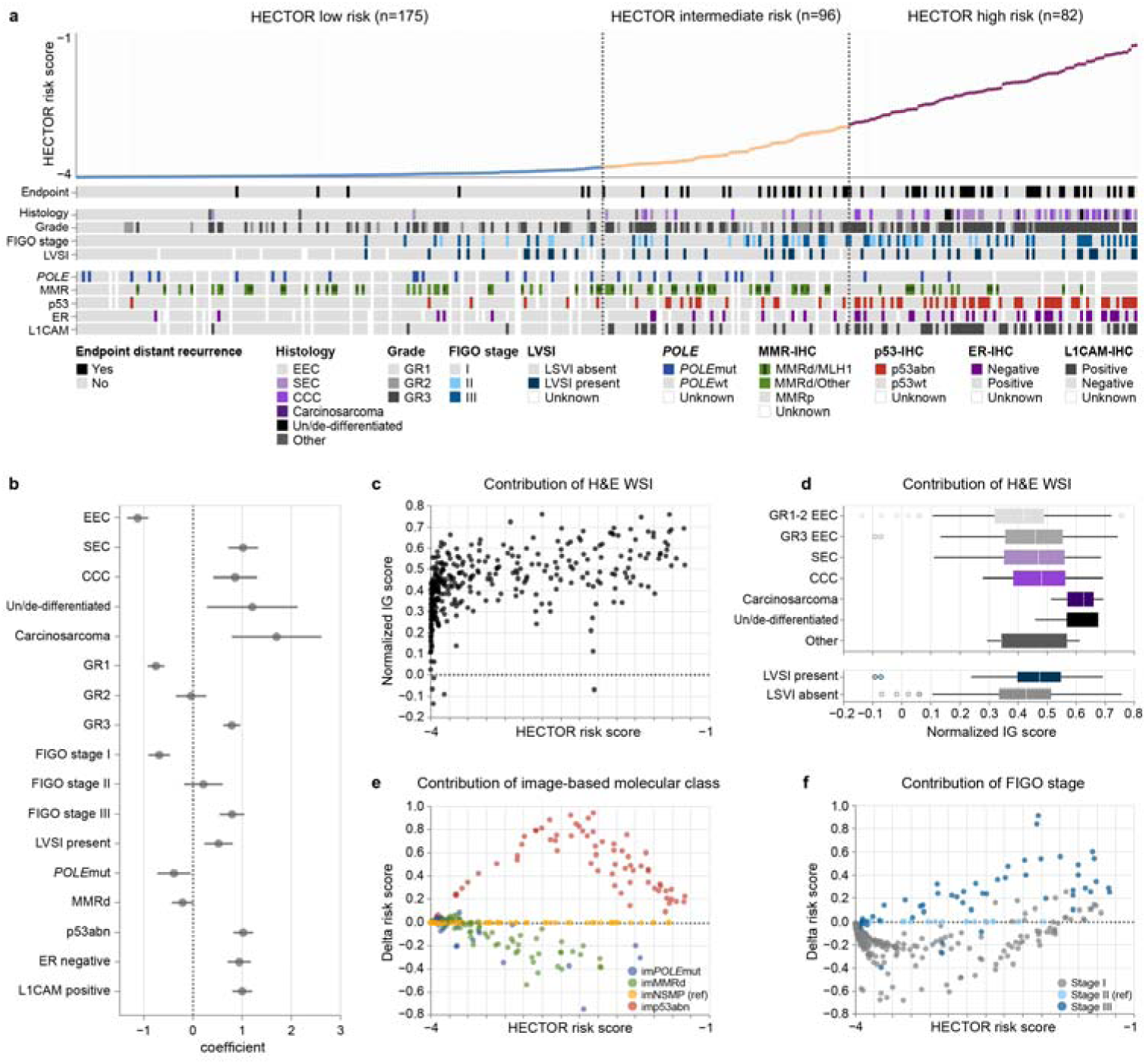
HECTOR explainability by analysis of HECTOR risk group composition with clinicopathological and molecular data and analysis of input contribution to the HECTOR risk scores. **a**, Heatmap of prognostic factors for patients included in the internal test set (*n* = 353 patients) ordered by predicted risk scores. **b**, Association of the clinicopathological data and continuous HECTOR risk scores using single linear regression. **c**, Analysis of the contribution to the HECTOR risk scores of the whole slide image (WSI) modality in the internal test set using the Integrated Gradients (IG) method^47^. **d**, Integrated gradient normalized values of the WSIs stratified by histological subtypes (top) and presence of lymphovascular space invasion (LVSI) (bottom). **e**, The contribution of the image-based molecular classes to the continuous HECTOR risk score in the internal test set, using the imNSMP as reference (ref) group. The difference in predicted risk score is computed between the risk score given by the image-based molecular class and the one produced by imNSMP. **f**, The contribution of 2009 FIGO stage to the continuous HECTOR risk score in the internal test set, using 2009 FIGO stage II as reference (ref) group. *POLE*mut = *POLE* mutated; MMRd = mismatch repair deficient; p53abn = p53 abnormal; ER = estrogen receptor; EEC = endometrioid EC; SEC = serous; CCC = clear cell; GR1-3 = grade 1-3.

For deeper explainability, we evaluated the impact of the H&E WSI, im4MEC, and tumor stage on the prediction, that is whether each modality decreased (negative contribution) or increased (positive contribution) the HECTOR risk scores of developing distant recurrence. We used the normalized Integrated Gradients values for the H&E WSIs, and differences in predicted risk scores with fixed value of im4MEC or FIGO tumor stage for the same case in the internal test set (Fig. 3c). The H&E WSIs mainly had a positive contribution with values linearly increasing alongside HECTOR risk scores (Fig. 3d; Supplementary Fig. 12). We also noted higher magnitude of contributions towards Grade 3 EEC or non-EEC histological subtypes and LVSI (Fig. 3e), indicating the strong influence of unfavorable WSI-based features. The use of image-based molecular class and FIGO stage I-III was consistent with domain expertise in EC with im*POLE*mut and imMMRd mainly decreasing and imp53abn strongly increasing the HECTOR risk scores, and higher tumor stage increasing the HECTOR risk scores (Fig. 3f,g, Supplementary Fig. 13,14).

These analyses enabled us to dissect data of the six patients with distant recurrence predicted as HECTOR low risk in the internal test set (Supplementary Table 9, Fig. 15). Experimental tests, in which the image-based molecular class was replaced by the true molecular class, showed no effect of misclassification by im4MEC in these instances onto the HECTOR risk group. Review of the single WSI input by an expert gyneco-pathologist revealed that at least in two cases WSIs were missing unfavorable visual features that were reported in the pathology report (substantial LVSI or high-grade tumoral areas).

### Morphological correlates of outcome risk

To identify the prognostic morphological features that may have been used by HECTOR, the top 5% regions of the H&E WSIs with the highest impact on the risk scores (decreasing and increasing) were extracted and reviewed by an expert gyneco-pathologist in the internal test set (Fig. 4a; Supplementary Fig. 16-20). Within the HECTOR low risk group, the morphological features decreasing the risk score were identified as smooth luminal borders, inflamed stroma and intraepithelial lymphocytes, intraepithelial neutrophils, and abundant compact normal myometrium without tumor. Morphological features increasing the risk score in the HECTOR high risk group were a ragged luminal tumor surface (also referred to as hobnailing), LVSI, solid tumor growth with marked nuclear atypia, desmoplastic stromal reaction, and presence of mitotic figures (Fig. 4a). Within the HECTOR low risk group, we observed morphological features with positive contribution, though relatively less common, as surface changes mimicking hobnailing, retraction artifacts mimicking LVSI, loose myometrium with edema mimicking desmoplasia and solid tumor growth with scattered high-grade atypia (Extended Figure Data 3a.).

**Fig. 4:**
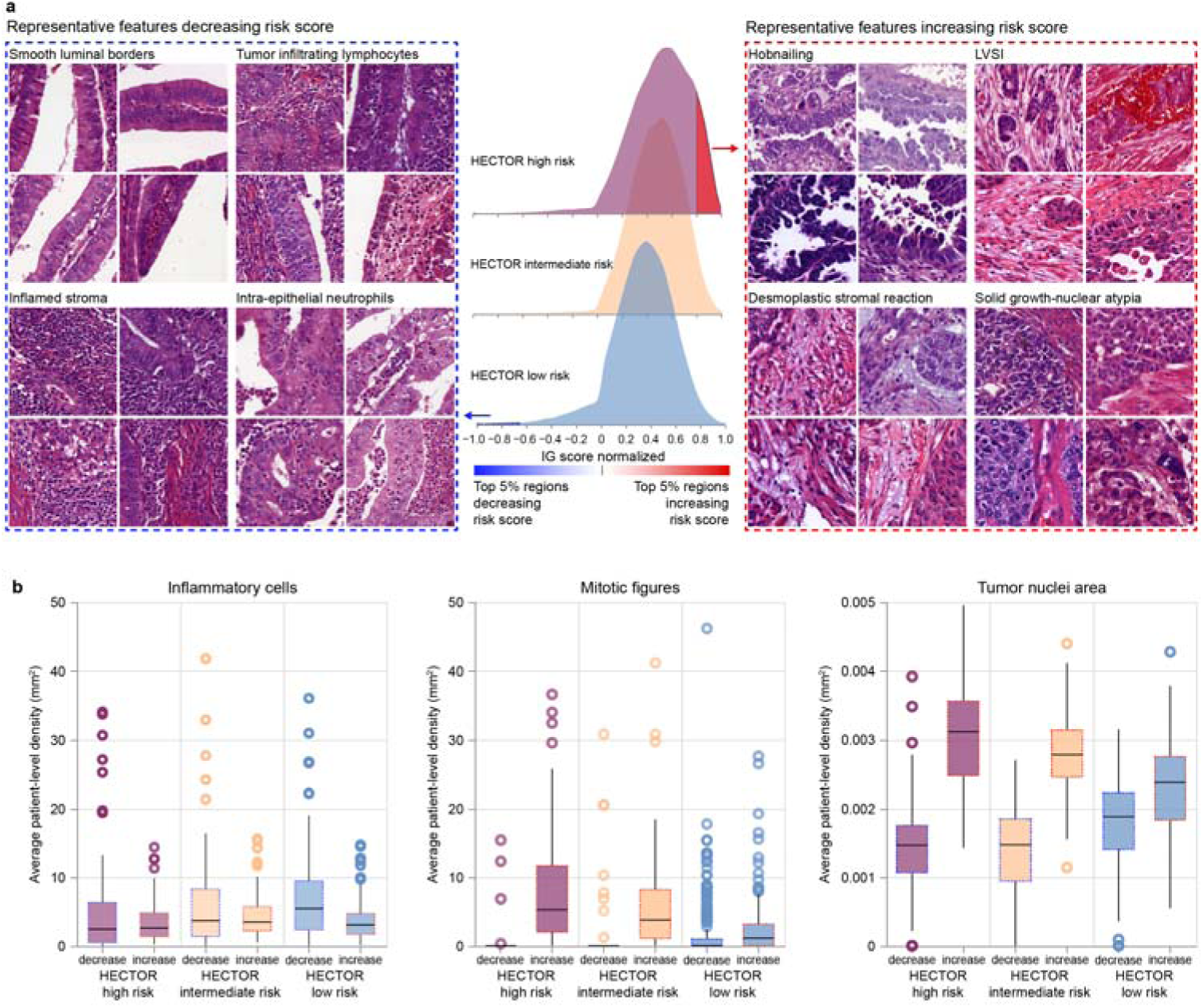
Morphological features contributing to HECTOR risk scores. **a**, The top 5% of the regions increasing and decreasing the risk score, from the Integrated Gradient (IG) method^47^, were extracted for qualitative review and quantitative analysis. A representative selection of four patches for each morphological subtype (each selected from a different patient) increasing the risk score in the HECTOR high risk group (top right). A representative selection of four patches for each morphological subtype (each selected from a different patient) decreasing the risk score in the HECTOR low risk group (top left). **b**, Among the top 5% regions decreasing and increasing the risk score, inflammatory cells, and mitotic figures were detected and counted with deep learning-based image analysis tools^12,48^, as well as the size of tumor nuclei.

Mitotic activity, inflammatory cell density, and the size of the tumor nuclei were quantified using deep learning-based image analysis tools (Fig. 4b; Methods). More inflammatory cells were present in the top 5% regions decreasing the risk scores and this effect was more pronounced in the HECTOR low risk group (*P* = 0.011). A higher mitotic density and larger tumor nuclei were found in the top 5% regions in the HECTOR high risk group (both *P* < 0.001). These results remained consistent across image-based molecular classes and FIGO stage I-III (Supplementary Figs. 21-23), and when filtering in regions containing tumor cells (Supplementary Fig. 24). In a quantitative spatial analysis, we computed the overlap of the top 5% regions with the tumor and invasive border areas (Extended Figure Data 3b.). The latter showed regions increasing the risk scores were picked out more from the tumor than from the invasive border area. Tumor and invasive border areas contributed nearly the same in regions decreasing the risk scores, notably in the HECTOR low risk group.

### Genomic alterations, immune and tumor-related transcriptional signatures

For comprehensive analysis of the molecular correlates of HECTOR risk scores, we analyzed the TCGA-UCEC (*n* = 381 FIGO stage I-III ECs) dataset (Fig. 5; Supplementary Fig. 25). Coding driver mutations in *ARID1A*, *CTCF*, *CTNNB1*, *FGFR2*, *KRAS,* and *PTEN* were enriched in HECTOR low risk group (all *P <* 0.005), while *PPP2R1A* and *TP53* mutations were more frequent in the HECTOR high risk group (*P* = 2.19e-03 and *P* = 2.81e-07 respectively) (Fig. 5a; Supplementary Table 10). Using transcriptional data, we performed an analysis of CIBERSORT-defined lymphocyte populations using multiple linear regression (Fig. 5b). This revealed that increasing HECTOR scores were positively correlated with memory B-cells (*P* = 0.008), activated dendritic cells (*P* < 0.001), and resting mast cells (*P* = 0.029), and inversely correlated with CD8+ T-cells (*P* < 0.001), follicular helper T-cells (*P* < 0.001), regulatory T-cells (*P* < 0.001), and NK cell activation (*P* = 0.049). Notably, these associations were independent of EC molecular class and tumor mutational burden (Supplementary Table 11). Further transcriptomic analysis (Fig. 5c, Supplementary Fig. 25c) confirmed variation in lymphocyte populations was reflected in differential expression of canonical immune cell markers including *CD1C*, *BTLA* and *CD40LG* (enriched in the HECTOR low risk cases). This also demonstrated that HECTOR high risk tumors demonstrated upregulation of genes predictive of worse outcomes in EC, including *L1CAM* and Claudin-6 (*CLDN6*), while HECTOR low risk cases showed upregulation of genes associated with hormone signaling (*C1orf64*; *OVGP1*).

**Fig. 5:**
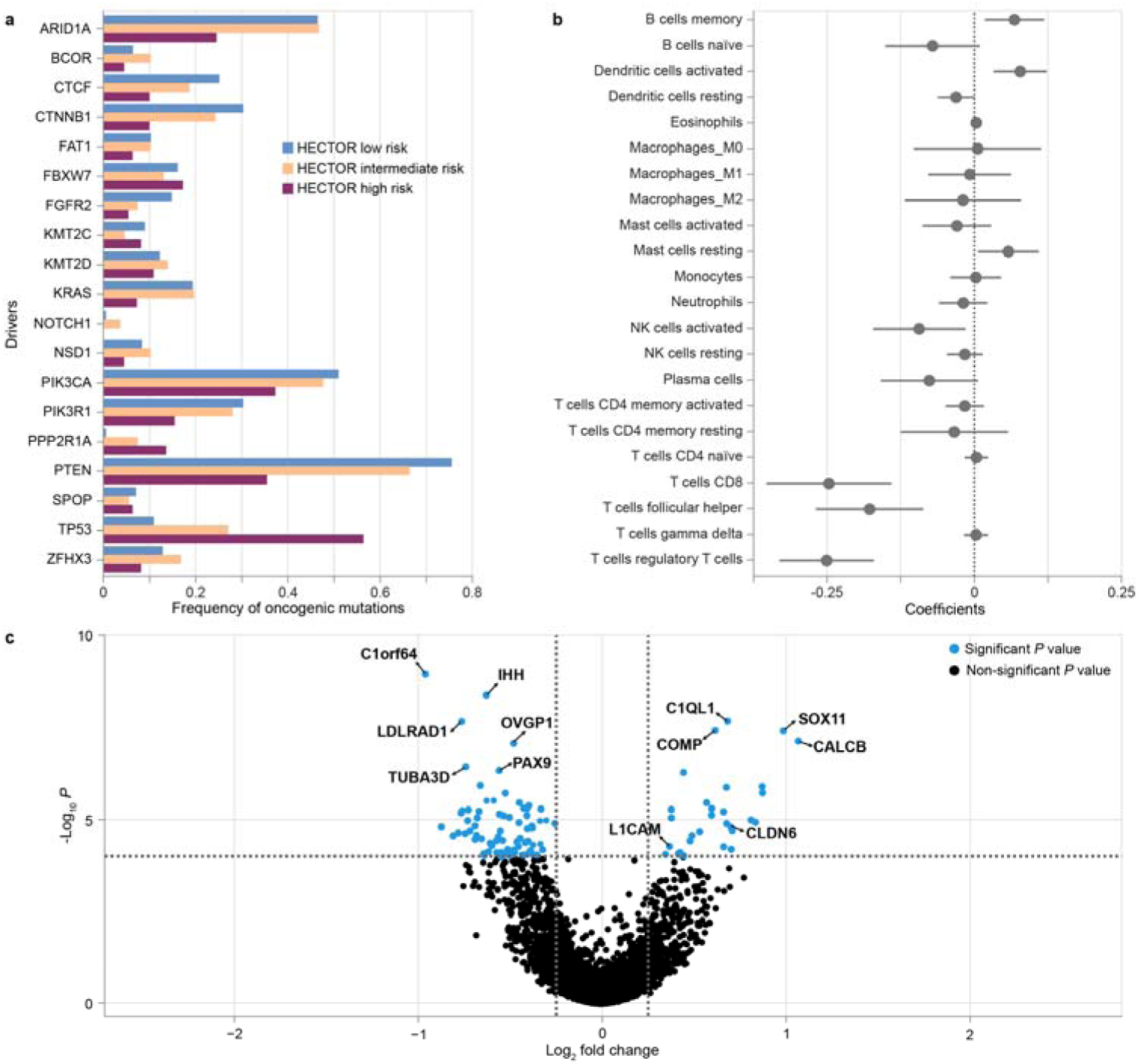
Genomic and transcriptomic correlations of HECTOR risk groups using TCGA-UCEC (*n* = 381). **a**, Analysis of the mutational frequency of the top-20 genes recognized as key oncogenic alterations in endometrial cancer for each HECTOR risk group. **b**, Association of HECTOR risk score with the immune activation gene. **c**, Differential gene expression of HECTOR high-risk versus HECTOR low-risk TCGA-UCEC cases.

### Adjuvant treatment response prediction by HECTOR

The investigation whether HECTOR could predict benefit of chemotherapy for distant recurrence risk was conducted using the PORTEC-3 randomized trial. In this trial, patients with high-risk stage I-III EC were randomized to concurrent and adjuvant external beam radiotherapy with or without platinum and paclitaxel-based chemotherapy. HECTOR risk scores were predicted on all PORTEC-3 cases whom WSI was available (*n* = 441), which included the patients who underwent chemotherapy (*n* = 225). Importantly, these 225 cases had not been used in either training or test sets (Extended Data Fig. 4, Supplementary Table 12, Fig. 26). Analysis of distant recurrence-free probabilities by treatment arm and HECTOR demonstrated a statistically significant interaction between chemotherapy and HECTOR risk score as either continuous or categorical variable (*P*_INTERACTION_ = 0.014 and *P*_INTERACTION_ = 0.064 respectively).

We examined this in detail across HECTOR risk groups (Fig. 6a). Within HECTOR low (*n* = 92) and HECTOR intermediate risk (*n* = 177) groups, outcomes were similarly favorable in both treatment arms, as evidenced by similar probability of EC distant recurrence (log-rank *P* = 0.244 and 0.807 respectively). In contrast, among women classified as HECTOR high risk, those who received chemotherapy had significantly improved distant recurrence-free probabilities as compared to those treated with external beam radiotherapy alone (five-year distant recurrence-free probability of 62.2% (95% CI: 0.511-0.715) versus 42.0% (95% CI: 0.311-0.526); log-rank *P* = 0.007; HR = 0.561 (95% CI: 0.366-0.862; *P* = 0.008)). Exploratory analysis suggested that the predictive accuracy was greater than that provided by prognostic factors currently used to identify patients with high-risk tumors who were likely to benefit from chemotherapy, including serous histological subtype, FIGO stage III and the p53abn molecular class (Fig. 6b). Further exploratory analyses suggested that HECTOR identified patients who benefited from adjuvant chemotherapy within the NSMP and MMRd molecular classes (Supplementary Fig. 27,28). These results remained consistent when sub-stratifying by the image-based molecular class arm of HECTOR (Supplementary Fig. 29). Thus HECTOR demonstrated significant predictive utility that may exceed that offered by current methods.

**Fig. 6:**
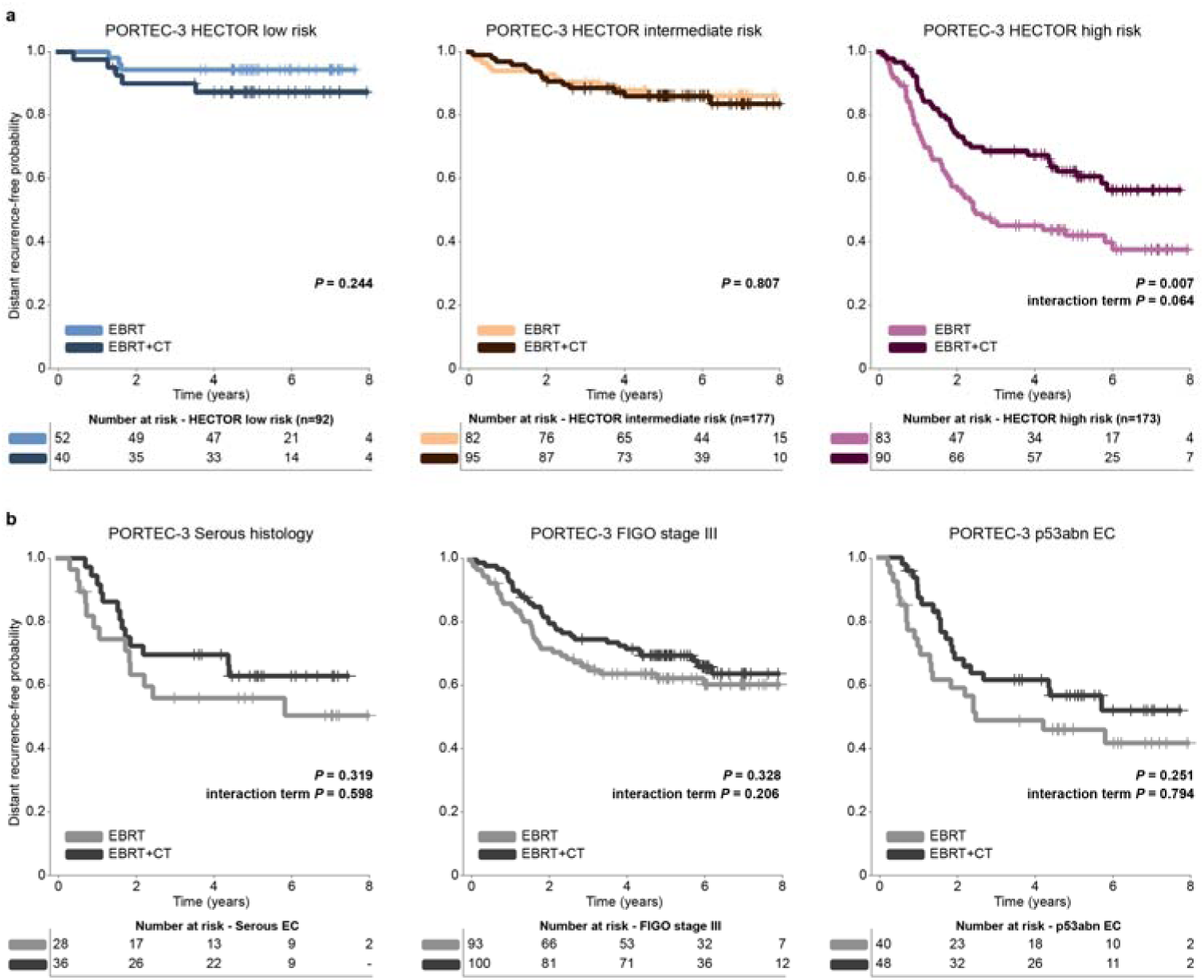
Impact of the addition of adjuvant chemotherapy to external beam radiotherapy on distant recurrence in the PORTEC-3 randomized trial by HECTOR risk group. **a**, Eight-year distant recurrence-free probability by Kaplan-Meier’s analysis, and log-rank test *P* value is shown for each HECTOR risk groups stratified by randomly allocated treatment. *P* value of the interaction term using categorical HECTOR risk group is shown. There was also a significant interaction between the HECTOR continuous risk scores and the treatment (*P*_INTERACTION_ = 0.014). **b**, For comparison with HECTOR selection, distant recurrence probability by Kaplan-Meier’s analysis from the PORTEC-3 study is shown, for different gold standard prognostic factors in EC relying on serous histology, 2009 FIGO stage III, and the p53abn molecular class respectively. The log-rank test and interaction term *P* values are displayed.

## Discussion

HECTOR, a DL model trained and validated in 1,912 patients^2,24–28^, accurately predicts EC distant recurrence risk using only H&E-stained tumor slide(s) and tumor stage. HECTOR outperformed the current gold standard of combined pathological and molecular analysis for distant recurrence prediction, and was also found predictive of chemotherapy benefit in the PORTEC-3 trial. Our results suggest that HECTOR has the potential to be a highly effective tool for individualized prognostication of women with EC while delivering shorter turnaround times and reducing testing costs. HECTOR may also enable new biomarker discoveries for improving targeted treatment decision-making.

HECTOR performance results from a novel multimodal integrative three-arm architecture which leveraged prognostic information from the H&E WSI, the im4MEC image-based molecular class, and FIGO 2009 stage. This multimodal architecture outperformed alternative DL models using only H&E-based information, corroborating other studies^14,35^. Interestingly, nesting of the pre-trained im4MEC model within HECTOR (an approach inspired by multi-task learning^36^) boosted HECTOR performance, in contrast to other studies where integration of copy number variation or transcriptomics did not improve prediction of overall survival in EC^14^. We demonstrated that the prognostic value of categorical risk factors, such as EC stage, can be learned end-to-end by the DL model to increase predictive accuracy, which may benefit similar studies in other cancer types where unimodal DL models have been developed on H&E WSI only^15,18^.

Our preliminary investigations of model explainability and risk score correlates offer good prospects to improve our understanding of the biology of EC and other cancer types. For example, the association of HECTOR low risk scores with immune cell infiltrate is consistent with data showing better prognosis of immune-infiltrated EC^8^, although at present it is unclear if HECTOR directly quantified lymphocyte subtypes such as T-cells from H&E WSIs. The upregulation of *CLDN6* in HECTOR high risk ECs is consistent with this as being a predictor of distant recurrence^36^. Cases with combined HECTOR high risk and *CLDN6* upregulated could be actionable as a chimeric antigen receptor (CAR) T cells target^37^. While desmoplastic stromal reaction is known to predict bad prognosis in colorectal cancer, the novel association we report here has not previously been reported in EC^38^. Whether this represents a morphological readout of *L1CAM* overexpression^39^ is presently unclear. We also confirmed well-established unfavorable histopathological risk factors in EC aligning with higher HECTOR risk scores^4^. Thus, we expect the outperformance of standard histopathology by HECTOR being likely driven by the non-linear combination of each factor and, more importantly, the non-categorical processing of the visual information from the WSIs.

HECTOR holds considerable promise for clinical application worldwide. The design consisting of using the image-based instead of the true molecular classes regards HECTOR as a relatively cost-effective and scalable prognostic tool as it requires two broadly available inputs, one digitized H&E-stained tumor slide and the tumor stage. We strongly foresee HECTOR being utilized to triage women with EC between low and high risk of distant recurrence, which subsequently provides a means to de-escalate treatment for the low risk group and select targeted adjuvant systemic therapy or perform individualized additional molecular tests for the high risk group. While our data supports that HECTOR may reduce under– and over-treatment for women with EC, it would also spare challenges and expenses of resource-limited environments where molecular testing and expert pathologist review are difficult or unfeasible. Towards future scalable technical improvements of HECTOR, the range of input could be extended to consecutive digitized H&E-stained tumor sections followed by 3D reconstruction^40^, routinely-performed IHC-stained WSIs^41^, or radiology images^42^.

Our study has several strengths. Our total cohort of 2,590 patients, including three randomized trials, makes this the largest DL-based prognostic study in EC performed to date. Our state-of-the-art multimodal DL methodology allowed us to leverage prognostic information from multiple factors, including those beyond the H&E image alone. Expert pathology review and molecular profiling enabled us to benchmark our novel methodology against the current gold-standard in risk stratification of EC. Limitations of our study are that our current model based on multiple instance learning is unaware of the spatial relationship between regions and was not designed to leverage information between multiple WSIs, both of which may improve performance^43,44^; although context-aware architectures have not been found to improve performance in this task. In addition, complex interactions between the morphology, molecular, and tumor stage may not be yet fully optimized as we limited experiments to late-fusion techniques^21,35^. Some patients in the study did not undergo surgical staging-lymphadenectomy^24,25^, a consideration which may create some noise in the tumor stage input and may explain the residual prognostic value of advanced disease stage in multivariable analysis. Furthermore, not all morphological correlates observed in the H&E regions, (e.g. structural changes), were quantified in this study due to the lack of available labeled datasets which could have been used for training deep learning-based EC-specific image analysis tools. Finally, our results need further validation in prospective trials, and in cohorts more diverse than the ones of largely European ancestry we examined.

In summary, HECTOR is a DL-based prognostic tool requiring only a scanned H&E tumor slide and tumor stage, which outperforms the current gold standard of risk assessment with combined pathologist evaluation and molecular testing. Validation and extension of HECTOR could help delivery of precision medicine to advance prognostication, with improvement on both targeted systemic therapy recommendation and treatment de-escalation of women with EC worldwide.

**Extended Data Fig 1:**
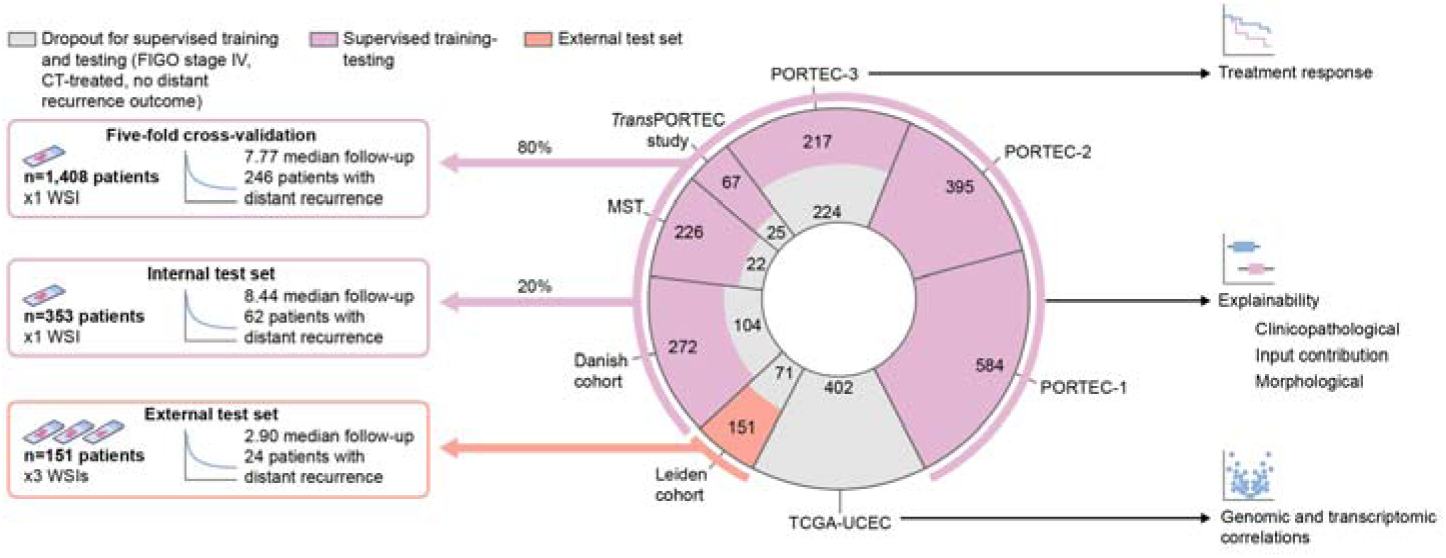
Overview of the data split and downstream analyses performed in this study. One representative WSI per patient from an Formalin-Fixed Paraffin-Embedded (FFPE) block was included. 20% of cases meeting inclusion criteria were randomly held out for an internal test set (*n* = 353). The remaining 80% was used for five-cross validation (*n* = 1,408 patients). This training dataset was enriched with dropped WSIs of FIGO stage IV cases or those with missing outcome such as the TCGA-UCEC cohort^21^ for training with self-supervised learning (*n* = 1,862). The external test set contains 151 patients with up to three FFPE blocks per case. More details for training and data split are in Methods. Altogether, including the two training steps and all downstream analyses, this comprehensive analysis comprised data of 2,590 tumors of women.

**Extended Data Fig. 2:**
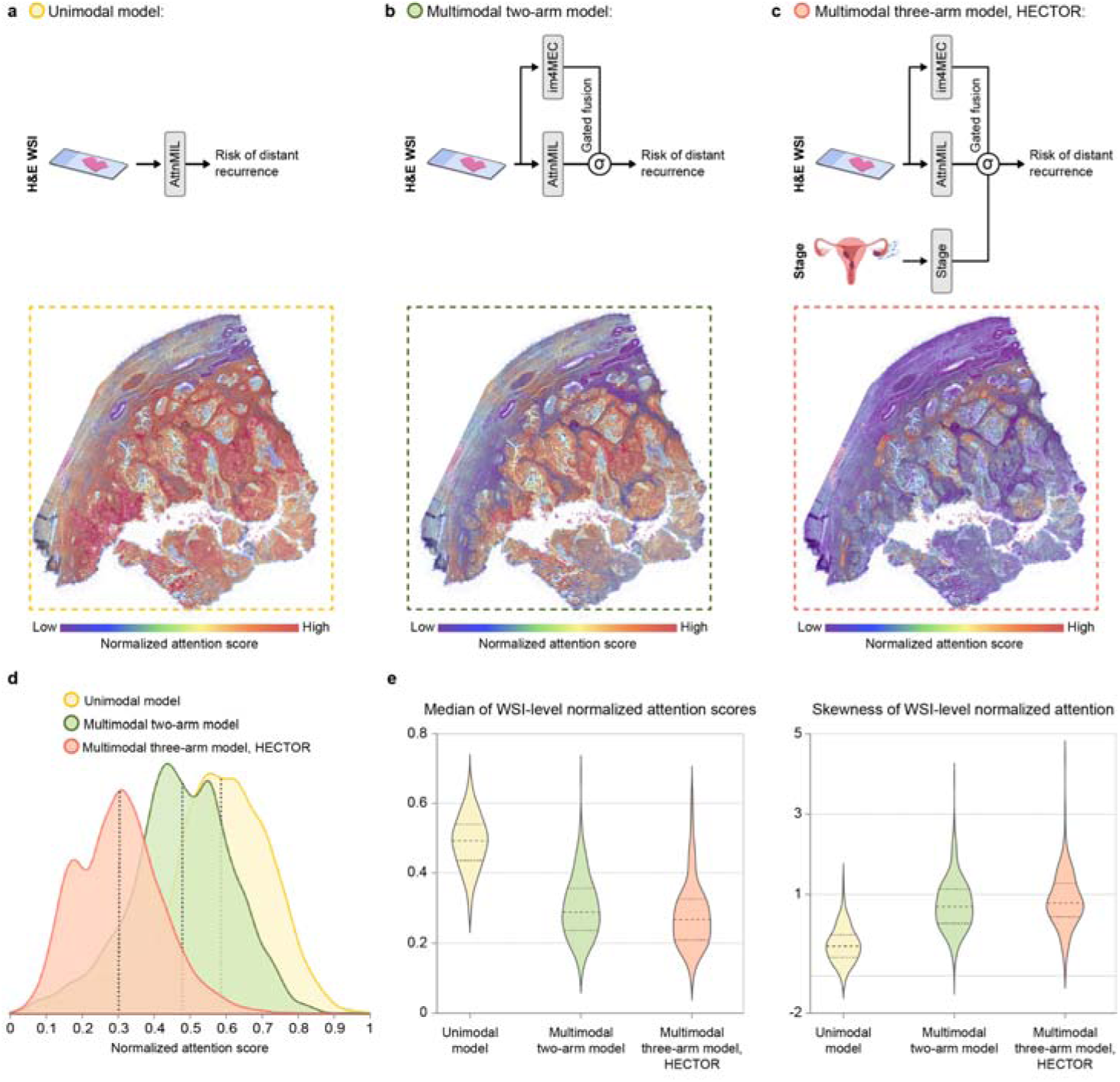
Shifts of attention scores from unimodal to multimodal model. **a**, Architectures using only H&E WSI (unimodal), two-arm with H&E WSI and image-based molecular class predicted by im4MEC, and the three-arm HECTOR model (multimodal) with H&E WSI, image-based molecular class, and stage. **b,** Two examples of the normalized attention scores shown as overlaid on the H&E whole slide image (WSI) as a heatmap where red is high attention score and blue low attention score, and as a density plot for the three models. The patch with the highest attention score is displayed. **c**, quantitative analysis of the distribution shift between the three models in the internal test set (*n* = 353 patients) using the WSI-level skewness and median of the normalized attention scores.

**Extended Data Fig. 3:**
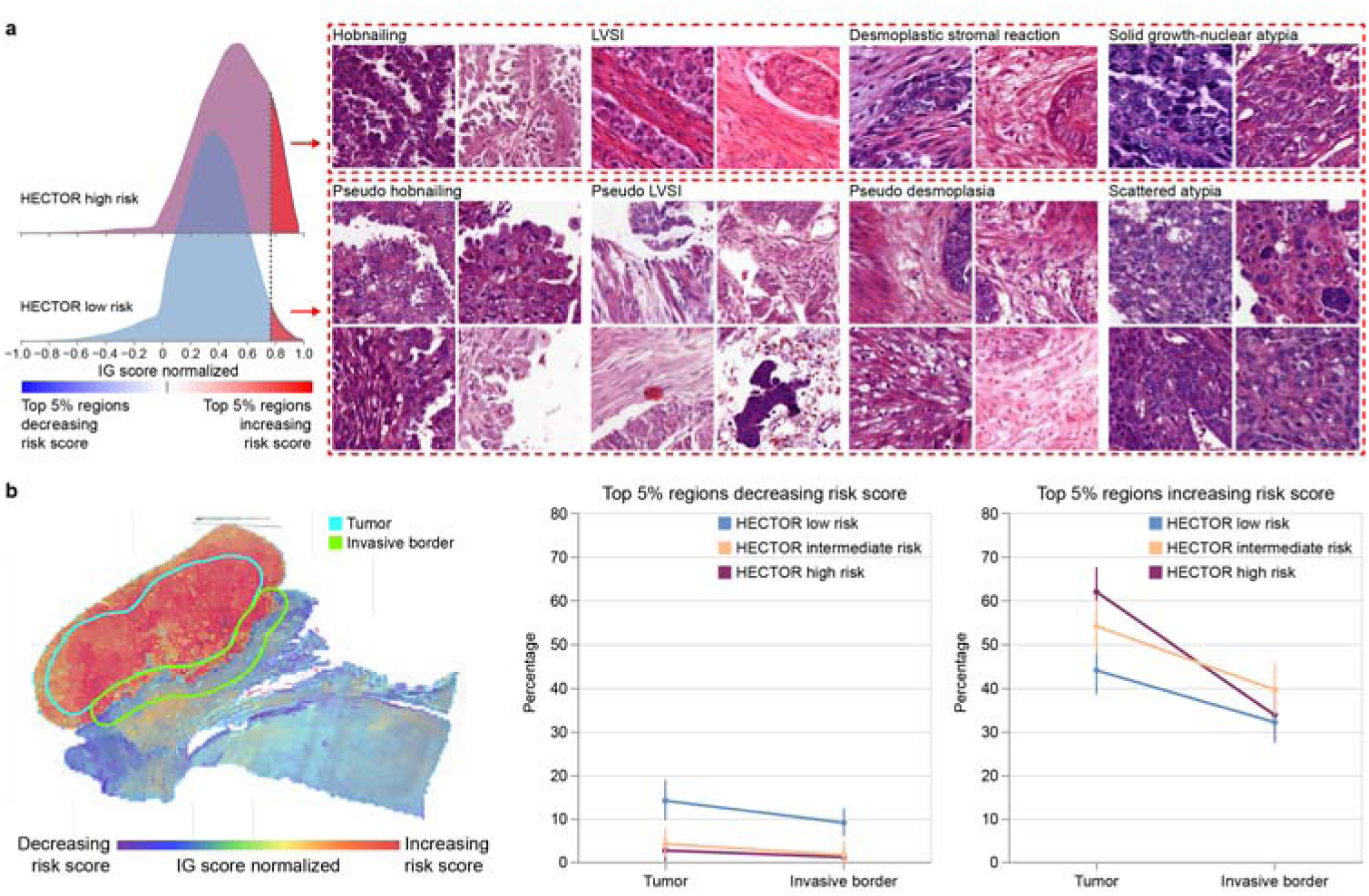
Morphological features increasing risk score in HECTOR high versus low risk group and quantitative spatial analysis. **a**, A representative selection of four patches for each morphological subtype (each selected from a different patient) increasing the risk score in the HECTOR low risk group as compared to the features increasing the risk score in the HECTOR high risk. **b,** Spatial analysis of top 5% regions decreasing and increasing the risk score based on the annotated areas: tumor and invasive border. (left) An example showing the annotation of the tumor area and invasive border of a whole slide image and heatmap showing the contribution of the regions using the Integrated Gradient (IG) analysis. (right) The relative contribution of these two annotated areas shown for each HECTOR risk group. IG = Integrated Gradient.

**Extended Data Fig. 4:**
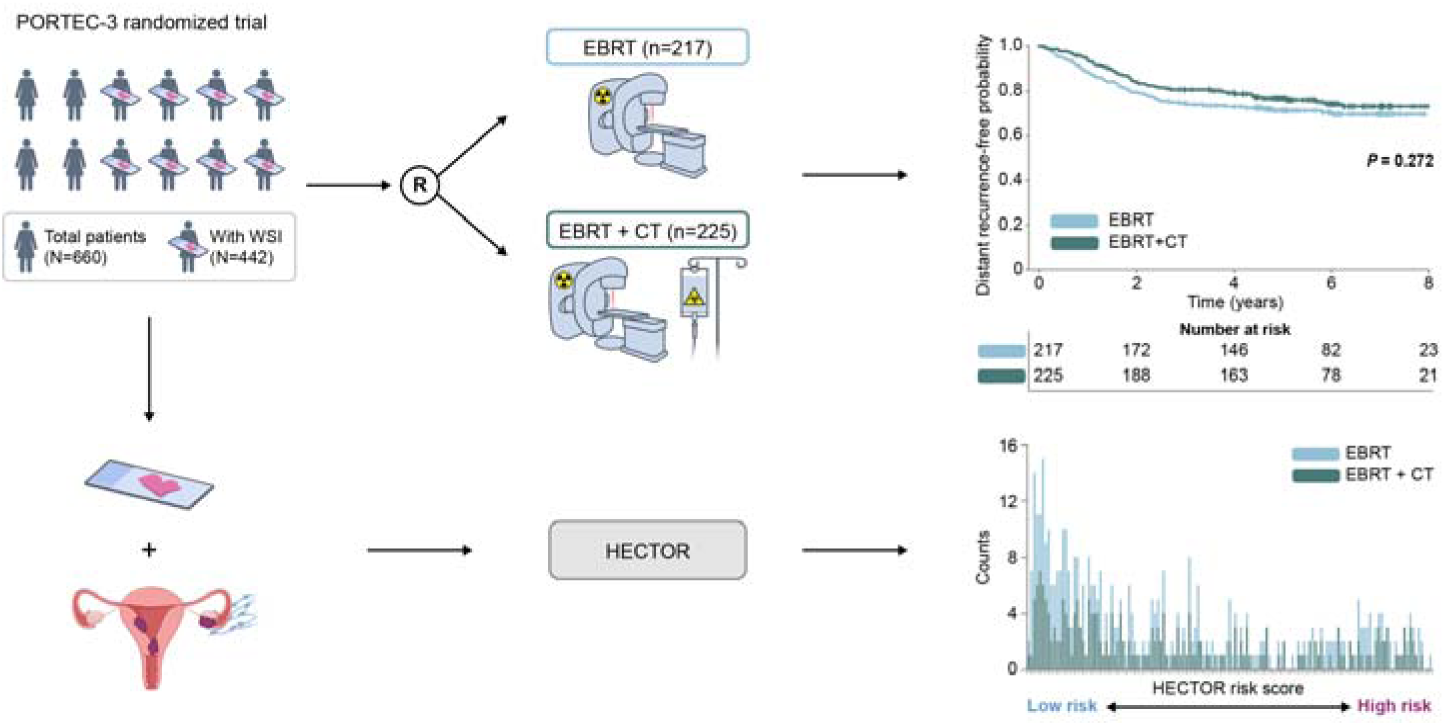
Overview of the PORTEC-3 randomized trial and analysis of treatment response prediction by HECTOR. In PORTEC-3^2^, 660 evaluable patients were randomized (1:1) between adjuvant external beam radiotherapy (EBRT) alone and external beam radiotherapy in combination with concurrent and adjuvant chemotherapy (CT). For 442 patients whose whole slide images (WSIs) were available, HECTOR risk scores were inferred. HECTOR risk groups cutoffs were kept the same as the training set (Methods).

## Online Methods

### Cohorts

We used formalin-fixed paraffin-embedded (FFPE) tumor material and clinicopathological data of patients with EC from three randomized trials and five clinical cohorts.

The PORTEC-1 trial recruited 714 women with early-stage intermediate risk EC from 1990 to 1997, and after primary surgery, randomly assigned to pelvic external beam radiotherapy or no adjuvant treatment^24^. The PORTEC-2 trial randomized 427 women with early-stage high-intermediate risk EC between 2000 to 2006 to external beam radiotherapy or vaginal brachytherapy^25^. The PORTEC-3 randomized trial included 660 women with stage I-III high risk EC from 2006 and 2013, and randomly allocated them to pelvic external beam radiotherapy alone or external beam radiotherapy combined with concurrent and adjuvant chemotherapy^2^. The retrospective TransPORTEC study included 116 high-risk EC tumors from international patients using the same inclusion criteria as the PORTEC-3 from five institutions (Leiden University Medical Center, The Netherlands; University Medical Center Groningen, The Netherlands; University College London, United Kingdom; St Mary’s Hospital, Manchester, United Kingdom; and Institute Gustave Roussy, Villejuif, France)^26^. The prospective cohort of Medisch Spectrum Twente (MST) included 257 patients with stage I-III high risk EC, with the same inclusion criteria as the PORTEC-3, who were treated between 1987 and 2015 at MST, Enschede in the Netherlands^27^. The Danish cohort consisted of 451 high-grade EC of patients who were prospectively registered in the Danish gynecological cancer database^28^. The Leiden cohort is a retrospectively collected population-based cohort of 222 patients diagnosed and treated at the Leiden University Medical Center between 2012 and 2021. Finally, the publicly available TCGA-UCEC cohort^29^ of 529 patients was downloaded from the cBioPortal^49,50^.

### Datasets

One representative H&E-stained slide of the hysterectomy specimen was included for each patient depending on the availability of the tumor material (Supplementary Fig. 1,2, Table 1,2,12). For the Leiden cohort, we collected three diagnostic H&E-stained tumor slides per patient case with EC, each from a different FFPE tumor tissue block. H&E slides were scanned at 40× magnification using two scanners 3DHistech P250 (resolution 0.19µm/pixel) and 3DHistech P1000 (resolution 0.24µm/pixel). Qualitative review was conducted on all WSIs, after which cases with no tumor, poor tissue quality and out-of-focus scanning issues were excluded, yielding 2,559 cases with at least one WSI per case.

In this study, some cases were excluded from the supervised training of HECTOR based on the following criteria: i) missing time to distant recurrence follow-up data^1^, ii) 2009 FIGO stage IV^31^ as they already have distant recurrence at time of diagnosis, iii) treatment with adjuvant chemotherapy as it may have lowered the risk of distant recurrence^2,3^. The categorical FIGO stage I, II, III is defined following the FIGO 2009 classification^31^. Distant recurrence was defined as any recurrence outside the pelvis. Hence, distant recurrence included abdominal metastasis and para-aortic lymph node metastasis. Time to distant recurrence was defined to start at randomization (for PORTEC-1, –2, and –3) or date of primary surgery (MST, HR pilot, Leiden cohort, Danish cohort), and to end at the date of the diagnosis of metastasis, or the date of last follow-up or death in patients without metastasis. We also stress that adjuvant chemotherapy was not the standard of care at the time the clinical cohorts were collected, and that the vast majority of patients treated with adjuvant chemotherapy originated from the PORTEC-3 randomized trial (*n* = 224).

Following the aforementioned criteria, 1,912 cases were included for the supervised train-test split: 584 from PORTEC-1^24^, 395 from PORTEC-2^25^, 217 PORTEC-3^2^, 67 from *Trans*PORTEC study^26^, 226 from MST cohort^27^, 272 from Danish cohort,^28^ and 151 from the Leiden cohort. Then we held out one internal test set and one external test set both representing an unselected population: the internal test set by randomly sampling 20% of supervised training set, stratified by discrete time intervals and censorship status (*n* = 353 of which 116 from PORTEC-1, 100 from PORTEC-2, 43 from PORTEC-3, 13 from *Trans*PORTEC study, 35 from MST cohort and 46 from Danish cohort; median follow-up of 8.45 years with 62 events) and the external test set that is the complete Leiden cohort (*n* = 151 patients: 121 with three WSIs, 21 with two WSIs and 9 with one WSI; 2.90-year median follow-up time with 24 events). The split of the internal test set was only stratified following the distribution of uncensored cases by discrete time intervals to ensure the presence of enough events across time in the internal test set. Finally, the remaining 1,408 WSIs were used for supervised training of HECTOR (468 from PORTEC-1, 295 from PORTEC-2, 174 from PORTEC-3, 54 from *Trans*PORTEC study, 191 from MST cohort and 226 from Danish cohort; median follow-up of 7.77 years with 246 events).

Additionally, the HECTOR risk scores were predicted on the previously excluded chemotherapy-treated cases from the PORTEC-3 randomized trial^2^ (*n* = 224) as well as the patients with stage I-III from TCGA-UCEC (*n* = 383).

For the self-supervised learning, we only used the 1,408 WSIs already reserved for supervised training, thus strictly limited to only those which were not part of the internal and external test sets. Additionally, the self-supervised learning training was enriched by cases with any stage of disease, whose treatment or distant recurrence outcome data was unknown (*n* = 454 of which 31 from *Trans*PORTEC study, 5 from MST cohort, 16 from Danish cohort, 402 from TCGA-UCEC), resulting in 1,862 cases for self-supervised learning.

### Performance evaluation

Hyperparameter optimization and model comparisons (including architecture choices for patch representation learning with self-supervised learning) were evaluated on the supervised downstream task guided by the C-index metric^30^. To this end, a five-fold cross-validation routine was performed on the 1,408 WSIs reserved for supervised training. The most performant architecture was selected based on the highest mean C-index. The cumulative area under the receiver operating curve^33^ and Brier scores^34^ were additionally computed.

Given the fact that the external test set contains up to three WSIs per case as opposed to one in the internal test set, we performed multiple experiments to derive patient-level risk scores using bootstrap sampling. First, we randomly selected one WSI per case and repeated it 100 times. Second, we randomly selected up to two WSIs for each case when available, we then averaged with the mean the two risk scores per patient, and repeated it 100 times. Third, we selected all available WSIs of the external test set with up to three WSIs per case when available, and computed the mean and median of the two or three risk scores. In an additional experiment, we combined each patient’s WSIs by merging the patch features from all available WSIs into a single feature bag.

### Whole slide image preprocessing

WSI segmentation was performed using Otsu thresholding and non-overlapping patching was performed at 180µm and patches were resized to 256×256 pixels. On average, this procedure generated a bag of 10,185 patches per WSI.

### Vision transformer-based patch representation learning

We followed advancements in self-supervised learning by adopting vision transformer-based deep learning models that are capable of learning fine-grained patch-level representation at multiple resolutions. For this, we trained EsVIT^45^ and compared it with CtransPath^51^, an alternative model trained on the histopathology domain (Supplementary Table 3). We modified the initial proposed four-stage Swin^52^ transformer-based architecture of EsVIT to capture cell– and region-level tissue information and to fit our computational resources. The patch size of stage-one was doubled to eight pixels to reduce the sequence length and increase field of view to capture cell views. In stages two to four, we kept the two-factor feature map merging rate, and resized the input images to 256×256 pixels instead of 224×224 pixels to avoid indivisible patch size at stage 4. Finally, the number of stacked transformers in stage three was reduced from six to four and the rest were kept to two. The first embedding dimension remained unchanged at 96 and the number of attention heads by stage was also kept unchanged, i.e. 3, 6, 12 and 24 (Supplementary Table 5).

A dataset of 3,702,447 patches was curated by randomly extracting up to 2,000 patches per WSI at 180µm resized to 256×256 pixels from the 1,862 WSIs appointed for self-supervised learning. Thereafter, the modified EsVIT was trained on three Nvidia RTX 8000 GPUs with a batch size of 128 for 100 epochs with a window of 14 to encourage learning of long term dependencies between patches. For performance improvement, we also used the view and region-level prediction DINO heads with no weight normalization and frozen layers at first epoch and the default output dimension of 65,536^45^. We followed the EsVIT authors’ recommendations with a smaller batch size by increasing the momentum teacher to 0.9996 and starting with the initial teacher temperature of 0.04. The teacher temperature was adjusted halfway through training from 0.04 to 0.02 for further loss decrease. We optimized with AdamW and default parameters, default optimization routines of the learning rate (linear warm up for ten epochs followed by cosine scheduler to 1×10^−6^), and weight decay (cosine scheduler from 0.04 to 0.4). The data augmentation was used exactly as done in the original publication^45^. Pytorch (version 1.8.1) was used.

After the training was completed, the patch-level features were extracted from the attention heads of the stacked transformers at each stage. For our downstream task, we observed an improvement by extracting the last eight blocks as compared to the default last four mentioned in the publication^45^, yielding feature vectors of size 3,456 (Supplementary Table 3).

### Multimodal deep learning prognostic model

To build the multimodal model for distant recurrence prediction task referred to as HECTOR, ablation studies were first performed using the H&E WSI modality only (referred to as H&E-based one-arm model) followed by integrating the FIGO stage and the image-based molecular classes derived from the H&E-based predictions of im4MEC^9^.

The H&E-based one-arm model took as input the bag of 180µm patch-level features of size 3,456 extracted from EsVIT^45^ where the number of patches per bag varies. We subsequently adapted the state-of-art WSI classification architectures^13,20^ for our distant recurrence prediction task. Given a batch size of one, the time scale was discretized into four intervals based on the quartiles of the distribution of uncensored patients and the negative log likelihood loss were used^46^. Our ablation study on H&E-based one-arm model showed that the attention-based multiple instance learning model (AttnMIL)^19^ outperformed spatial context-aware architectures including graph attention network^13^ and transformers^20^ while featuring far lower computational complexity (Supplementary Table 3).

Within the AttnMIL model, we reported a slight performance increase by adding another WSI preprocessing step. Specifically, WSI morphological information was spatially and semantically compressed by averaging highly correlated nearby patch-level features using a L2 norm threshold of three patches and a cosine similarity of 0.8. This step reduced the bag of features from 10,185 patches on average to 1,723 at 180µm (Supplementary Table 3). Each mean patch-level feature is compressed by three fully connected layers gradually down to 512. The attention module computes attention scores on latent features reduced to 256 before pooling, resulting in a slide-level embedding of size 512.

To fuse prognostic information of the categorical FIGO stage I-III variable and the image-based molecular classes derived from the H&E-based predictions of im4MEC^9^ to the H&E slide-level embedding (referred to as HECTOR), we proposed a novel approach of first encoding each categorical risk factor to higher-dimensional vector space with a learnable embedding of size 16 followed by Elu activation function and one fully connected layer of size 8. Next, a gating-based attention mechanism with bilinear product was applied on the embeddings from different modalities to weight the importance of each modality based on *Chen et al. 2022*^14^. To capture all interactions and retain unimodal embeddings, 1 was appended to the attention-weighted embeddings and then fused using the Kronecker product^32^. It is important to note that for using the image-based molecular class as an input modality for HECTOR, we re-trained the im4MEC model on the training set specifically designed for this study. This was done to avoid any information leakage as some cases used for training the original im4MEC model were used as testing on validation in this study.

The final multimodal embedding was further reduced by using two fully connected layers of size 256 and 128 before the survival categorical head of a fully connected layer with output size as the number of discrete time intervals. Each fully connected layer in the architecture was followed by a dropout of 0.25 and a ReLU activation function.

HECTOR was trained for 24 epochs with an initial learning rate of 3×10^−5^ decayed by a factor of 10 at epoch 2, 5, and 15. The Adam optimizer was used with default parameters and a weight decay of 1×10^−5^. Deep learning models were implemented with Pytorch (version 1.10.0).

### Association with clinicopathological data analysis

We performed multiple single linear regression analyses using the HECTOR continuous risk scores as the dependent variable and the clinicopathological data as the regressor. The statistical significance was accepted with *P* values below 0.050.

### Input contributions

The Integrated Gradients (IG) method^47^ was used to measure the contribution of the WSI and to identify the patches within a WSI relevant towards the prediction of the hazard function. Given the discrete time intervals, IG scores were averaged over the four neuron targets. The IG baseline for feature missingness was represented as patch-level features derived from white patches. All IG scores were patient-wise normalized between –1 and +1 while maintaining the sign and the IG score of zero, and further averaged to get a WSI-level IG score. Positive IG value towards 1 means contributed positively to increase the risk score, while negative means contributed to decrease the risk score. The Captum Python package (version 0.6.0) was used for implementation.

The contribution of the predicted image-based molecular class by im4MEC and the FIGO stage was calculated by fixing the stage and image-based molecular class values with the value of our choice (referred to as “reference group”) followed by computing the difference in predicted risk scores. Similar to the IG method, a positive or negative difference means a positive or negative contribution to the risk score respectively.

### Cell-level composition

As part of the explainability section of HECTOR to quantify visual features of extracted patches with high contribution, we first used the cell segmentation and classification Hover-Net^12^ deep learning model to obtain inflammatory cell counts, re-trained on EC-specific WSIs^9^. Then, mitotic figures were detected with a pan-cancer deep learning-based detector^48^ that was fine-tuned on EC tissue for the purpose of this study. Fine-tuning was performed by extending the original training set^53^ with additional data points we internally annotated in 10 WSIs from the PORTEC datasets that were selected to cover the variability of EC histological types. Region-level inflammatory and mitotic activity density were defined as absolute count normalized by the area in mm^2^; further averaged over the number of regions to obtain a patient-level density value. Statistical association between HECTOR risk scores and the quantity of visual features within the regions with either negative or positive contribution was tested with linear regression and *P* values significances accepted below 0.050.

### Outcome analysis

Analysis of distant recurrence-free probabilities was conducted according to Kaplan Meier’s method and the two-sided log-rank test with statistical significance was accepted with *P* below 0.050. Cutoffs for the HECTOR risk groups were defined by taking the quantiles (25%, 50%, 75%) of the distribution of HECTOR risk scores in the training set only. In the training set, the first two groups (below 25% and between 25% and 50%) did not show any major difference in prognosis and were therefore merged into one group named the HECTOR low risk group. As a result, we defined the HECTOR low risk group as cases with a risk score below the median risk score value of the training set, the HECTOR intermediate risk group as those with a risk score between median and third quartile values of the training set, and HECTOR high risk group as those with a risk score greater than the third quartile value of the training set. These same cutoff values were applied to the unseen internal and external test sets, and the TCGA-UCEC and PORTEC-3.

To compare the deep learning model performance to a Cox Proportional Hazard (CPH) model, we fitted CPH models on the well-established clinicopathological risk factors in EC. First using risk factors that can be visually assigned on histologic slides: the histological subtype, the grade and lympho-vascular space invasion. Then we added the FIGO stage I-III variable. Finally, we included the molecular class of endometrial cancer (*POLE*mut, MMRd, NSMP and p53abn). To keep consistent validation sets in the five-fold cross-validation and the internal test set, missing molecular class (115 out of 1,408 in cross-validation and 38 out of 353 in the internal test set) was imputed using mean substitution.

To estimate HECTOR’s prognostic value, we computed Hazard ratios using CPH with HECTOR continuous risk scores. For these analyses, we included all cases with a complete set of clinicopathological and molecular risk factors (*n* = 1,254). First, we corrected the HECTOR risk scores for all clinicopathological risk factors combined into one risk score in a multivariable analysis. To this end, a CPH model was first fitted onto these clinicopathological risk factors. Then risk scores were calculated by taking the linear combination of the CPH coefficients and the variables. In the second analysis, we corrected HECTOR’s continuous risk scores for the histological subtype, the grade, lympho-vascular space invasion, stage, the molecular class, and in addition, L1CAM and the age as continuous data. Statistical significance was accepted with *P* values below 0.050.

The histological subtype categorical variable was processed as Grade 3 endometrioid versus the reference group low-grade endometrioid and non-endometrioid versus the reference endometrioid. The reference group for molecular class was NSMP and stage I for the 2009 FIGO stage variable.

Cox Proportional Hazard models and Kaplan Meier’s method were implemented with the Lifelines Python package (version 0.27.1).

### Genomic and transcriptomic correlation analysis

To analyze the frequency of driver mutations by HECTOR risk groups, the genomic features were extracted from Thorsson et al.^54^ using MC3 MAF data. The mutational status of the top-20 oncogenic drivers in EC were downloaded from the cBioPortal portal^49,50^ and annotated by OncoKB^55^. The statistical comparison of proportions with oncogenic mutations between HECTOR risk groups was performed with the Chi square tests for each individual gene, with *P* values below 0.050 accepted as significant.

The association between the HECTOR continuous risk scores and each immune cell subset was performed using the log 2 transformed proportion of the immune cell subset as a fraction of the whole tumor, using the leukocyte fraction values. Linear regressions were performed with the HECTOR continuous risk scores as the independent variable. In addition, we tested the associations by correcting for the molecular class and tumor mutational burden (TMB) as additional independent variables.

mRNA-seq and clinical data from TCGA-UCEC were downloaded from firebrowse.org. Differentially expressed genes were assessed between HECTOR high risk and HECTOR low risk cases by DESeq2^56^ (version 1.40.1). Genes with a Benjamini–Hochberg FDR below 0.050 were selected as significantly different (Supplementary Table 13).

### Analysis of adjuvant chemotherapy effect on the risk of distant recurrence

We predicted the HECTOR risk scores onto the patients included in PORTEC-3^2^ treatment arm who did receive concurrent and adjuvant chemotherapy (*n* = 224) and, thus, who had been \previously left out from training and any test sets. The effect of the combination of adjuvant chemotherapy and external beam radiotherapy over external beam radiotherapy alone was analyzed by i) analyzing distant recurrence-free probabilities by treatment arm stratified by HECTOR risk group and measuring group-wise treatment effect with Kaplan Meier method the log-rank test; and/or HR of treatment variable with univariable Cox model, ii) calculating the statistical significance of the interaction term between the HECTOR continuous risk scores and the treatment binary variable, iii) calculating the statistical significance of the interaction term between the HECTOR high-risk group and the treatment binary variable (corrected for HECTOR intermediate risk group and using HECTOR low risk as reference group). To measure the statistical significance of the interaction term defined as the HECTOR risk score (continuous or categorical) multiplied by the treatment binary variable, a multivariable Cox regression analysis was performed. Similar analyses were performed to test the interaction between serous histological subtype and the chemotherapy treatment binary variable (corrected for Endometrioid and Clear Cells histological subtype), and of the FIGO 2009 stage III (corrected for stage I-II), and of p53abn (corrected for MMRd, NSMP as a reference group and *POLE*mut tumors removed to reach convergence).

Cox Proportional Hazard models and Kaplan Meier’s method implemented with the Lifelines Python package (version 0.27.1). The statistical significance was accepted with *P* values below 0.050.

## Data availability

The tumor material and datasets generated during or analyzed in this study are not publicly available due to restrictions by privacy laws. Data and tumor material from PORTEC-1, PORTEC-2, PORTEC-3, MST, the *Trans*PORTEC study, are held by the PORTEC study group and the international *Trans*PORTEC consortium. Data and tumor material from the Danish cohort are held by the coauthor of this article GO. Data and material from PORTEC-3 and the *Trans*PORTEC study are available to members of the international *Trans*PORTEC Research Consortium. Requests for sharing of all data and material should be addressed to the corresponding author within 15 years of the date of publication of this Article and include a scientific proposal. Depending on the specific research proposal, the *Trans*PORTEC consortium (PORTEC-3 and *Trans*PORTEC study) or the PORTEC study group (PORTEC-1, PORTEC-2, MST), or co-author G.Ø. will determine when, for how long, for which specific purposes, and under which conditions the requested data can be made available, subject to ethical consent. TCGA-UCEC data are publicly available via the cBioPortal^49,50^ for Cancer Genomics at https://www.cbioportal.org/study/clinicalData?id=ucec_tcga_pan_can_atlas_2018.

## Code availability

The code base is available at https://github.com/AIRMEC/HECTOR.

## Acknowledgements

This work was supported by a translational research project grant from the Hanarth Foundation and the Swiss Federal Institutes of Technology (strategic focus area of personalized health and related technologies; 2021–367) and a grant from the Promedica Foundation (F-87701–41–01) during the conduct of the study. The PORTEC-1, PORTEC-2, and PORTEC-3 trials were funded by grants from the Dutch Cancer Society (CKTO 90–01, CKTO 2001–04, and CKTO 2006–04, respectively). We first and foremost thank the women who participated in these studies and donated a tumor sample for translational research. We are grateful to the international and local investigators, and the data management teams who recruited and followed the women who participated in these studies, and to the many pathologists who collected samples for the PORTEC-1, PORTEC-2, and PORTEC-3 randomized trial biobanks, as well as the *Trans*PORTEC Research Consortium for the establishment of the *Trans*PORTEC study. We thank the investigators of the prospective MST cohort, and G.Ø. and E.H. and investigators of the Danish cohort. We are indebted to Tessa Rutten and Natalja ter Haar, Leiden University Medical Center, Leiden, the Netherlands, for excellent technical support, slide collection and scanning. We thank Lisa Vermij, Alicia Leon-Castillo, and Ellen Stelloo for the contribution to molecularly classifying the samples. We thank Viktoria Szabina Hadnagy, University Hospital Zurich, Zurich, Switzerland, for contributing to the annotation of the EC image dataset used to develop the mitosis detector used in this study. We also thank the Light Microscopy team of the Cell and Chemical Biology Department, Leiden University Medical center, for the technical support and use of the 3DHISTECH P250 scanner, and the Netherlands Cancer Institute for the use of their 3DHISTECH P1000 scanner. We acknowledge and thank the SHARK team, the computational cluster of the Leiden University Medical Center, for their technical support and the installation of the Nvidia RTX 8000 GPUs. We also thank Katie Yost for her work and support regarding the figure illustrations.

## Contributions

Conception and study design: S.FV., N.H., V.H.K., T.B.; model design and training: S.FV.; coding, implementation and technical support: S.FV., S.A, J.WB; acquisition of data: S.FV., N.H.; data analysis and interpretation: S.FV., N.H., S.A., J.WB., M.W.L., J.D., M.B., D.C., C.L.C., V.H.K., T.B.; drafting of the manuscript: S.F.; substantial revision of the manuscript: S.FV., N.H., S.A., J.WB., M.W.L., M.B., D.C., V.H.K., T.B.; All authors critically reviewed the manuscript, the results, and approved the final version.

## Footnotes

1 Stage category data was available for all included WSIs.

