## Supplementary data for "HECTOR: multimodal deep learning predicts recurrence risk in endometrial cancer"

### Supplementary content

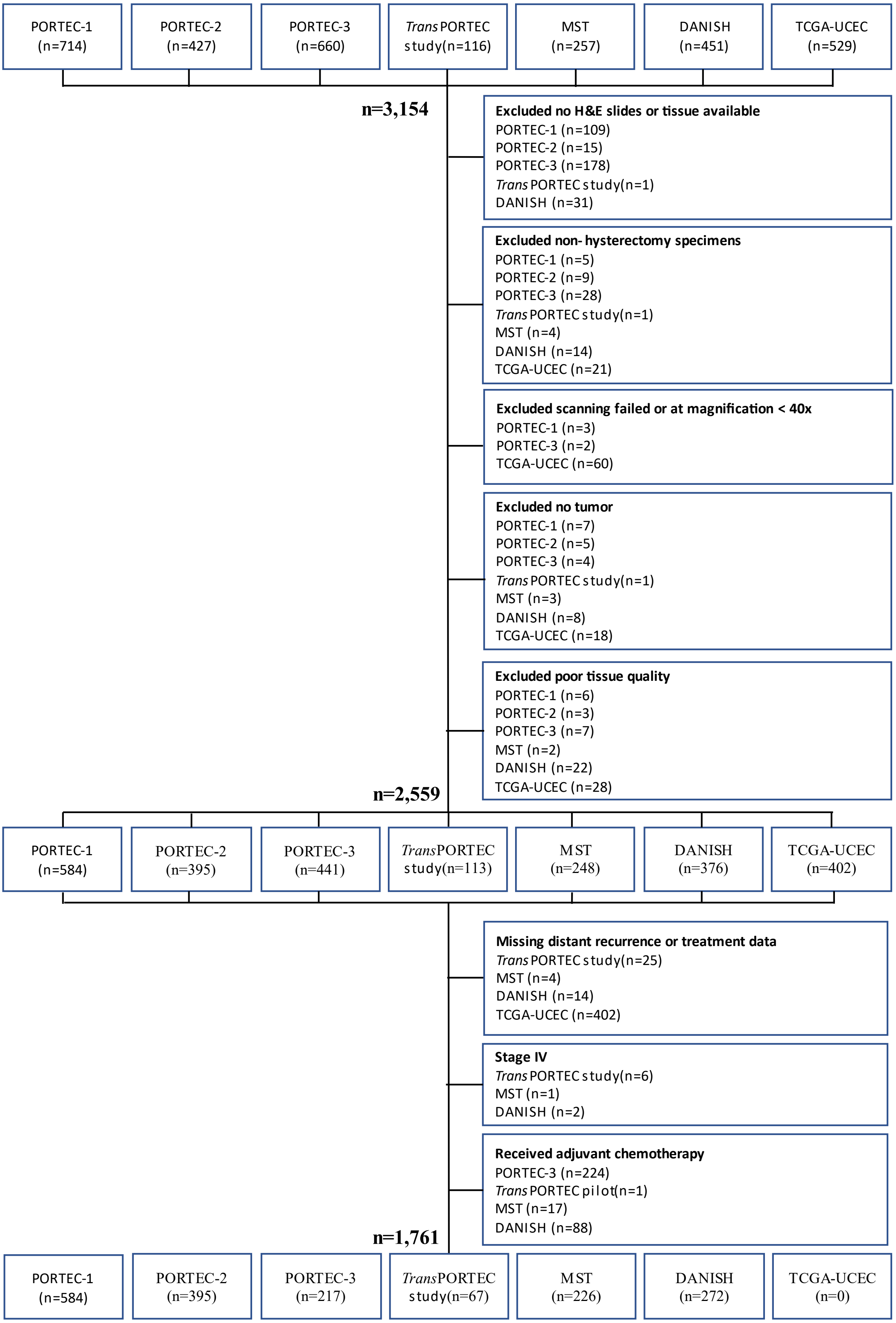


**Figure 1:** Flow chart showing exclusion of the cases. The exclusion of cases with no distant recurrence outcomes, 2009 FIGO Stage IV, or received adjuvant chemotherapy was only applied for supervised training of HECTOR.

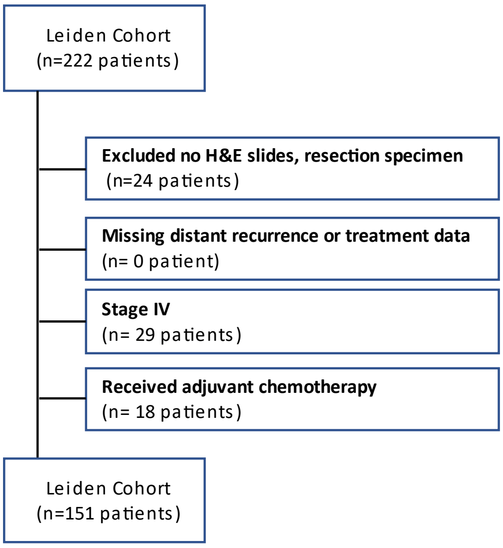


| | Inclusion | Leiden Cohort |
|---|---|---|
| **Number of patients (%)** | | **222** |
| | yes | 151 (68.0%) |
| | no | 71 (32.0%) |
| **Follow-up: median** | | |
| | yes | 2.90 |
| | no | 2.62 |
| **Age: median** | | |
| | yes | 66 |
| | no | 68 |
| **Histotype (%)** | | |
| Endometrioid grade 1-2 | yes | 88 (39.6%) |
| | no | 23 (10.4%) |
| Endometrioid grade 3 | yes | 22 (9.9%) |
| | no | 10 (4.5%) |
| Serous carcinoma | yes | 17 (7.7%) |
| | no | 18 (8.1%) |
| Clear cell carcinoma | yes | 12 (5.4%) |
| | no | 4 (1.8%) |
| Carcinosarcoma | yes | 3 (1.4%) |
| | no | 9 (4.1%) |
| Un-dedifferentiated | yes | 3 (1.4%) |
| | no | 3 (1.4%) |
| Other | yes | 6 (2.7%) |
| | no | 4 (1.8%) |
| **LVSI (%)** | | |
| Present | yes | 14 (6.3%) |
| | no | 11 (5.0%) |
| Focal or absent | yes | 127 (57.2%) |
| | no | 46 (20.7%) |
| Unknown | yes | 10 (4.5%) |
| | no | 14 (6.3%) |
| **2009 FIGO stage (%)** | | |
| IA | yes | 76 (34.2%) |
| | no | 19 (8.6%) |
| IB | yes | 42 (18.9%) |
| | no | 7 (3.2%) |
| II | yes | 14 (6.3%) |
| | no | 0 (0%) |
| IIIA | yes | 8 (3.6%) |
| | no | 6 (2.7%) |
| IIIB | yes | 1 (0.5%) |
| | no | 2 (0.9%) |
| IIIC | yes | 10 (4.5%) |
| | no | 7 (3.2%) |
| IV | yes | 0 (0%) |
| | no | 4 (1.8%) |
| Unkown | yes | 0 (0%) |
| | no | 26 (11.7%) |
| **Adjuvant treatment (%)** | | |
| None | yes | 79 (35.6%) |
| | no | 17 (7.7%) |
| VBT alone | yes | 36 (16.2%) |
| | no | 7 (3.2%) |
| EBRT alone | yes | 25 (11.3%) |
| | no | 6 (2.7%) |
| VBT + EBRT | yes | 11 (5.0%) |
| | no | 0 (0%) |
| Chemotherapy + EBRT | yes | 0 (0%) |
| | no | 15 (6.8%) |
| Chemotherapy alone | yes | 0 (0%) |
| | no | 25 (11.3%) |
| Unkown | yes | 0 (0%) |
| | no | 1 (0.5%) |

**Figure 2:** Flow chart (left) and characteristics of the Leiden Cohort (right). VBT = vaginal brachytherapy; EBRT = external beam radiotherapy.

| | | PORTEC-1 | PORTEC-2 | PORTEC-3 | *Trans* PORTEC study | MST | Danish |
|---|---|---|---|---|---|---|---|
| | **Inclusion** | | | | | | |
| **Number of patients (%)** | | **714** | **427** | **660** | **116** | **257** | **451** |
| | yes train | 468 (65.5%) | 295 (69.1%) | 174 (26.4%) | 54 (46.6%) | 191 (74.3%) | 226 (50.1%) |
| | yes test | 116 (16.2%) | 100 (23.4%) | 43 (6.5%) | 13 (11.2%) | 35 (13.6%) | 46 (10.2%) |
| | no | 130 (18.2%) | 32 (7.5%) | 443 (67.1%) | 49 (42.2%) | 31 (12.1%) | 179 (39.7%) |
| **Follow-up: median** | | | | | | | |
| | yes train | 10.54 | 9.49 | 4.99 | 3.30 | 5.92 | 6.57 |
| | yes test | 10.77 | 10.03 | 5.23 | 3.02 | 3.34 | 6.33 |
| | no | 9.79 | 9.58 | 5.05 | 1.36 | 4.94 | 5.84 |
| **Age: median** | | | | | | | |
| | yes train | 67 | 70 | 61 | 65 | 70 | 70 |
| | yes test | 67 | 69 | 63 | 71 | 65 | 70 |
| | no | 64 | 69 | 63 | 69 | 66 | 68 |
| **Histotype (%)** | | | | | | | |
| Endometrioid grade 1-2 | yes train | 389 (54.5%) | 261 (61.1%) | 73 (11.1%) | 13 (11.2%) | 77 (30.0%) | 0 (0%) |
| | yes test | 91 (12.7%) | 85 (19.9%) | 19 (2.9%) | 1 (0.9%) | 14 (5.4%) | 0 (0%) |
| | no | 121 (16.9%) | 28 (6.6%) | 165 (25.0%) | 4 (3.4%) | 10 (3.9%) | 0 (0%) |
| Endometrioid grade 3 | yes train | 68 (9.5%) | 28 (6.6%) | 54 (8.2%) | 39 (33.6%) | 52 (20.2%) | 111 (24.6%) |
| | yes test | 19 (2.7%) | 9 (2.1%) | 8 (1.2%) | 10 (8.6%) | 11 (4.3%) | 24 (5.3%) |
| | no | 8 (1.1%) | 4 (0.9%) | 123 (18.6%) | 19 (16.4%) | 6 (2.3%) | 57 (12.6%) |
| Serous carcinoma | yes train | 7 (1.0%) | 5 (1.2%) | 20 (3.0%) | 1 (0.9%) | 20 (7.8%) | 71 (15.7%) |
| | yes test | 1 (0.1%) | 6 (1.4%) | 8 (1.2%) | 2 (1.7%) | 2 (0.8%) | 14 (3.1%) |
| | no | 1 (0.1%) | 0 (0%) | 77 (11.7%) | 9 (7.8%) | 3 (1.2%) | 60 (13.3%) |
| Clear cell carcinoma | yes train | 2 (0.3%) | 0 (0%) | 15 (2.3%) | 1 (0.9%) | 8 (3.1%) | 18 (4.0%) |
| | yes test | 2 (0.3%) | 0 (0%) | 7 (1.1%) | 0 (0%) | 2 (0.8%) | 6 (1.3%) |
| | no | 0 (0%) | 0 (0%) | 40 (6.1%) | 17 (14.7%) | 4 (1.6%) | 12 (2.7%) |
| Carcinosarcoma | yes train | 0 (0%) | 0 (0%) | 0 (0%) | 0 (0%) | 20 (7.8%) | 8 (1.8%) |
| | yes test | 0 (0%) | 0 (0%) | 0 (0%) | 0 (0%) | 2 (0.8%) | 1 (0.2%) |
| | no | 0 (0%) | 0 (0%) | 0 (0%) | 0 (0%) | 2 (0.8%) | 4 (0.9%) |
| Un-dedifferentiated | yes train | 0 (0%) | 0 (0%) | 2 (0.3%) | 0 (0%) | 7 (2.7%) | 11 (2.4%) |
| | yes test | 0 (0%) | 0 (0%) | 1 (0.2%) | 0 (0%) | 1 (0.4%) | 1 (0.2%) |
| | no | 0 (0%) | 0 (0%) | 11 (1,7%) | 0 (0%) | 1 (0.4%) | 5 (1.1%) |
| Other | yes train | 2 (0.3%) | 1 (0.2%) | 10 (1.5%) | 0 (0%) | 7 (2.7% | 7 (1.6%) |
| | yes test | 3 (0.4%) | 0 (0%) | 0 (0%) | 0 (0%) | 3 (1.2%) | 0 (0%) |
| | no | 0 (0%) | 0 (0%) | 27 (4.1%) | 0 (0%) | 5 (1.9%) | 41 (9.1%) |
| **LVSI (%)** | | | | | | | |
| Present | yes train | 20 (2.8%) | 13 (3.0%) | 112 (17.0%) | 25 (21.6%) | 40 (15.6%) | 23 (5.1%) |
| | yes test | 4 (0.6%) | 6 (1.4%) | 23 (3.5%) | 6 (5.2%) | 7 (2.7%) | 1 (0.2%) |
| | no | 2 (0.3%) | 1 (0.2%) | 254 (38.5%) | 24 (20.7%) | 6 (2.3%) | 31 (6.9%) |
| Focal or absent | yes train | 410 (57.4%) | 269 (63.0%) | 62 (9.4%) | 28 (24.1%) | 151 (58.8%) | 202 (44.8%) |
| | yes test | 101 (14.1%) | 90 (21.1%) | 20 (3.0%) | 7 (6.0%) | 27 (10.5%) | 45 (10.0%) |
| | no | 26 (3.6%) | 14 (3.3%) | 189 (28.6%) | 5 (4.3%) | 24 (9.3%) | 107 (23.7%) |
| Unknown | yes train | 38 (5.3%) | 13 (3.0%) | 0 (0%) | 1 (0.9%) | 0 (0%) | 1 (0.2%) |
| | yes test | 11 (1.5%) | 4 (0.9%) | 0 (0%) | 0 (0%) | 1 (0.4%) | 0 (0%) |
| | no | 102 (14.3%) | 17 (4.0%) | 0 (0%) | 20 (17.2%) | 1 (0.4%) | 41 (9.1%) |

**Table 1**: Characteristics (such as median follow-up, age, histoype and LVSI) of the cohorts used in this study stratified by whether patients were included in the supervised training, supervised testing or excluded. Exclusion criteria included lack of material (H&E slide), missing distant recurrence data, 2009 FIGO stage IV and patients who received adjuvant chemotherapy. LVSI = lymphovascular space invasion.

| | Inclusion | PORTEC-1 | PORTEC-2 | PORTEC-3 | *Trans* PORTEC study | MST | Danish |
|---|---|---|---|---|---|---|---|
| **2009 FIGO stage (%)** | | | | | | | |
| IA | yes train | 190 (26.6%) | 47 (11.0%) | 21 (3.2%) | 3(2.6%) | 20 (7.8%) | 126 (27.9%) |
| | yes test | 47 (6.6%) | 16 (3.7%) | 5 (0.8%) | 1 (0.9%) | 2 (0.8%) | 24 (5.3%) |
| | no | 57 (8.0%) | 8 (1.9%) | 52 (7.9%) | 7 (6.0%) | 3 (1.2%) | 24 (5.3%) |
| IB | yes train | 278 (38.9%) | 244 (57.1%) | 32 (4.8%) | 20 (17.2%) | 57 (22.2%) | 66 (14.6%) |
| | yes test | 69 (9.7%) | 83 (19.4%) | 6 (0.9%) | 4 (3.4%) | 9 (3.5%) | 12 (2.7%) |
| | no | 73 (10.2%) | 24 (5.6%) | 79 (12.0%) | 7 (6.0%) | 2 (0.8%) | 18 (4.0%) |
| II | yes train | 0 (0%) | 2 (0.5%) | 50 (7.6%) | 13 (11.2%) | 62 (24.1%) | 17 (3.8%) |
| | yes test | 0 (0%) | 0 (0%) | 10 (1.5%) | 4 (3.4%) | 7 (2.7%) | 3 (0.7%) |
| | no | 0 (0%) | 0 (0%) | 110 (16.7%) | 4 (3.4%) | 10 (3.9%) | 9 (2.0%) |
| IIIA | yes train | 0 (0%) | 2 (0.5%) | 18 (2.7%) | 9 (7.8%) | 33 (12.8%) | 1 (0.2%) |
| | yes test | 0 (0%) | 0 (0%) | 6 (0.9%) | 2 (1.7%) | 11 (4.3%) | 2 (0.4%) |
| | no | 0 (0%) | 0 (0%) | 59 (8.9%) | 7 (6.0%) | 6 (2.3%) | 3 (0.7%) |
| IIIB | yes train | 0 (0%) | 0 (0%) | 16 (2.4%) | 1 (0.9%) | 8 (3.1%) | 5 (1.1%) |
| | yes test | 0 (0%) | 1 (0.2%) | 1 (0.2%) | 0 (0%) | 4 (1.6%) | 3 (0.7%) |
| | no | 0 (0%) | 0 (0%) | 25 (3.8%) | 3 (2.6%) | 6 (2.3%) | 6 (1.3%) |
| IIIC | yes train | 0 (0%) | 0 (0%) | 37 (5.6% | 8 (6.9%) | 11 (4.3%) | 11 (2.4%) |
| | yes test | 0 (0%) | 0 (0%) | 15 (2.3%) | 2 (1.7%) | 2 (0.8%) | 2 (0.4%) |
| | no | 0 (0%) | 0 (0%) | 118 (17.9%) | 9 (7.8%) | 2 (0.8%) | 48 (10.6%) |
| IV | yes train | 0 (0%) | 0 (0%) | 0 (0%) | 0 (0%) | 0 (0%) | 0 (0%) |
| | yes test | 0 (0%) | 0 (0%) | 0 (0%) | 0 (0%) | 0 (0%) | 0 (0%) |
| | no | 0 (0%) | 0 (0%) | 0 (0%) | 11 (9.5%) | 2 (0.8%) | 32 (7.1%) |
| Unkown | yes train | 0 (0%) | 0 (0%) | 0 (0%) | 0 (0%) | 0 (0%) | 0 (0%) |
| | yes test | 0 (0%) | 0 (0%) | 0 (0%) | 0 (0%) | 0 (0%) | 0 (0%) |
| | no | 0 (0%) | 0 (0%) | 0 (0%) | 1 (0.9%) | 0 (0%) | 0 (0%) |
| **Molecular class (%)** | | | | | | | |
| *POLE* mut | yes train | 31 (4.3%) | 17 (4.0%) | 21 (3.2%) | 12 (10.3%) | 12 (4.7%) | 29 (6.4%) |
| | yes test | 9 (1.3%) | 7 (1.6%) | 6 (0.9%) | 2 (1.7%) | 2 (0.8%) | 2 (0.4%) |
| | no | 2 (0.3%) | 0 (0%) | 24 (3.6%) | 2 (1.7%) | 2 (0.8%) | 7 (1.6%) |
| MMRd | yes train | 111 (15.5%) | 85 (19.9%) | 59 (8.9%) | 10 (8.6% | 58 (22.6%) | 61 (13.5%) |
| | yes test | 22 (3.1%) | 20 (4.7%) | 11 (1.7%) | 1 (0.9%) | 7 (2.7%) | 14 (3.1%) |
| | no | 4 (0.6%) | 5 (1.2%) | 69 (10.5%) | 9 (7.8%) | 6 (2.3%) | 35 (7.8%) |
| NSMP | yes train | 209 (29.3%) | 162 (37.9%) | 46 (7.0%) | 21 (18.1%) | 66 (25.7%) | 36 (8.0%) |
| | yes test | 51 (7.1%) | 64 (15.0%) | 11 (1.7%) | 7 (6.0%) | 16 (6.2%) | 10 (2.2%) |
| | no | 5 (0.7%) | 6 (1.4%) | 65 (9.8%) | 15 (12.9%) | 8 (3.1%) | 28 (6.2%) |
| p53abn | yes train | 30 (4.2%) | 24 (5.6%) | 32 (4.8%) | 10 (8.6%) | 51 (19.8%) | 100 (22.2%) |
| | yes test | 9 (1.3%) | 4 (0.9%) | 8 (1.2%) | 3 (2.6%) | 9 (3.5%) | 20 (4.4%) |
| | no | 1 (0.1%) | 2 (0.5%) | 59 (8.9%) | 21 (18.1%) | 9 (3.5%) | 70 (15.5%) |
| Unkown | yes train | 87 (12.2%) | 7 (1.6%) | 16 (2.4%) | 1 (0.9%) | 4 (1.6%) | 0 (0%) |
| | yes test | 25 (3.5%) | 5 (1.2%) | 7 (1.1%) | 0 (0%) | 1 (0.4%) | 0 (0%) |
| | no | 118 (16.5%) | 19 (4.4%) | 226 (34.2%) | 2 (1.7%) | 6 (2.3%) | 39 (8.6%) |
| **Adjuvant treatment (%)** | | | | | | | |
| None | yes train | 241 (33.8%) | 1 (0.2%) | 0 (0%) | 3 (2.6%) | 0 (0%) | 198 (43.9%) |
| | yes test | 63 (8.8%) | 0 (0%) | 0 (0%) | 1 (0.9%) | 0 (0%) | 43 (9.5%) |
| | no | 65 (9.1%) | 2 (0.5%) | 0 (0%) | 6 (5.2%) | 0 (0%) | 30 (6.7%) |
| VBT alone | yes train | 0 (0%) | 148 (34.7%) | 0 (0%) | 1 (0.9%) | 17 (6.6%) | 0 (0%) |
| | yes test | 0 (0%) | 50 (11.7%) | 0 (0%) | 0 (0%) | 2 (0.8%) | 0 (0%) |
| | no | 0 (0%) | 17 (4.0%) | 0 (0%) | 0 (0%) | 3 (1.2%) | 0 (0%) |
| EBRT alone | yes train | 227 (31.8%) | 146 (34.2%) | 174 (26.4%) | 42 (36.2%) | 174 (67.7%) | 28 (6.2%) |
| | yes test | 53 (7.4%) | 50 (11.7%) | 43 (6.5%) | 8 (6.9%) | 33 (12.8%) | 3 (0.7%) |
| | no | 65 (9.1%) | 13 (3.0%) | 116 (17.6%) | 2 (1.7%) | 7 (2.7%) | 4 (0.9%) |
| EBRT + VBT | yes train | 0 (0%) | 0 (0%) | 0 (0%) | 8 (6.9%) | 0 (0%) | 0 (0%) |
| | yes test | 0 (0%) | 0 (0%) | 0 (0%) | 4 (3.4%) | 0 (0%) | 0 (0%) |
| | no | 0 (0%) | 0 (0%) | 0 (0%) | 1 (0.9%) | 0 (0%) | 0 (0%) |
| Chemotherapy + EBRT | yes train | 0 (0%) | 0 (0%) | 0 (0%) | 0 (0%) | 0 (0%) | 0 (0%) |
| | yes test | 0 (0%) | 0 (0%) | 0 (0%) | 0 (0%) | 0 (0%) | 0 (0%) |
| | no | 0 (0%) | 0 (0%) | 327 (49.5%) | 11 (9.5%) | 15 (5.8%) | 8 (1.8%) |
| chemotherapy alone | yes train | 0 (0%) | 0 (0%) | 0 (0%) | 0 (0%) | 0 (0%) | 0 (0%) |
| | yes test | 0 (0%) | 0 (0%) | 0 (0%) | 0 (0%) | 0 (0%) | 0 (0%) |
| | no | 0 (0%) | 0 (0%) | 0 (0%) | 5 (4.3%) | 0 (0%) | 98 (21.7%) |
| Unkown | yes train | 0 (0%) | 0 (0%) | 0 (0%) | 0 (0%) | 0 (0%) | 0 (0%) |
| | yes test | 0 (0%) | 0 (0%) | 0 (0%) | 0 (0%) | 0 (0%) | 0 (0%) |
| | no | 0 (0%) | 0 (0%) | 0 (0%) | 24 (20.7%) | 6 (2.3%) | 39 (8.6%) |

**Table 2**: Characteristics (such as 2009 FIGO stage, Molecular class, Adjuvant treatment) of the cohorts used in this study stratified by whether patients were included in the supervised training, supervised testing or excluded. Exclusion criteria included lack of material (H&E slide), missing distant recurrence

data, 2009 FIGO stage IV and patients who received adjuvant chemotherapy. *POLE*mut = *POLE* mutant; MMRd = mismatch repair deficient; NSMP = No specific molecular profile; p53abn = p53 abnormal; VBT = vaginal brachytherapy; EBRT = external beam radiotherapy.

| Self-supervised features | C-index⇧ | Cumulative AUC⇧ | IBS⇩ |
|---|---|---|---|
| Modified EsVIT (8 blocks) | 0.770±0.0193 | 0.823±0.0353 | 0.114±0.0118 |
| Modified EsVIT (4 blocks) | 0.742±0.0294 | 0.777±0.0503 | 0.105±0.0056 |
| CtransPath | 0.728±0.0332 | 0.765±0.0650 | 0.100±0.0021 |
| Unimodal model (H&E only) | | | |
| AttentionMIL + merged EsVIT features | 0.775±0.0312 | 0.810±0.0257 | 0.124±0.0057 |
| AttentionMIL + EsVIT features | 0.770±0.0193 | 0.823±0.0353 | 0.114±0.0118 |
| GAT + merged EsVIT features | 0.766±0.0109 | 0.804±0.0441 | 0.0986±0.0069 |
| Transformer + merged EsVIT features | 0.762±0.0201 | 0.801±0.0611 | 0.109±0.0101 |
| Multimodal model (in addition to merged EsVIT features) | | | |
| AttentionMIL (unimodal baseline) | 0.775±0.0312 | 0.810±0.0257 | 0.124±0.0057 |
| AttentionMIL + fusion with image embeddings produced by im4MEC | 0.764±0.0151 | 0.806±0.0371 | 0.111±0.0102 |
| AttentionMIL + fusion with Embedding layer for im4MEC classes | 0.779±0201 | 0.820±0.0354 | 0.110±0.0281 |
| AttentionMIL + fusion with attention-gated Embedding layer for im4MEC classes | 0.782±0.0264 | 0.824±0.0402 | 0.113±0.0094 |
| AttentionMIL + fusion with categorical 2009 FIGO stage IA-IB-II-IIIA-IIIB-IIIC | 0.767±0.0199 | 0.822±0.0367 | 0.104±0.0077 |
| AttentionMIL + fusion with categorical 2009 FIGO stage I-II-III | 0.772±0.0160 | 0.816±0.0245 | 0.101±0.0081 |
| AttentionMIL + fusion with Embedding layer for 2009 FIGO stage I-II-III | 0.786±0.0081 | 0.841±0.0424 | 0.106±0.0243 |
| AttentionMIL + fusion with attention-gated Embedding layer for 2009 FIGO stage I-II-III | 0.790±0.0089 | 0.830±0.0320 | 0.102±0.0065 |
| **AttentionMIL + 2-sided attention-gated Embedding layer for im4MEC classes and categorical 2009 FIGO stage I-II-III (HECTOR)** | **0.795±0.0307** | **0.844±0.0290** | **0.097±0.0122** |

**Table 3:** Ablation studies on five-fold cross-validation (*n* = 1,408 patients). (Top) Comparison of EsVIT[1] (a state-of-the-art transformer-based model trained with self-supervised learning, re-trained on endometrial cancer-specific patches, where patch-level embeddings are extracted from either the last four or height transformer blocks) and CtransPath[2] (trained on histology images). Performance was measured on the downstream supervised task using unimodal (H&E only) AttentionMIL[3], that is the prediction of distant recurrence-free probabilities. Modified EsVIT refers to the architecture described in Table 3. (Middle) Comparison of state-of-the-art supervised deep learning architectures for distant recurrence-free probabilities prediction using only H&E WSIs as input and EsVIT features from the last height blocks. All models were trained with the negative log likelihood loss[4] and a batch size of 1 for fair comparison. For GAT[5], we experimented a radius of 16 and 32 patches. Merged features extracted from EsVIT were obtained by averaging patches spatially and semantically merging using L2 norm of three patches and a cosine similarity above 0.8. Transformers do not include patch positional encoding[6]. (Bottom) Ablation studies with multimodal deep learning. We experimented using either the WSI-level embeddings produced by im4MEC or the categorical image-based molecular class as predicted by im4MEC[7]. We used the 2009 FIGO stage classification[8]. Mean and standard deviation

across validation folds of C-index[9], cumulative AUC[10], IBS[11] are reported. Final hyperparameter selection was performed based on the highest mean C-index. HECTOR, which returned the highest mean C-index, was selected for downstream analysis. C-index=concordance index; AUC = area under the receiver operating characteristic curve; IBS = integrated Brier score; GAT = graph attention networks; AttentionMIL = attention multiple instance learning.

| Input required | Model | C-index⇧ | Cumulative AUC⇧ | IBS⇩ |
|---|---|---|---|---|
| H&E tumor slide | Unimodal DL | 0.775±0.0312 | 0.810±0.0257 | 0.124±0.0057 |
| | Two-arm DL | 0.782±0.0264 | 0.824±0.0402 | 0.113±0.0094 |
| | CPH | 0.676±0.0601 | 0.721±0.0742 | 0.129±0.1023 |
| H&E tumor slide + 2009 FIGO stage I-III | HECTOR | **0.795±0.0307** | **0.844±0.0290** | **0.097±0.0122** |
| | CPH | 0.723±0.0449 | 0.762±0.0539 | 0.128±0.0443 |
| H&E tumor slide + 2009 FIGO stage I-III + true molecular class | CPH | 0.736±0.0328 | 0.809±0.0472 | 0.128±0.0029 |

**Table 4:** Comparison of unimodal and multimodal deep learning models and baseline Cox proportional hazard (CPH) for distant recurrence-free probabilities prediction in the five-fold cross validation (n = 1,408 patients). We report the cumulative AUC[10] and IBS[11]. H&E-based variables used in CPH are histologic subtype, grade, and lymphovascular space invasion. 2009 FIGO I-III classification was used[8]. Unlike the C-index and Cumulative AUC, a lower IBS value is better. H&E = Hematoxylin and eosin; C-index=concordance index; cumulative AUC = cumulative area under the receiver operating characteristic curve; IBS= integrated brier score.

| Modified Swin, W=14, S=256 | Stage 1 | Stage 2 | Stage 3 | Stage 4 |
|---|---|---|---|---|
| Merging rate<br>Feature map | 8x<br>32 x 32 | 16x<br>16 x 16 | 32x<br>8 x 8 | 64x<br>4 x 4 |
| Architecture | $\begin{bmatrix}\text{window size: } 14 \times 14 \\ 96\text{-}d\ (3\ heads)\end{bmatrix} \times 2$ | $\begin{bmatrix}\text{window size: } 14 \times 14 \\ 192\text{-}d\ (6\ heads)\end{bmatrix} \times 2$ | $\begin{bmatrix}\text{window size: } 14 \times 14 \\ 384\text{-}d\ (12\ heads)\end{bmatrix} \times 4$ | $\begin{bmatrix}\text{window size: } 14 \times 14 \\ 768\text{-}d\ (24\ heads)\end{bmatrix} \times 2$ |

**Table 5:** Architecture of the modified four-stage EsVIT used[1]. W = window size; S = Size of input image in pixels.

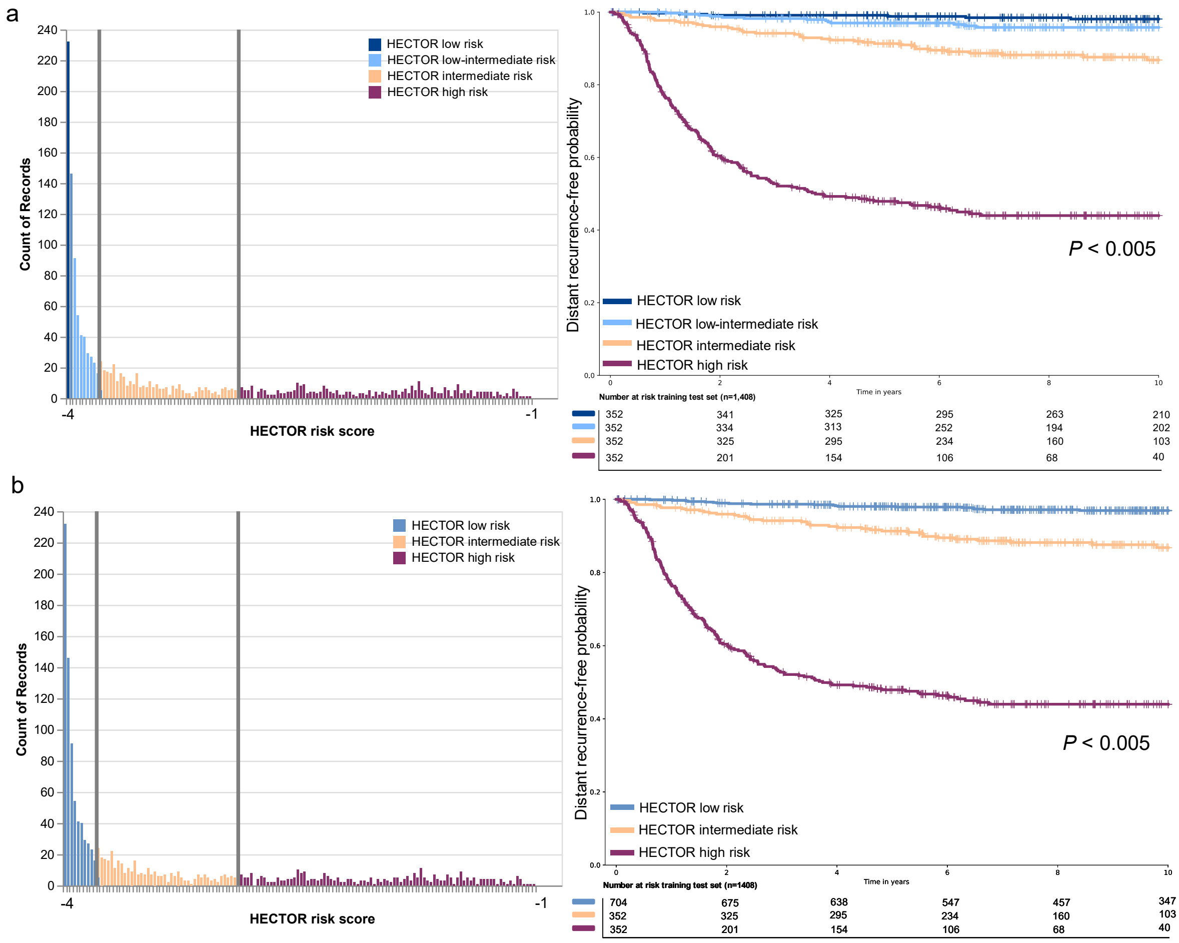


**Figure 3:** Categorization of the HECTOR continuous risk scores based on the quartiles of the distribution in the training set ($n$ = 1,408 patients) and the corresponding 10-year distant recurrence-free probabilities in the training set. a, The quartiles are used to create four groups low, low-intermediate, intermediate and high risk. b, Patients from low and low-intermediate were combined into the HECTOR low risk group.

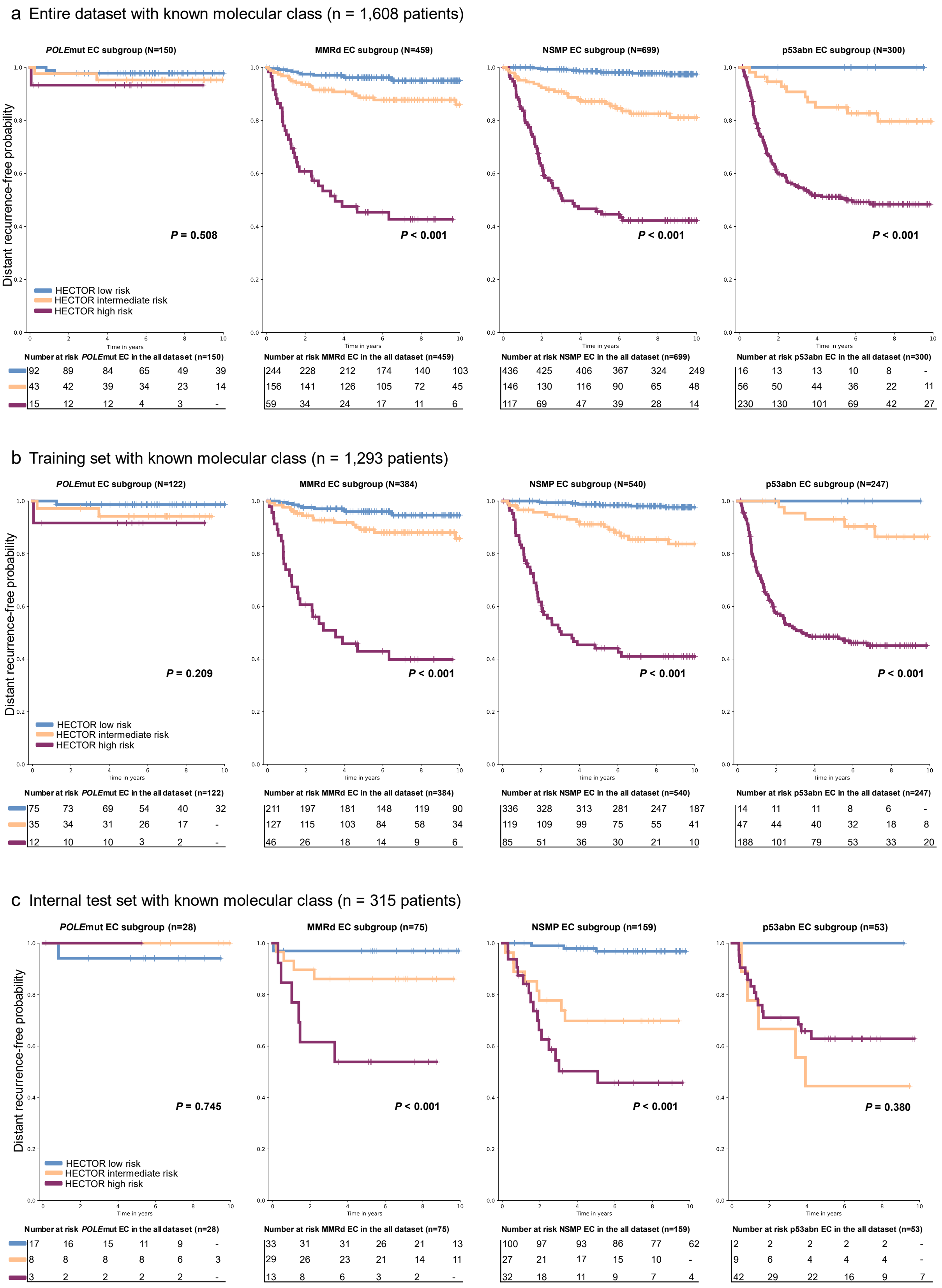


**Figure 4:** Distant recurrence-free probability analysis using Kaplan-Meier method within each molecular class of endometrial cancer stratified by HECTOR risk groups. MMRd= mismatch repair deficient; p53abn = p53 abnormal; NSMP = no specific molecular profile.

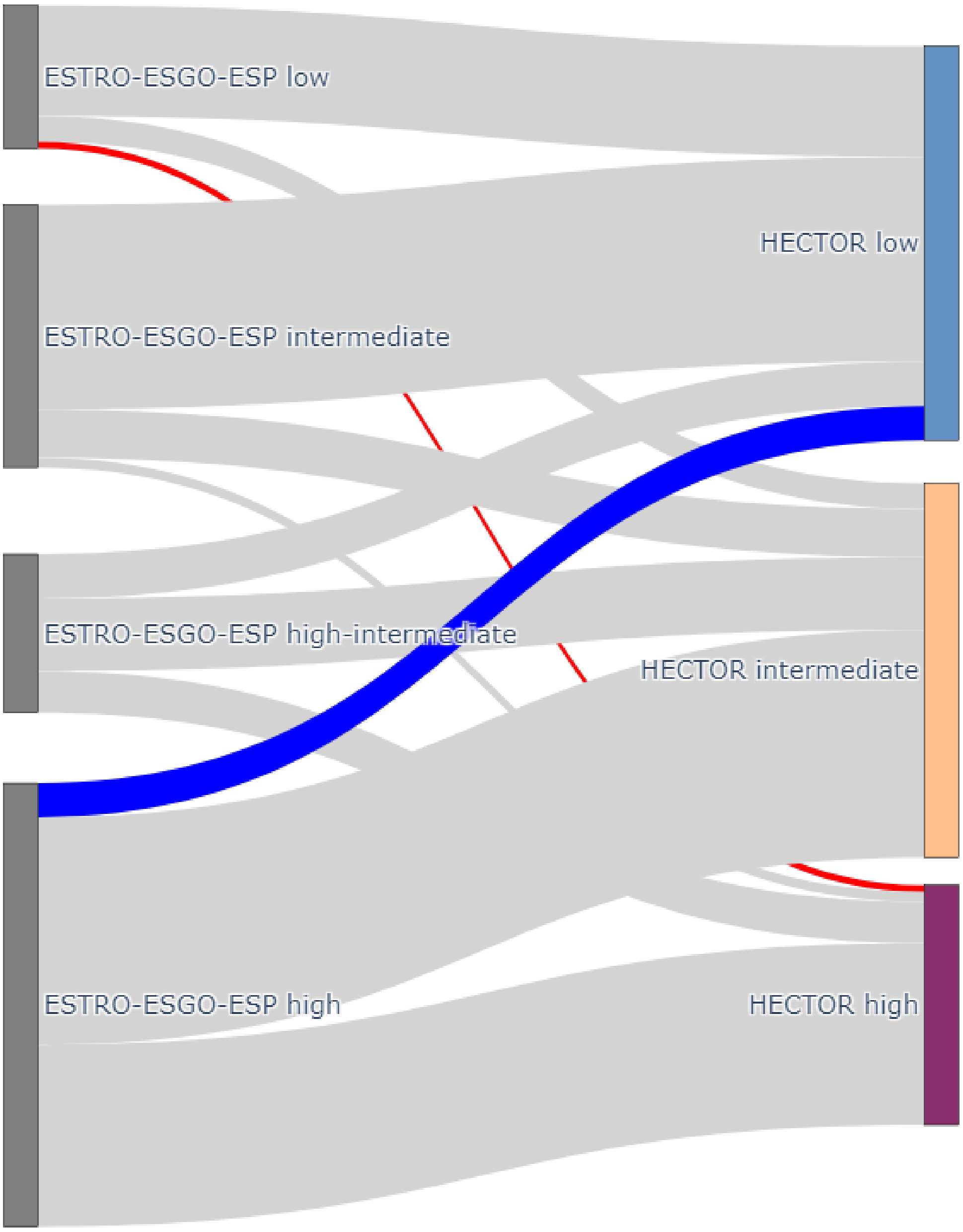


**Figure 5:** Shift between the risk assessment of the ESGO-ESTRO-ESP 2021 guidelines[12] and HECTOR risk group in the entire dataset ($n$ = 1,758 patients) with all known prognostic factors (such as LVSI and molecular class).

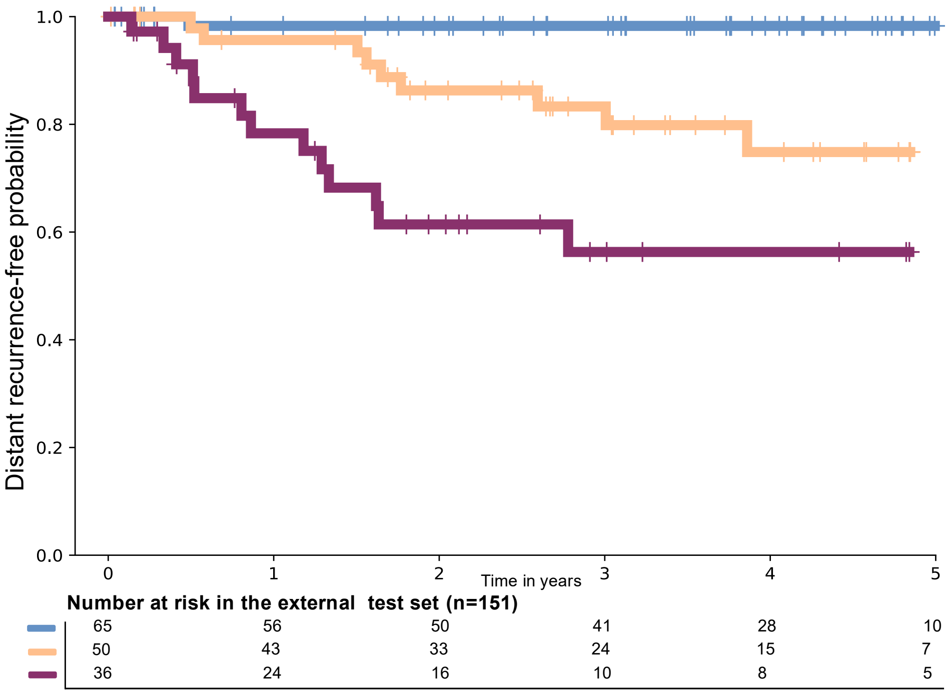


**Figure 6:** Five-year distant recurrence-free probabilities in the external test set (*n* = 151 patients) by post-aggregation with mean, i.e. taking the mean of the predicted HECTOR risk scores obtained for all the WSIs of each patient.

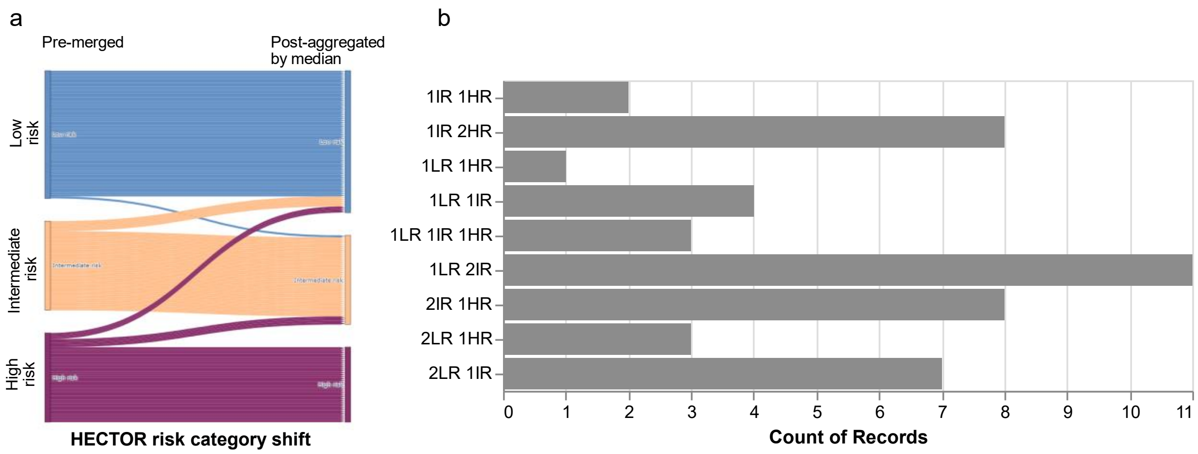


**Figure 7:** a, Sankey plot showing patient-level shifts between a scenario where the whole slide images are pre-merged into one bag of features (left) yielding a C-index of 0.805 and alternatively when they were post-aggregated by median (right) yielding a c-index of 0.816. b, Number of cases with more than one WSI with a different predicted risk category from HECTOR. LR = HECTOR low risk; IR = HECTOR intermediate risk; HR = HECTOR high risk.

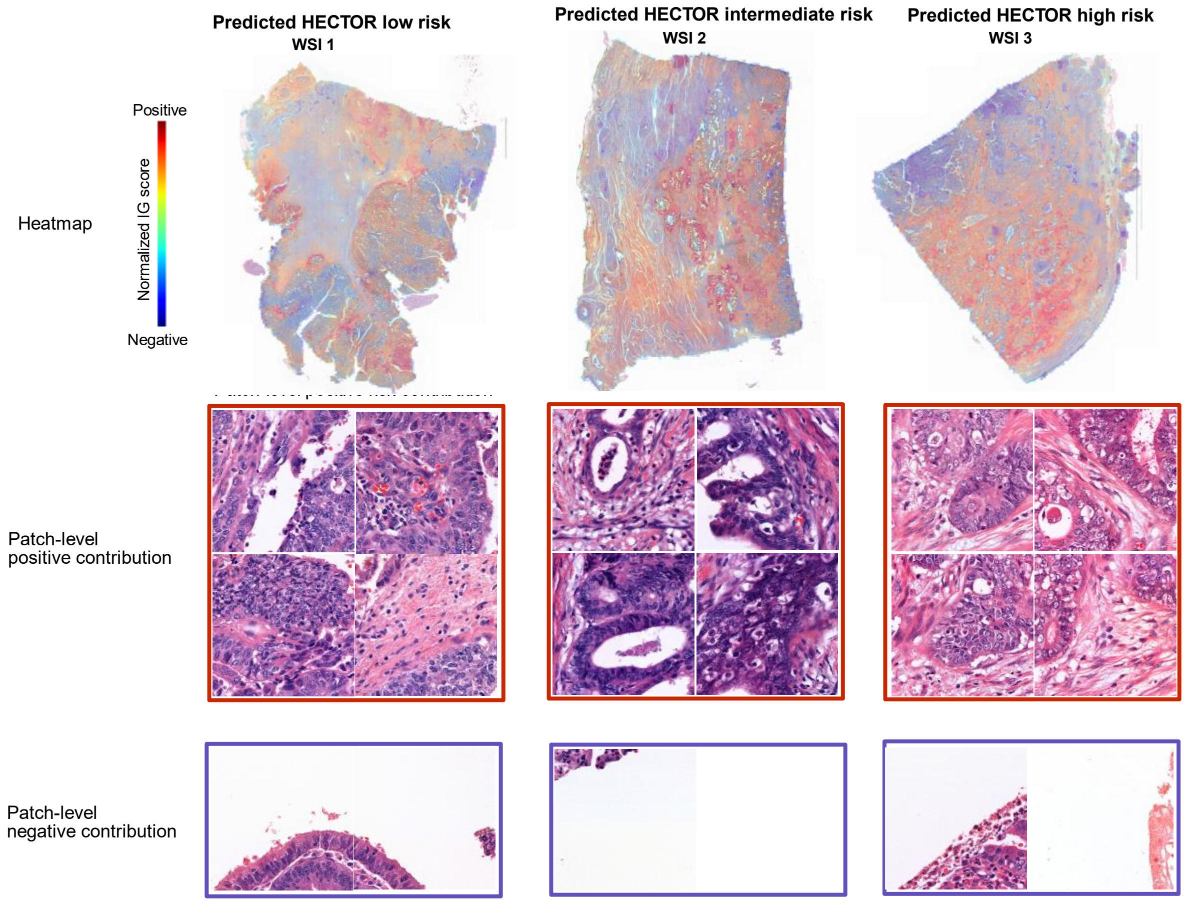


| | WSI 1 | WSI 2 | WSI 3 |
|---|---|---|---|
| Endpoint | Censored | | |
| Histological subytpe | Endometrioid | | |
| FIGO stage | III | | |
| im4MEC class | imNSMP | imNSMP | imNSMP |
| HECTOR risk group | low risk | intermediate risk | high risk |

**Figure 8:** First case out of three of the external test set (*n* = 151) with three WSIs respectively HECTOR low, intermediate and high risk. Contribution of each tissue region to the prediction was quantified with the Integrated Gradient method[13]. Im4MEC class is the image-based molecular class as predicted by im4MEC[7]. WSI = whole slide image; im = image-based; NSMP = no specific molecular profile.

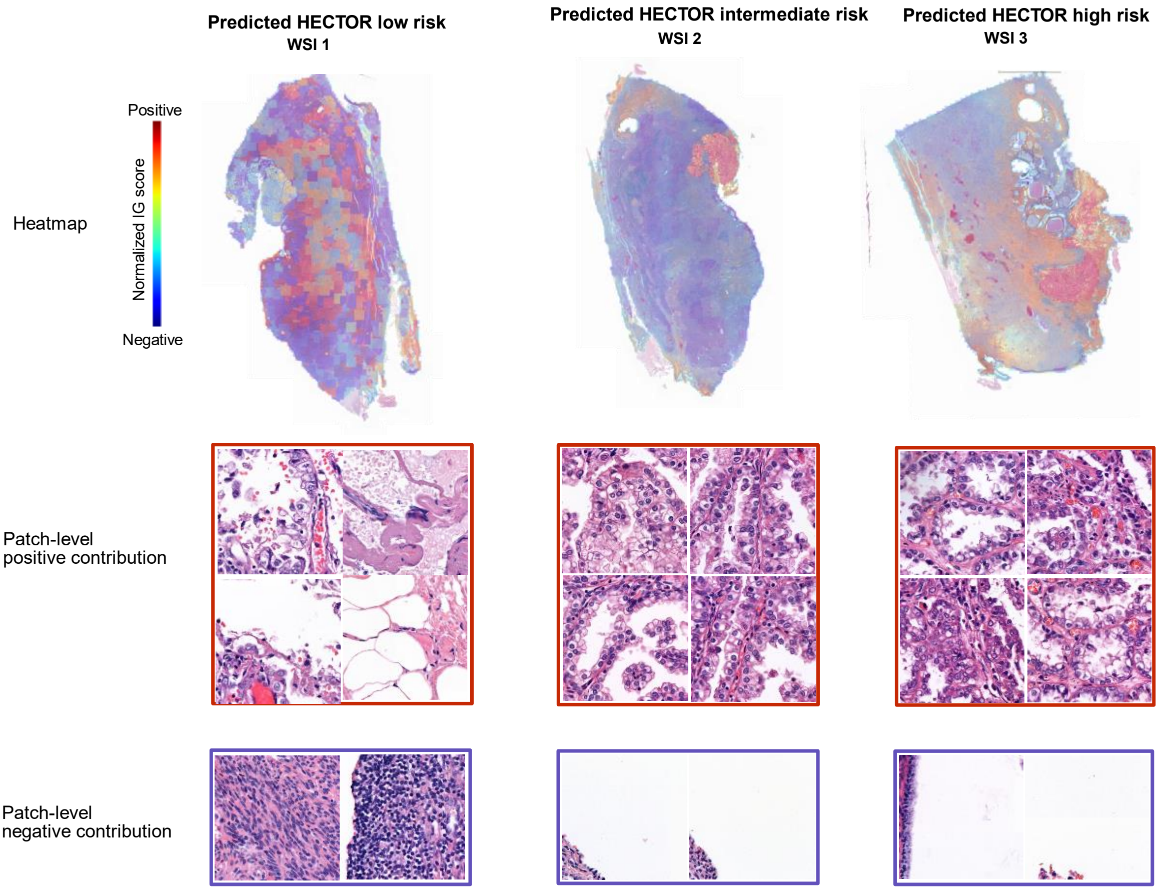


| | WSI 1 | WSI 2 | WSI 3 |
|---|---|---|---|
| Endpoint | Censored | | |
| Histological subytpe | Clear Cell | | |
| FIGO stage | II | | |
| im4MEC class | im*POLE*mut | imp53abn | imp53abn |
| HECTOR risk group | low risk | intermediate risk | high risk |

**Figure 9:** Second case out of three of the external test set (*n* = 151) with three WSIs predicted respectively HECTOR low, intermediate and high risk. Contribution of each tissue region to the prediction was quantified with the Integrated Gradient method[13]. Im4MEC class is the image-based molecular class as predicted by im4MEC[7]. WSI = whole slide image; im = image-based; *POLE*mut = *POLE* mutated; p53abn = p53 abnormal.

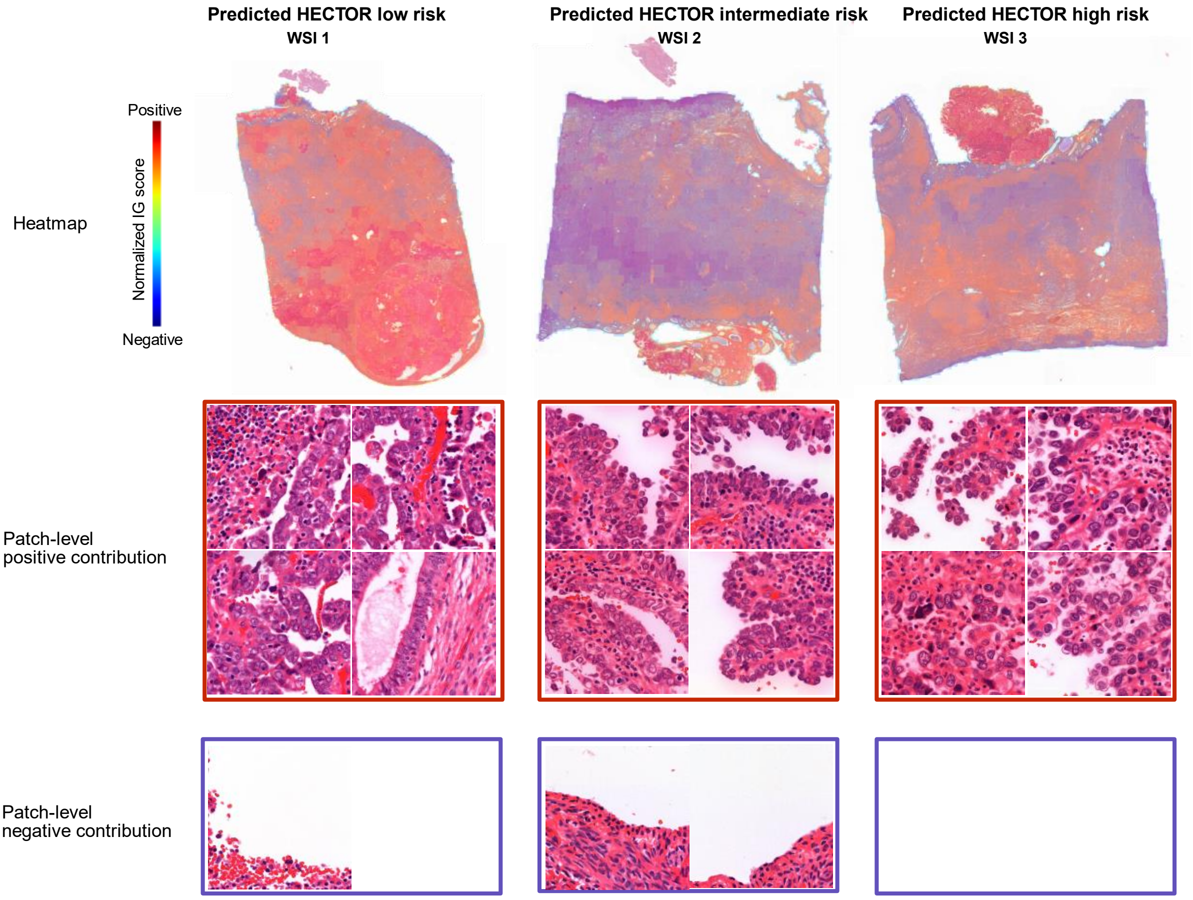


| | WSI 1 | WSI 2 | WSI 3 |
|---|---|---|---|
| Endpoint | Censored | | |
| Histological subytpe | Clear Cell | | |
| FIGO stage | I | | |
| im4MEC class | im*POLE*mut | im*POLE*mut | imp53abn |
| HECTOR risk group | low risk | intermediate risk | high risk |

**Figure 10:** Third case out of three of the external test set ($n$ = 151) with three WSIs predicted respectively HECTOR low, intermediate and high risk. Contribution of each tissue region to the prediction was quantified with the Integrated Gradient method[13]. Im4MEC class is the image-based molecular class as predicted by im4MEC[7]. WSI = whole slide image; im = image-based; *POLE*mut = *POLE* mutated; p53abn = p53 abnormal.

**Notes:**

Here in Supplementary Fig. 7a, two experiments performed in the external test set ($n$ = 151 patients; $n$ = 121 with three WSIs; $n$ = 21 with two and $n$ = 9 with one) are shown. The first one consisted of predicting the risk score separately for each WSI of each patient followed by taking the median of these scores (referred to as 'post-aggregation by median'). The second experiment consisted of merging all WSIs of each patient into one feature bag per patient, resulting in one risk score per patient (referred to as 'pre-merged'). These two aggregation methods resulted in different risk group predictions for only three cases which moved from HECTOR high risk using the pre-merged method to HECTOR low risk in the post-aggregated median method (since in the latter, two WSIs were predicted as HECTOR low risk and one WSI predicted as HECTOR high risk). This might indicate that, when WSIs are pre-merged and considered as a single input image, the visual features of the WSI predicted as HECTOR high risk may overrule the information present in the other two WSIs.

In Supplementary Fig. 7b, we showed that a number of patients with multiple WSIs had different HECTOR predictions for each WSI. Among the 142 cases with more than one WSI, 85 cases (59.9%) had consistent HECTOR risk group predictions across the WSIs. Only four cases (2.8%) had at least one WSI predicted as HECTOR low risk while another was predicted as HECTOR high risk. Only three cases (2.1%) each with three WSIs had a different predicted HECTOR risk group for each WSI. Upon review of these three cases, two cases showed the same pattern: one WSI was predicted as HECTOR low risk combined with im*POLE*mut, one WSI was predicted as HECTOR high risk and imp53abn which is in line with the respective negative and positive contributions to the risk score of the im*POLE*mut and imp53abn image-based molecular class. These three cases also showed heterogeneous tumor volumes and tumor features, including one case with the WSI predicted as HECTOR low risk presenting smooth luminal borders when the destructive component of the remaining two WSIs was more substantial (Supplementary Figs. 8-10).

| Variable | Coefficient [95% CI] | *P* value |
|---|---|---|
| Endometrioid (EEC) | -1.124 [-1.337;-0.910] | <0.001 |
| Serous (SEC) | 1.025 [0.722;1.328] | <0.001 |
| Clear cell (CCC) | 0.862 [0.416;1.309] | <0.001 |
| Un/de-differentiated | 1.212 [0.288;2.137] | 0.010 |
| Carcinosarcoma | 1.708 [0.794;2.621] | <0.001 |
| Grade 1 (GR1) | -0.751 [-0.923;-0.580] | <0.001 |
| Grade 2 (GR2) | -0.035 [-0.347;0.276] | 0.824 |
| Grade 3 (GR3) | 0.794 [0.622;0.967] | <0.001 |
| FIGO Stage I | -0.683 [-0.904;-0.463] | <0.001 |
| FIGO Stage II | 0.215 [-0.177;0.607] | 0.282 |
| FIGO Stage III | 0.801 [0.548;1.053] | <0.001 |
| LVSI presence | 0.526 [0.237;0.814] | <0.001 |
| *POLE*mut | -0.383 [-0.724;-0.042] | 0.028 |
| im*POLE*mut | -0.320 [-0.644;0.004] | 0.053 |
| MMRd | -0.208 [-0.430;0.014] | 0.067 |
| imMMRd | -0.228 [-0.442;-0.014] | 0.037 |
| p53abn | 1.028 [0.827;1.229] | <0.001 |
| imp53abn | 1.253 [1.066;1.439] | <0.001 |
| NSMP | -0.441 [-0.627;-0.256] | <0.001 |
| imNSMP | -0.543 [-0.727;-0.359] | <0.001 |
| ER negative | 0.948 [0.708;1.187] | <0.001 |
| L1CAM positive | 1.007 [0.802;1.211] | <0.001 |

**Table 6:** Coefficients of single linear regression between HECTOR risk scores and each variable. CI = confidence interval; LVSI = lymphovascular space invasion; MMRd = Mismatch repair deficient; NSMP = no specific molecular profile; p53abn = p53 abnormal; im = image-based predicted from im4MEC[7].

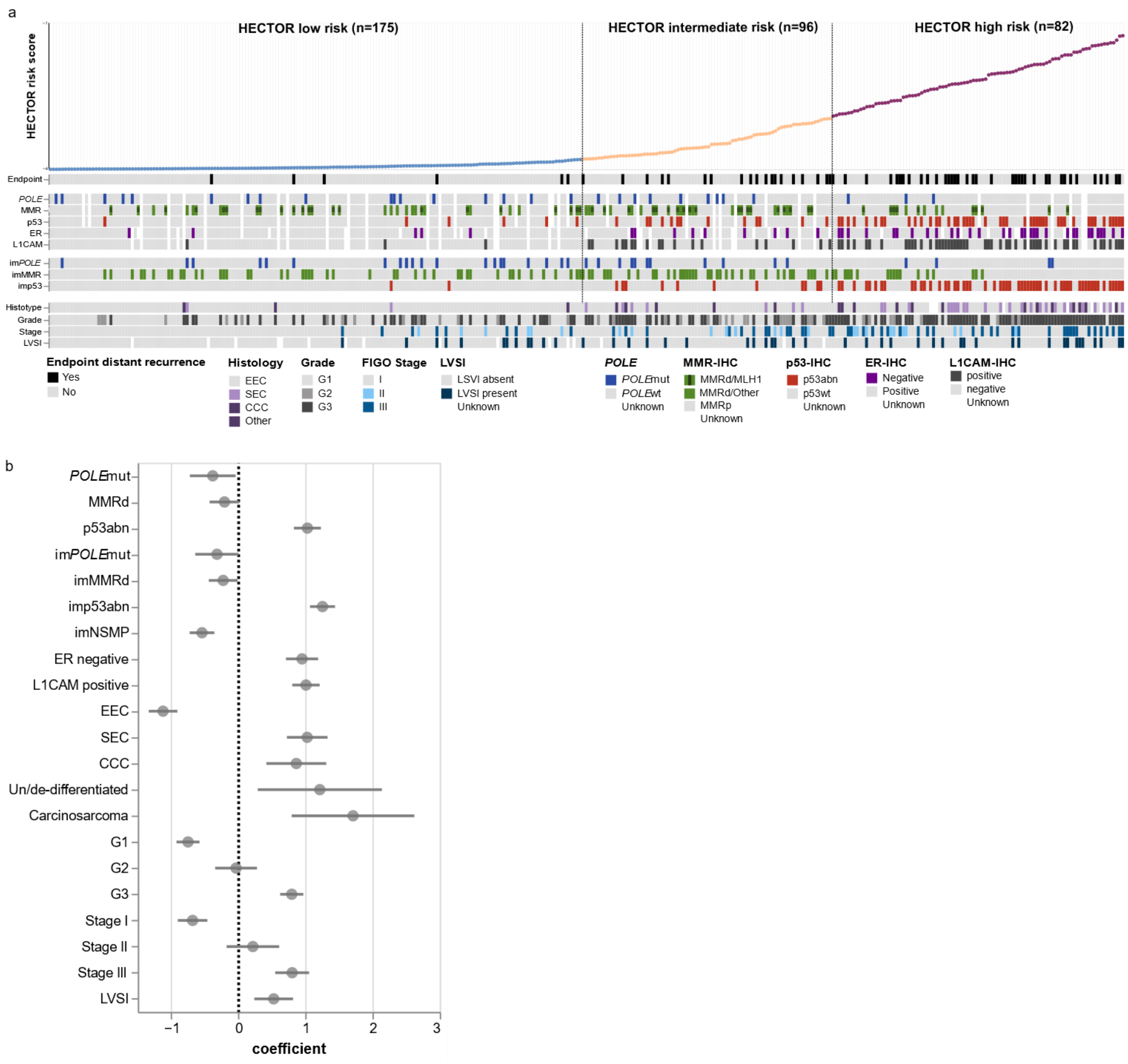


**Figure 11:** a, Extended heatmap of risk factors for patients included in the internal test set ($n$ = 353) ordered by predicted HECTOR risk scores, along with their image-based molecular class as predicted by im4MEC[7]. b, Coefficients and confidence intervals of the risk factors derived from separate single linear regression models with the continuous risk score as the target variable.

| n=1408 | im*POLE* mut | imMMRd | imp53abn | imNSMP |
|---|---|---|---|---|
| *POLE* mut | 111 | 6 | 4 | 1 |
| MMRd | 8 | 367 | 6 | 3 |
| p53abn | 0 | 1 | 246 | 0 |
| NSMP | 2 | 5 | 8 | 525 |
| Unkown | 14 | 29 | 13 | 59 |

| n=353 | m*POLE* mu | imMMRd | imp53abn | imNSMP |
|---|---|---|---|---|
| *POLE* mut | 11 | 13 | 1 | 3 |
| MMRd | 14 | 49 | 3 | 9 |
| p53abn | 2 | 1 | 43 | 7 |
| NSMP | 4 | 23 | 15 | 117 |
| Unkown | 4 | 12 | 5 | 17 |

**Table 7:** Confusion matrices of im4MEC[7] in the training set (left) and internal test set (right).

| | HECTOR low-risk | HECTOR intermediate-risk | HECTOR high-risk | Total |
|---|---|---|---|---|
| **Total** | **175** | **82** | **96** | **353** |
| **Histotype** | | | | |
| Endometrioid | 170 (58%) | 70 (24%) | 51 (18%) | **291** |
| Serous | 2 (6%) | 7 (21%) | 24 (73%) | **33** |
| Clear cells | 0 (0%) | 4 (24%) | 13 (76%) | **17** |
| Un/de-differentiated | 0 (0%) | 0 (0%) | 3 (100%) | **3** |
| Carcinosarcoma | 0 (0%) | 0 (0%) | 3 (100%) | **3** |
| Other | 3 (50%) | 1 (17%) | 2 (33%) | **6** |
| **Grade** | | | | |
| 1 | 123 (72%) | 32 (19%) | 15 (9%) | **170** |
| 2 | 16 (39%) | 17 (41%) | 8 (20%) | **41** |
| 3 | 36 (25%) | 33 (23%) | 73 (51%) | **142** |
| **2009 FIGO stage** | | | | |
| IA | 52 (55%) | 17 (18%) | 26 (27%) | **95** |
| IB | 109 (60%) | 44 (24%) | 30 (16%) | **183** |
| II | 6 (25%) | 8 (33%) | 10 (42%) | **24** |
| IIIA | 2 (10%) | 6 (29%) | 13 (62%) | **21** |
| IIIB | 0 (0%) | 1 (11%) | 8 (89%) | **9** |
| IIIC | 6 (29%) | 6 (29%) | 9 (43%) | **21** |
| **LVSI** | | | | |
| Present | 12 (26%) | 13 (28%) | 22 (47%) | **47** |
| Absent or focal | 150 (52%) | 66 (23%) | 74 (26%) | **290** |
| Unkown | 13 (81%) | 3 (19%) | 0 (0%) | **16** |
| **Molecular class** | | | | |
| *POLE* mut | 17 (59%) | 9 (31%) | 3 (10%) | **29** |
| *POLEwt* | 135 (47%) | 64 (22%) | 87 (30%) | **286** |
| MMRd | 36 (46%) | 29 (37%) | 14 (18%) | **79** |
| MMRp | 116 (49%) | 44 (19%) | 76 (32%) | **236** |
| p53abn | 6 (9%) | 13 (20%) | 45 (70%) | **64** |
| p53wt | 146 (58%) | 60 (24%) | 45 (18%) | **251** |
| Unkown | 23 (61%) | 9 (24%) | 6 (16%) | **38** |
| **ER IHC** | | | | |
| Negative | 5 (11%) | 9 (20%) | 31 (69%) | **45** |
| Positive | 144 (57%) | 56 (22%) | 52 (21%) | **252** |
| Unkown | 26 (46%) | 17 (30%) | 13 (23%) | **56** |
| **L1CAM** | | | | |
| Negative | 153 (61%) | 55 (22%) | 42 (17%) | **250** |
| Positive | 3 (4%) | 18 (26%) | 49 (70%) | **70** |
| Unkown | 19 (58%) | 9 (27%) | 5 (15%) | **33** |
| **im4MEC** | | | | |
| im*POLE* mut | 20 (57%) | 11 (31%) | 4 (11%) | **35** |
| imMMRd | 45 (46%) | 36 (37%) | 17 (17%) | **98** |
| imp53abn | 2 (3%) | 11 (16%) | 54 (81%0 | **67** |
| imNSMP | 108 (71%) | 24 (16%) | 21 (14%) | **153** |

**Table 8:** Composition of the HECTOR risk groups with clinicopathological prognostic factors and guideline prognostic groups. G = grade; EEC = endometrioid. LVSI = lymphovascular space invasion; *POLE*mut = *POLE* mutated; *POLE*wt = *POLE* wild type; MMRd = Mismatch repair deficient; MMRp = Mismatch repair proficient; NSMP = no specific molecular profile; p53abn = p53 abnormal; p53wt = p53 wild type; im = image-based predicted from im4MEC[7].

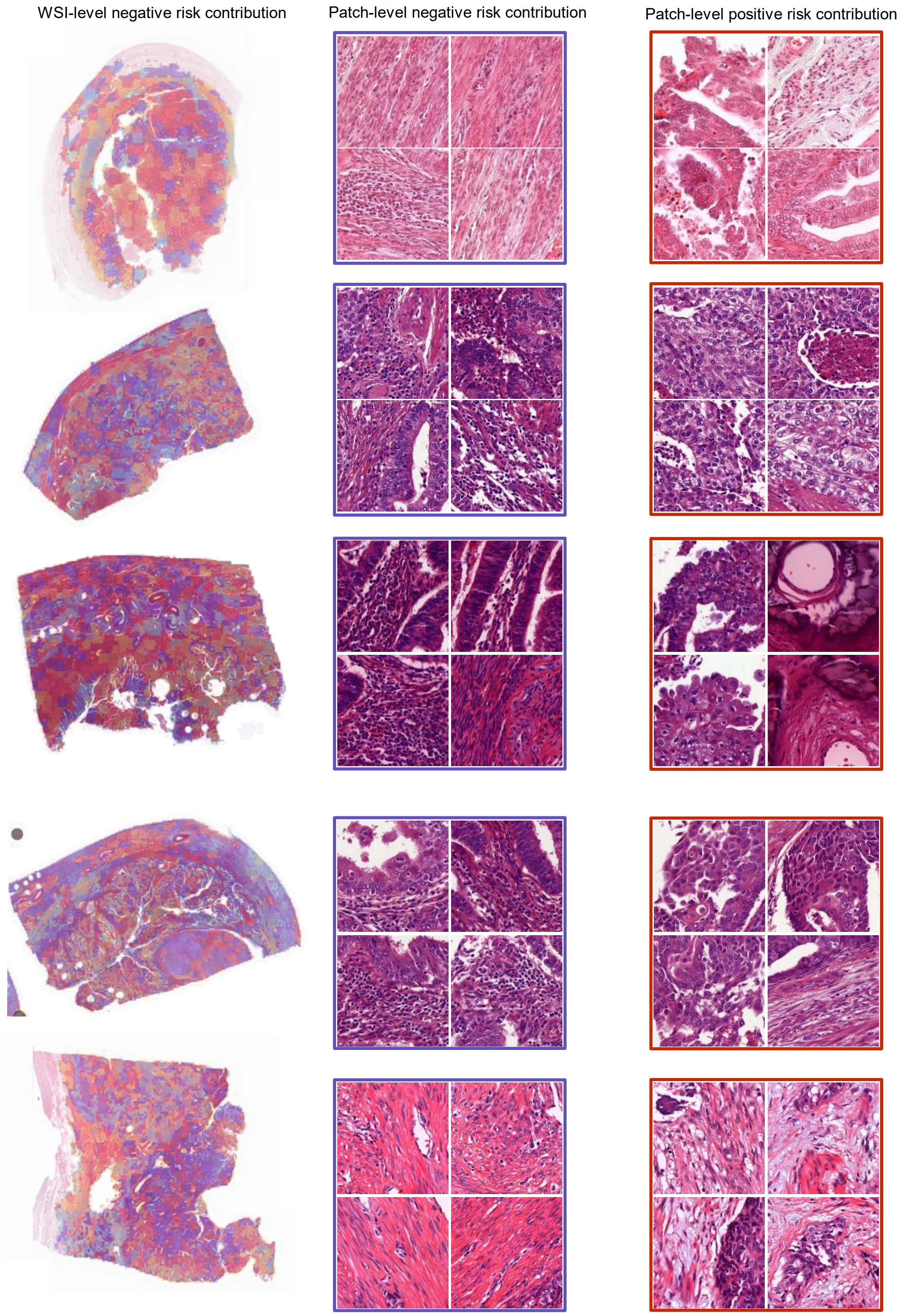


**Figure 12:** Cases with whole slide image (WSI)-level negative risk contribution as quantified using the Integrated Gradient method[13] (that is by avargeing all patch-level contribution scores given by the Integrated Gradient method). Left shows the heatmap overlay of the contributions given to each patch. Middle and left show a selection of four patches with high negative and positive contribution.

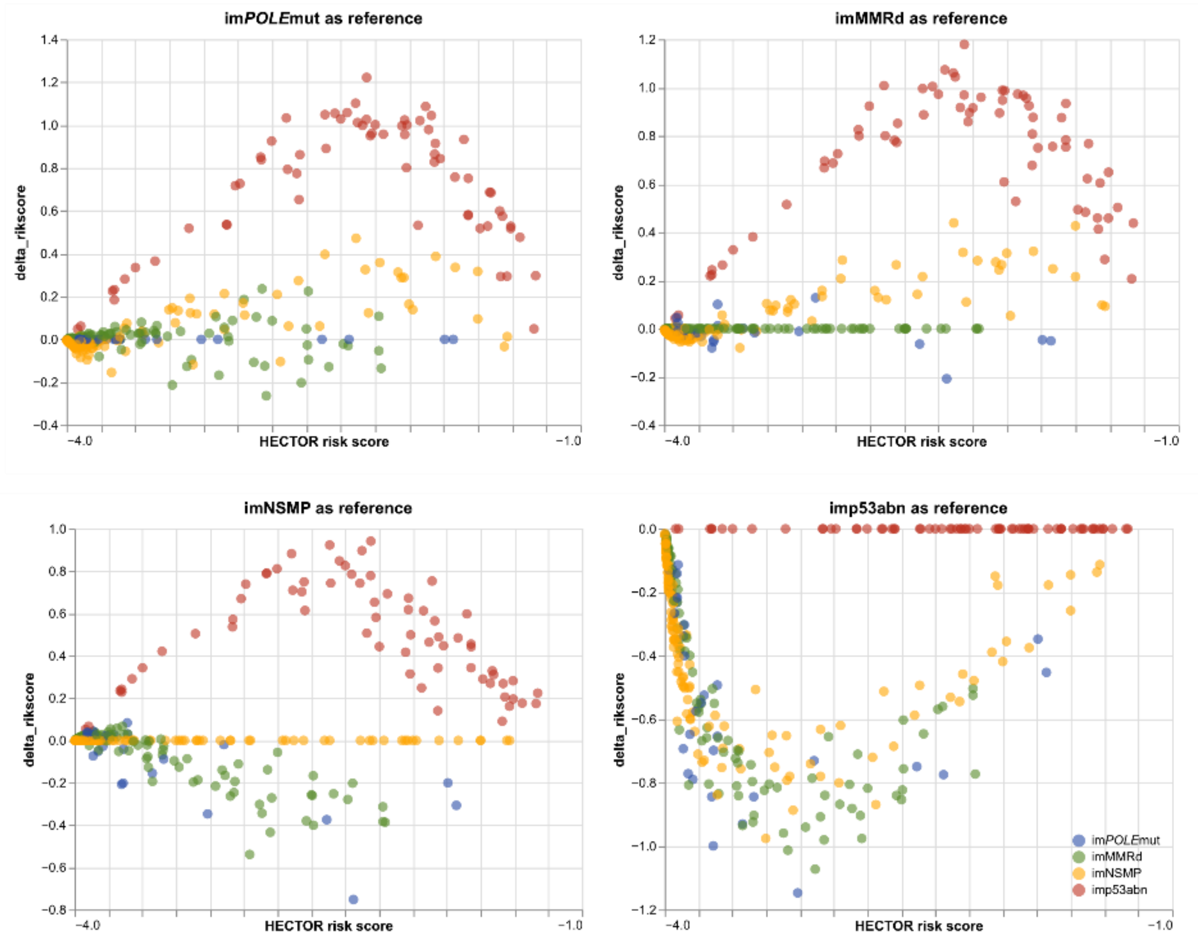


**Figure 13:** Contribution each the image-based molecular class (im4MEC) to the HECTOR risk scores. The difference in predicted HECTOR risk score is computed between the HECTOR risk score given by the image-based molecular class as precited by im4MEC[7] and the one given by hard-coding the image-based molecular class, referred to as “the reference group”: (top left) im*POLE*mut is the reference group; (top right) imMMRd is the reference group; (bottom left) imNSMP is the reference group; (bottom right) imp53abn is the reference group. im = image-based; MMRd = Mismatch repair deficient; NSMP = no specific molecular profile; p53abn = p53 abnormal.

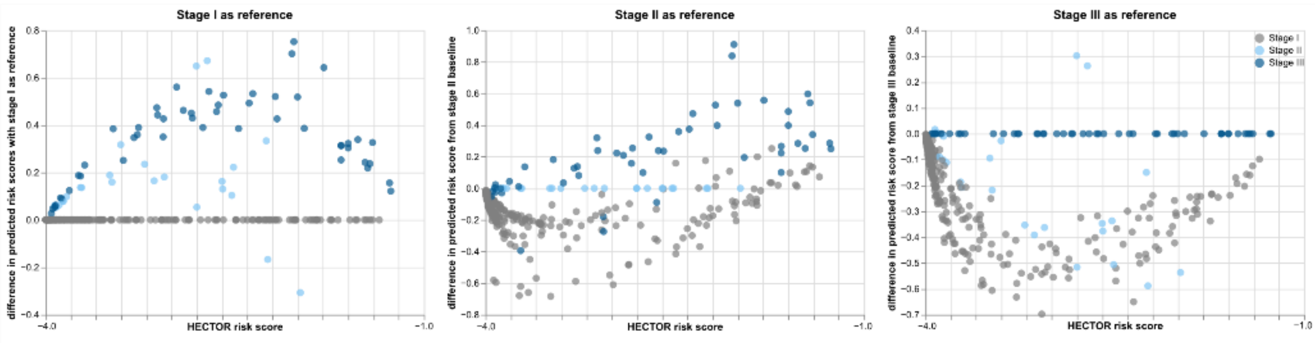


**Figure 14:** Contribution of the image-based molecular class as predicted by im4MEC to the HECTOR risk score. The difference in predicted risk score is computed between the risk score given by the FIGO stage and the reference group: (left) 2009 FIGO stage I is the reference group; (middle) 2009 FIGO stage II is the reference group; (bottom) 2009 FIGO stage III is the reference group.

| | Registered histological subtype/LVSI | Review by expert of the single input WSI | 2009 FIGO Stage | Image-based molecular class | HECTOR risk group | True molecular class | HECTOR risk group with true molecular class |
|---|---|---|---|---|---|---|---|
| Case 1 | G1 EEC / no LVSI | Concordant | I | imNSMP | Low risk | *POLE*mut | Low risk |
| Case 2 | G3 EEC / no LVSI | Concordant | I | im*POLE*mut | Low risk | MMRd | Low risk |
| Case 3 | G3 EEC / no LVSI | Concordant | I | imNSMP | Low risk | NSMP | Low risk |
| Case 4 | G1 EEC / LVSI | No LVSI | III | im*POLE*mut | Low risk | NSMP | Low risk |
| Case 5 | G1 EEC / no LVSI | Concordant | II | im*POLE*mut | Low risk | NSMP | Low risk |
| Case 6 | Other Non-EEC / Unknown | Endometrioid histology | I | im*POLE*mut | Low risk | Unknown | - |

**Table 9:** Clinicopathological data and HECTOR risk group predictions of the cases predicted as HECTOR low risk that had distant recurrence in the internal test set (*n* = 6 out of 353 patients). The image-based (im) molecular class is produced by the im4MEC arm[7].

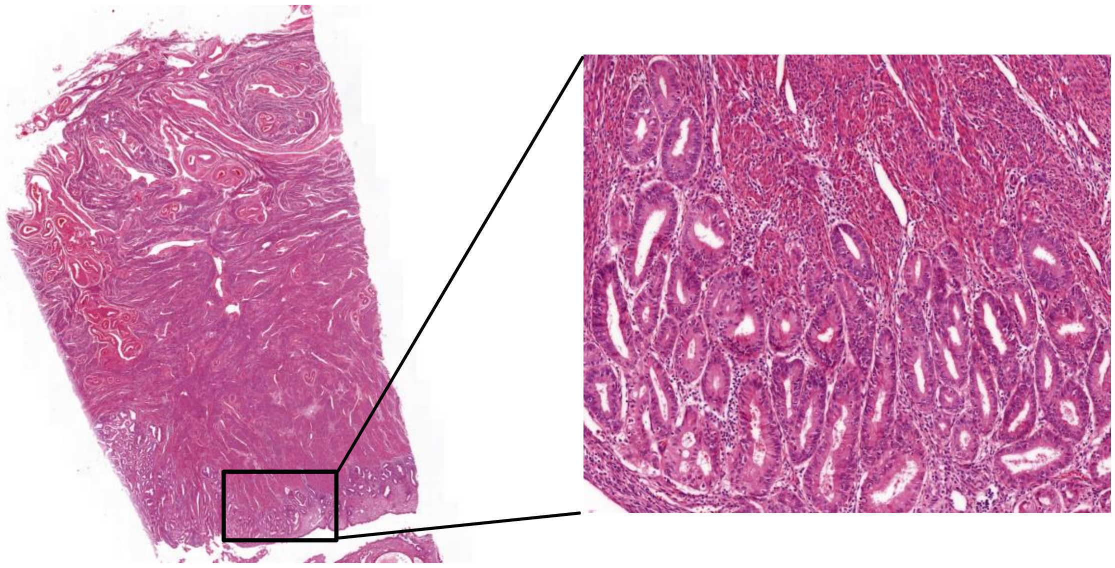

**Figure 15:** Example of a case with discrepancy between registered non-endometrioid histological subtype and the review of the input WSI by an expert, showing an endometrioid histology. Distant recurrence predicted HECTOR low risk in the internal test set (*n* = 353 patients).

HECTOR low risk & negative contribution - Intra-epithelial neutrophils

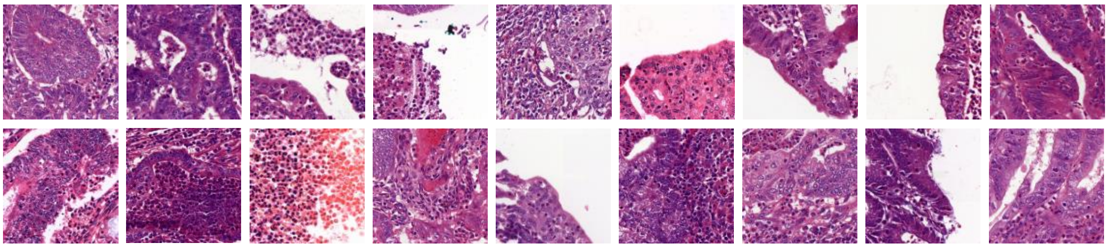

HECTOR low risk & negative contribution - Tumour infiltrating lymphocytes

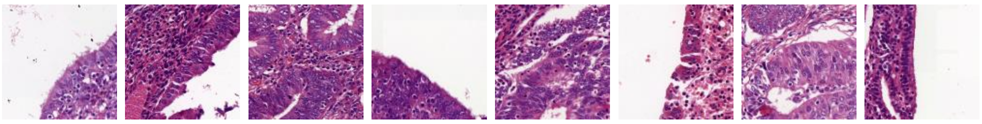

HECTOR low risk & negative contribution - Peritumoral lymphocytes

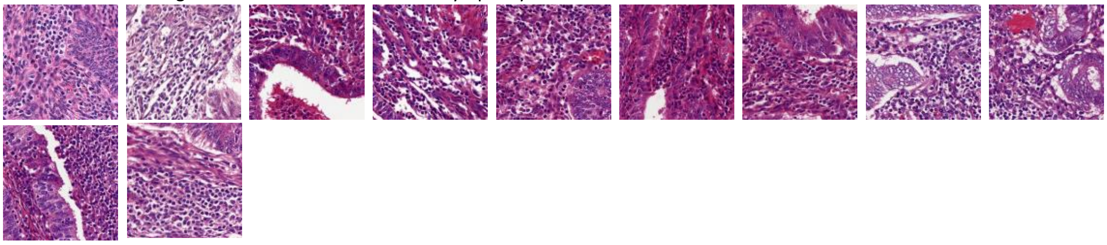

HECTOR low risk & negative contribution - Compact normal myometrium

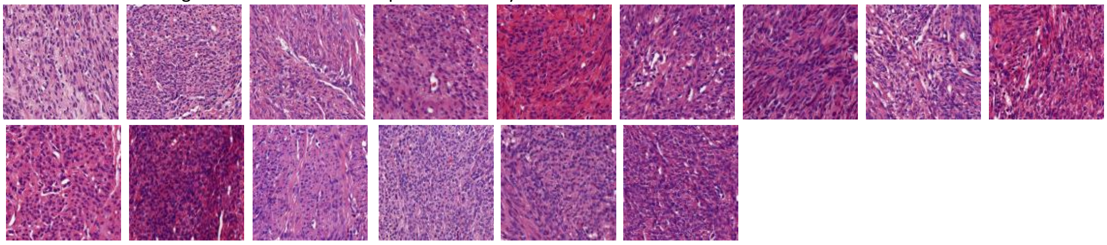

HECTOR low risk & negative contribution - Smooth luminal borders

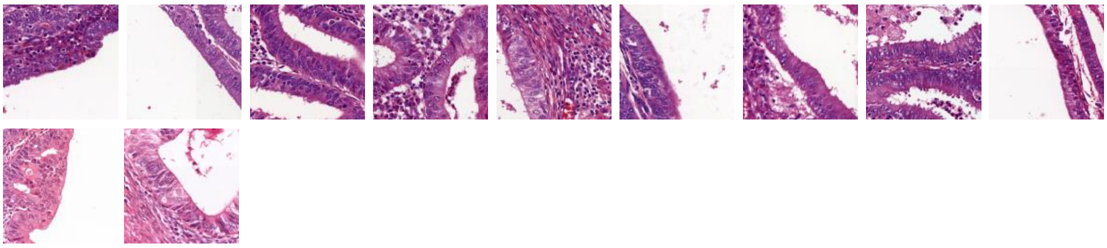

**Figure 16:** Additional representative patches which strongly decreased the HECTOR risk scores from all WSIs in the HECTOR low risk group (top 5% patches with negative contribution using the Integrated Gradient method[13]).

HECTOR high risk & negative contribution - Compact normal myometrium

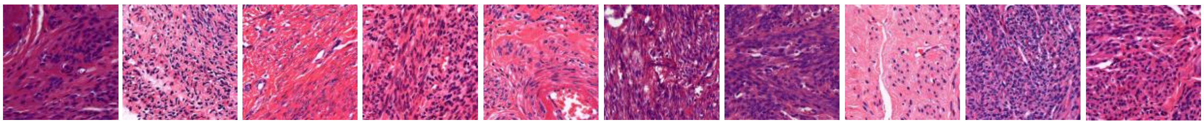

HECTOR high risk & negative contribution - Peritumoral lymphocytes

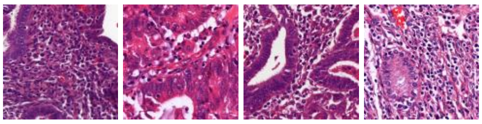

HECTOR high risk & negative contribution - Tumour infiltrating lymphocytes

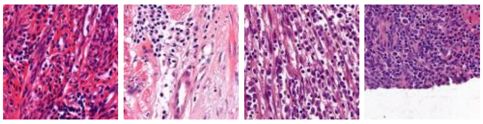

**Figure 17:** Additional representative patches which strongly decreased the HECTOR risk scores from all WSIs in the HECTOR high risk group (top 5% patches with negative contribution using the Integrated Gradient method[13]).

HECTOR high risk & positive contribution - Desmoplastic stromal reaction

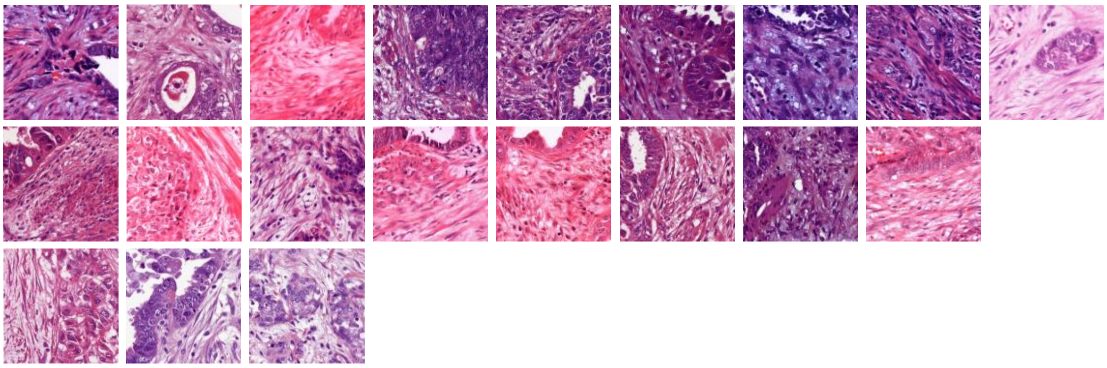

HECTOR high risk & positive contribution - Destructive invasion/trabecular growth

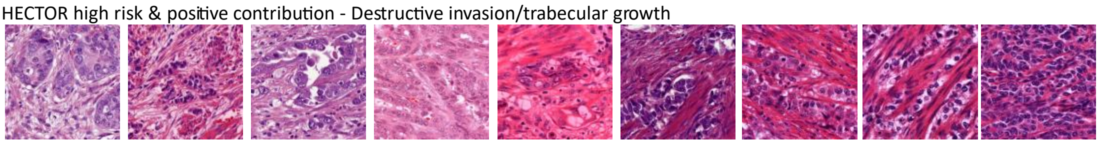

HECTOR high risk & positive contribution - Hobnailing

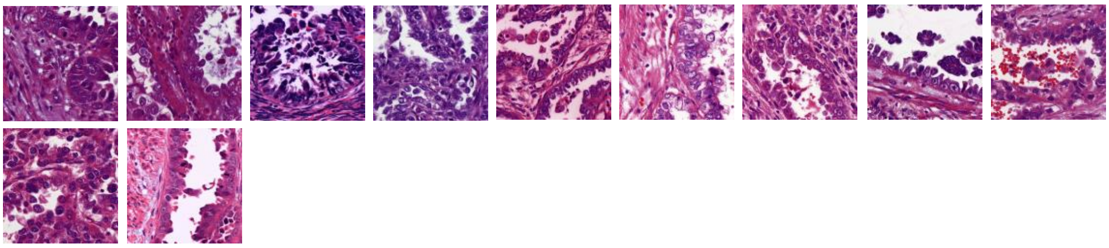

HECTOR high risk & positive contribution - Solid growth/nuclear atypia

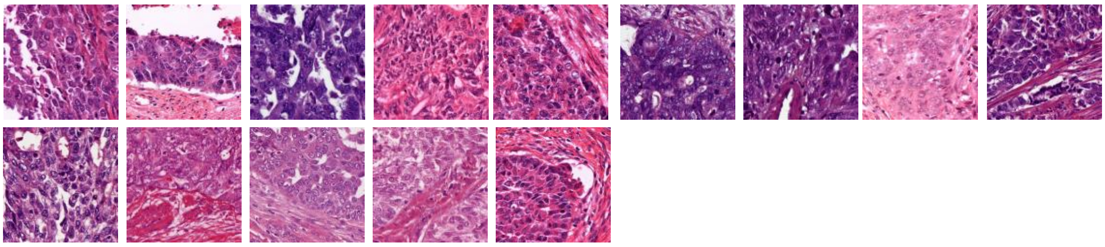

**Figure 18:** Additional representative patches which strongly increased the HECTOR risk scores from all WSIs in the HECTOR low risk group (top 5% patches with positive contribution using the Integrated Gradient method[13]).

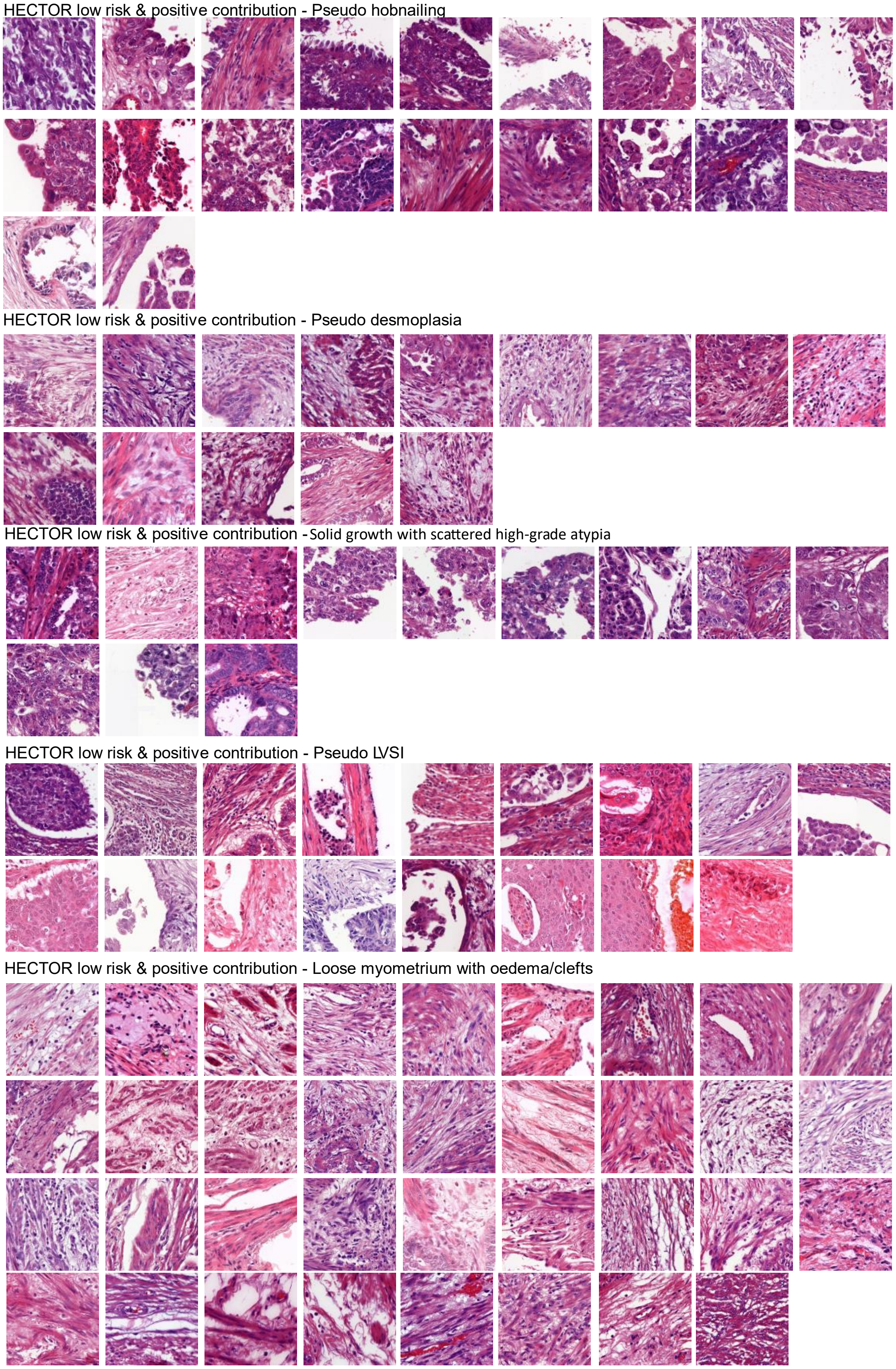

**Figure 19:** Additional representative patches which strongly increased the HECTOR risk scores from all WSIs in the HECTOR low risk group (top 5% patches with positive contribution using the Integrated Gradient method[13]).

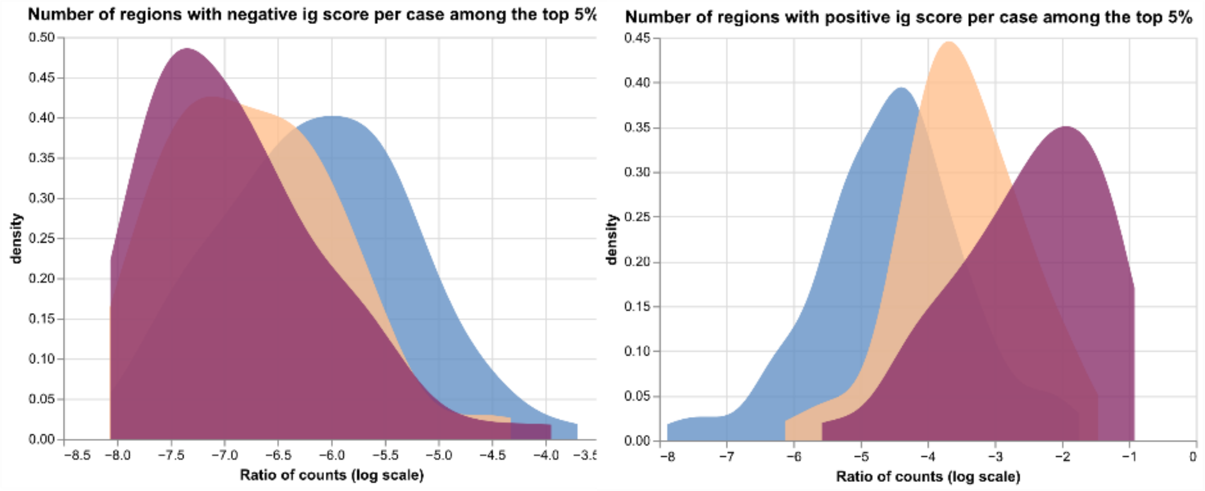


**Figure 20:** Density plots showing the patient-level ratio of the number of regions with negative or positive contribution among all top 5% regions with negative (left) or positive contribution (right) as quantified by the Integrated Gradient method[13].

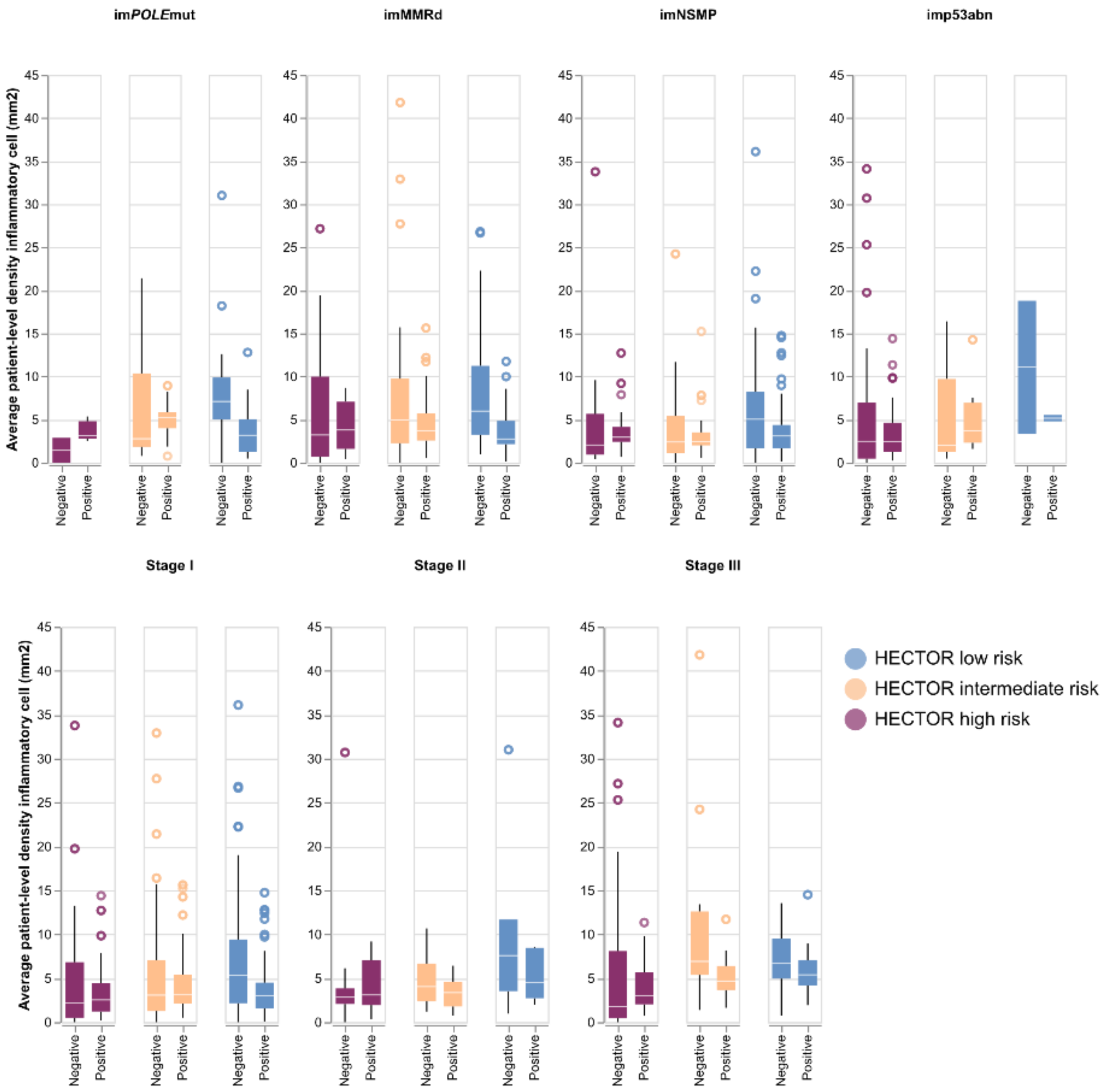


**Figure 21:** Average patient-level inflammatory cell density within the top 5% regions with positive and negative contribution, using the Integrated Gradient method[13], stratified by image-based molecular class (top) and 2009 FIGO stage I-III (bottom).

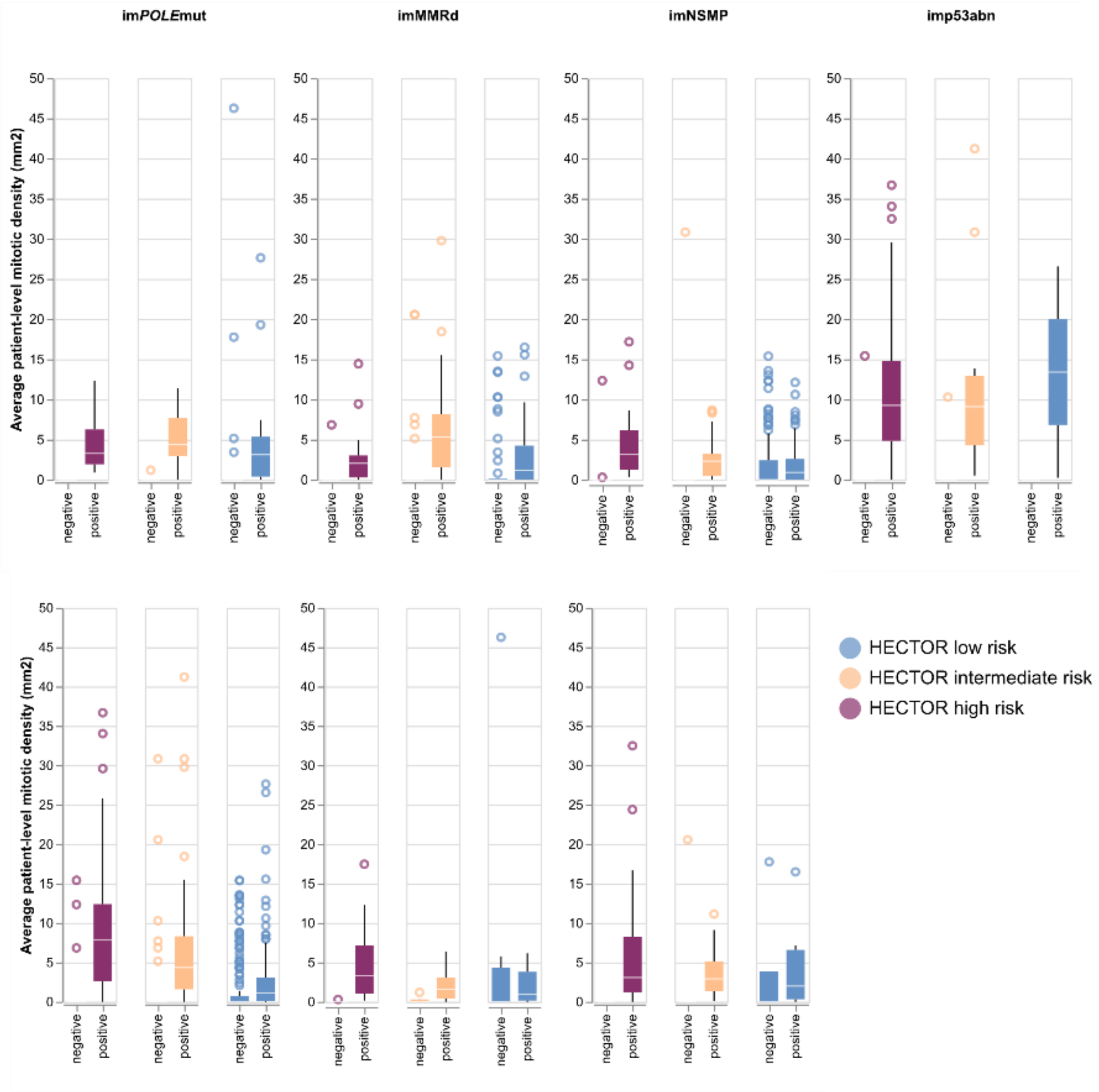


**Figure 22:** Average patient-level mitotic density within the top 5% regions with positive and negative Integrated Gradient[13] score filtered by regions, stratified by image-based molecular class[7] (top) and 2009 FIGO stage I-III (bottom).

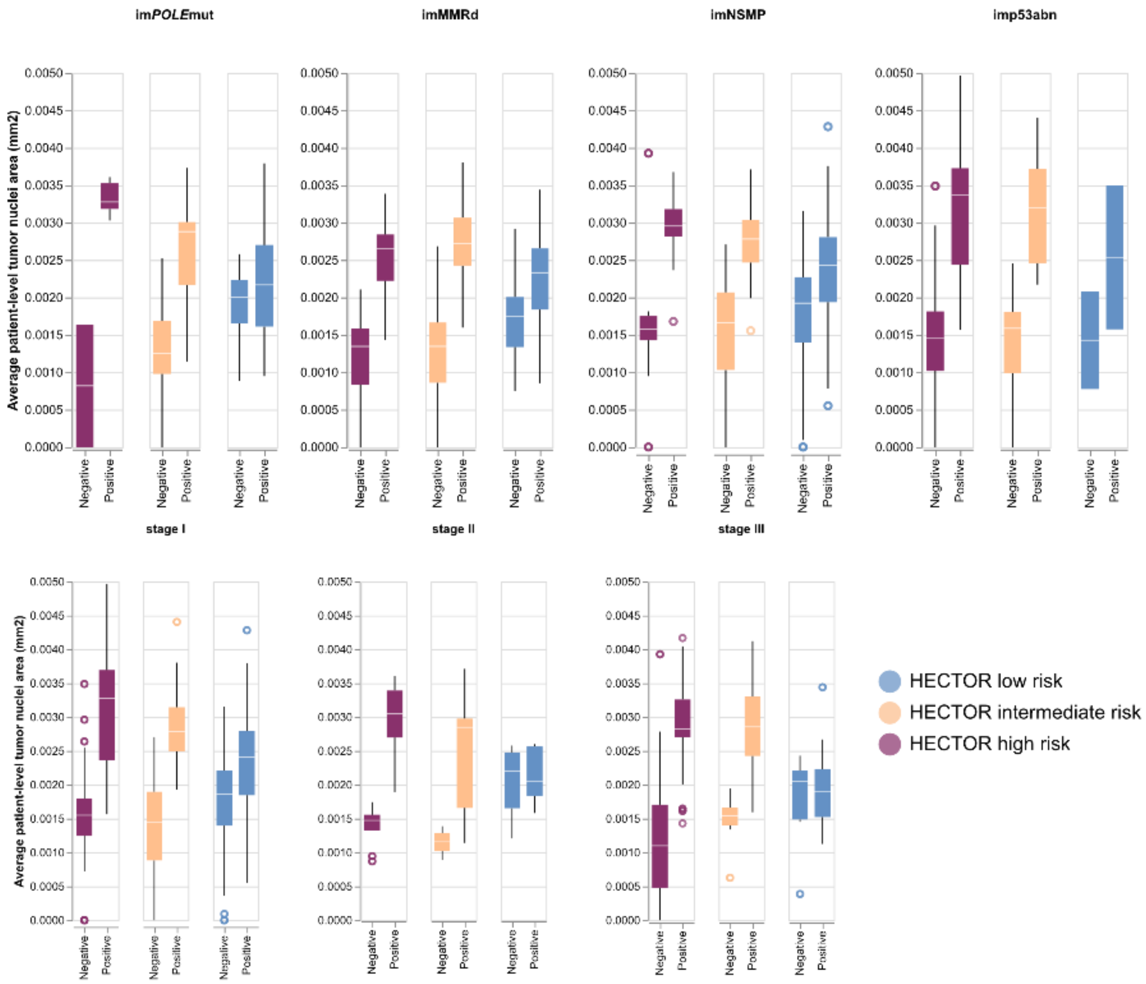


**Figure 23:** Average patient-level area of tumor nuclei within the top 5% regions with positive and negative Integrated Gradient[13] score filtered by regions, stratified by image-based molecular class[7] (top) and 2009 FIGO stage I-III (bottom).

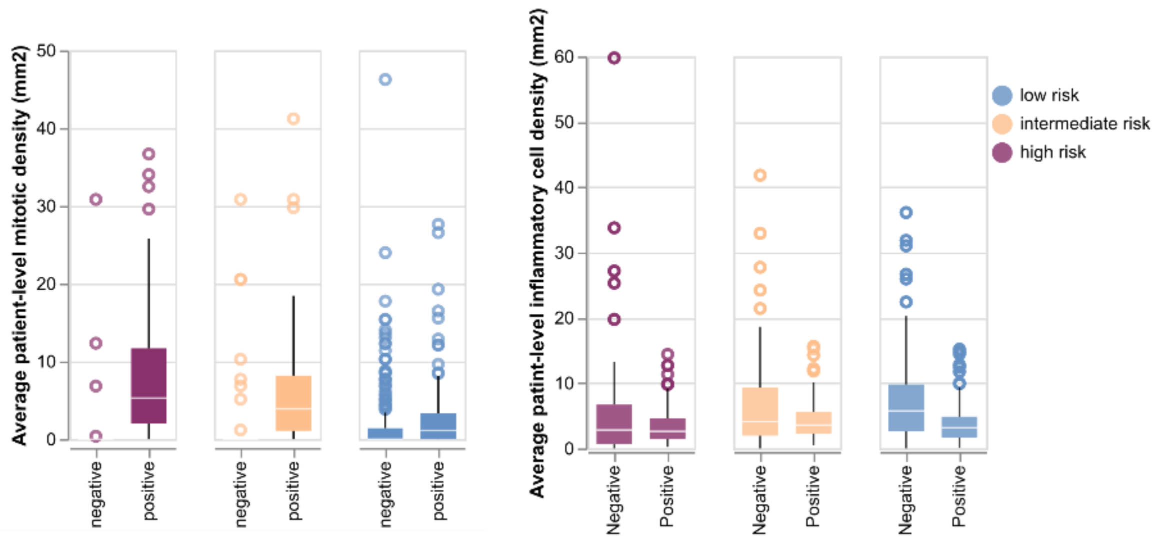


**Figure 24:** Average patient-level inflammatory cell count (left) and mitotic figure count (right) within the top 5% regions with positive and negative Integrated Gradient[13] score filtered by regions that contained tumour cells as predicted by Hover-Net[14].

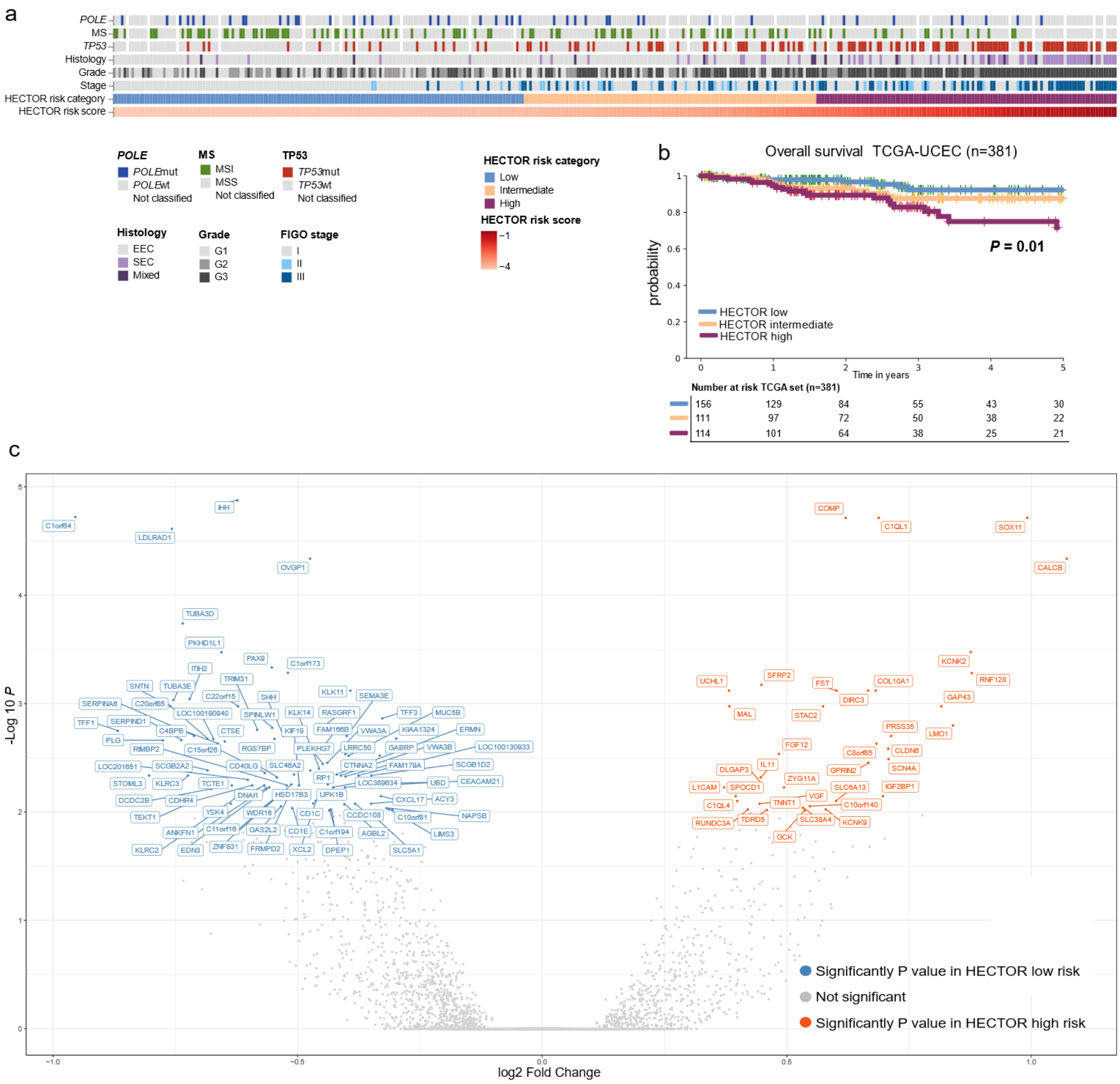


**Figure 25:** Analysis in TCGA-UCEC ($n$ = 381 patients) a, Association with clinicopathological data and HECTOR risk scores. b, Six-year overall survival analysis using the Kaplan Meier's method stratified by HECTOR risk group. c, Differential gene expression analysis by DESeq2[15]. Genes with a Benjamini–Hochberg FDR below 0.050 were selected as significantly different.

| | *P* value of test between HECTOR low, intermediate, high risk groups | *P* value of test between HECTOR low and high risk groups |
|---|---|---|
| *ARID1A* | 3.74e-05 | 6.11e-06 |
| *BCOR* | 0.304 | 0.197 |
| *CTCF* | 1.57e-04 | 7.50e-05 |
| *CTNNB1* | 8.47e-06 | 2.28e-06 |
| *FAT1* | 0.166 | 0.0606 |
| *FBXW7* | 0.208 | 0.365 |
| *FGFR2* | 9.12e-04 | 1.59e-03 |
| *KMT2C* | 0.113 | 0.297 |
| *KMT2D* | 0.447 | 0.209 |
| *KRAS* | 1.99e-03 | 3.59e-04 |
| *NOTCH1* | 0.180 | - |
| *NSD1* | 0.166 | 0.0593 |
| *PIK3CA* | 1.11e-03 | 5.23e-04 |
| *PIK3R1* | 7.29e-04 | 1.77e-04 |
| *PPP2R1A* | 2.19e-03 | 4.65e-04 |
| *PTEN* | 1.51e-09 | 4.24e-10 |
| *SPOP* | 0.417 | 0.346 |
| *TP53* | 2.81e-07 | 4.13e-07 |
| *ZFHX3* | 0.112 | 0.0411 |

**Table 10:** *P* values of the Chi square tests to compare proportions with oncogenic mutations between HECTOR risk groups on an individual gene basis.

| | Unadjusted | | Adjusted for EC molecular class | | Adjusted for EC molecular class and TMB | |
|---|---|---|---|---|---|---|
| Immune cells | Coefficient | *P* value | Coefficient | *P* value | Coefficient | *P* value |
| B cells memory | 0.068 | 8.47e-3 | 0.0474 | 0.137 | ND | ND |
| Denditric cells activated | 0.078 | 7.88e-4 | 0.0564 | 0.0419 | ND | ND |
| Denditric cells resting | -0.031 | 0.0492 | -0.00343 | 0.857 | 0.0588 | 0.0355 |
| Mast cells resting | 0.058 | 0.0287 | 0.0255 | 0.417 | ND | ND |
| CD8 t cells | -0.247 | 6.89e-6 | -0.155 | 0.0163 | -0.165 | 0.011 |
| T cells follicular helper | -0.178 | 1.53e-4 | -0.131 | 0.0161 | -0.147 | 6.71e-3 |
| T cells regulatory T regs | -0.251 | 2.21e-09 | -0.199 | 8.02e-5 | -0.196 | 1.19e-4 |

**Table 11:** Coefficients of linear regressions performed with the log2-transformed proportion of cell subset (Leukocyte_Fraction*immune_subset) as the dependent variable, and the the HECTOR risk scores as independent variables. Regressions were adjusted for molecular subgroup and tumor mutational burden (TMB) by adding these two variables as independent variables.

| | Inclusion | PORTEC-3 |
|---|---|---|
| **Number of patients (%)** | | **660** |
| | yes | 442 (67.0%) |
| | no | 218 (33,0%) |
| **Follow-up: median** | | |
| | yes | 5.06 |
| | no | 7.66 |
| **Age: median** | | |
| | yes | 62 |
| | no | 68 |
| **Histotype (%)** | | |
| Endometrioid grade 1-2 | yes | 177 (26.8%) |
| | no | 80 (12.1%) |
| Endometrioid grade 3 | yes | 127 (19.2%) |
| | no | 58 (8.8%) |
| Serous carcinoma | yes | 64 (9.7%) |
| | no | 41 (6.2%) |
| Clear cell carcinoma | yes | 42 (6.4%) |
| | no | 20 (3.0%) |
| Carcinosarcoma | yes | 0 (0.0%) |
| | no | 0 (0.0%) |
| Un-dedifferentiated | yes | 10 (1.5%) |
| | no | 4 (0.6%) |
| Other | yes | 22 (3.3%) |
| | no | 15 (2.3%) |
| **LVSI (%)** | | |
| Present | yes | 280 (42.4%) |
| | no | 109 (16.5%) |
| Focal or absent | yes | 162 (24.5%) |
| | no | 109 (16.5%) |
| **2009 FIGO stage (%)** | | |
| IA | yes | 56 (8.5%) |
| | no | 22 (3.3%) |
| IB | yes | 79 (12.0%) |
| | no | 38 (5.8%) |
| II | yes | 114 (17.3%) |
| | no | 56 (8.5%) |
| IIIA | yes | 50 (7.6%) |
| | no | 33 (5.0%) |
| IIIB | yes | 31 (4.7%) |
| | no | 11 (1.7%) |
| IIIC | yes | 112 (17.0%) |
| | no | 58 (8.8%) |
| IV | yes | 0 (0.0%) |
| | no | 0 (0.0%) |
| **Molecular class (%)** | | |
| *POLE* mut | yes | 50 (7.6%) |
| | no | 1 (0.2%) |
| MMRd | yes | 136 (20.6%) |
| | no | 3 (0.5%) |
| NSMP | yes | 120 (18.2%) |
| | no | 2 (0.3%) |
| p53abn | yes | 88 (13.3%) |
| | no | 11 (1.7%) |
| Unkown | yes | 48 (7.3%) |
| | no | 201 (30.5%) |
| **Adjuvant treatment (%)** | | |
| EBRT alone | yes | 214 (32.4%) |
| | no | 116 (17.6%) |
| Chemotherapy + EBRT | yes | 228 (34.5%) |
| | no | 102 (15.5%) |

**Table 12:** Characteristics of the PORTEC-3 randomized trial used for the analysis of treatment response by HECTOR. *POLE*mut = *POLE* mutant; MMRd = mismatch repair deficient; NSMP = No specific molecular profile; p53abn = p53 abnormal; EBRT = external beam radiotherapy.

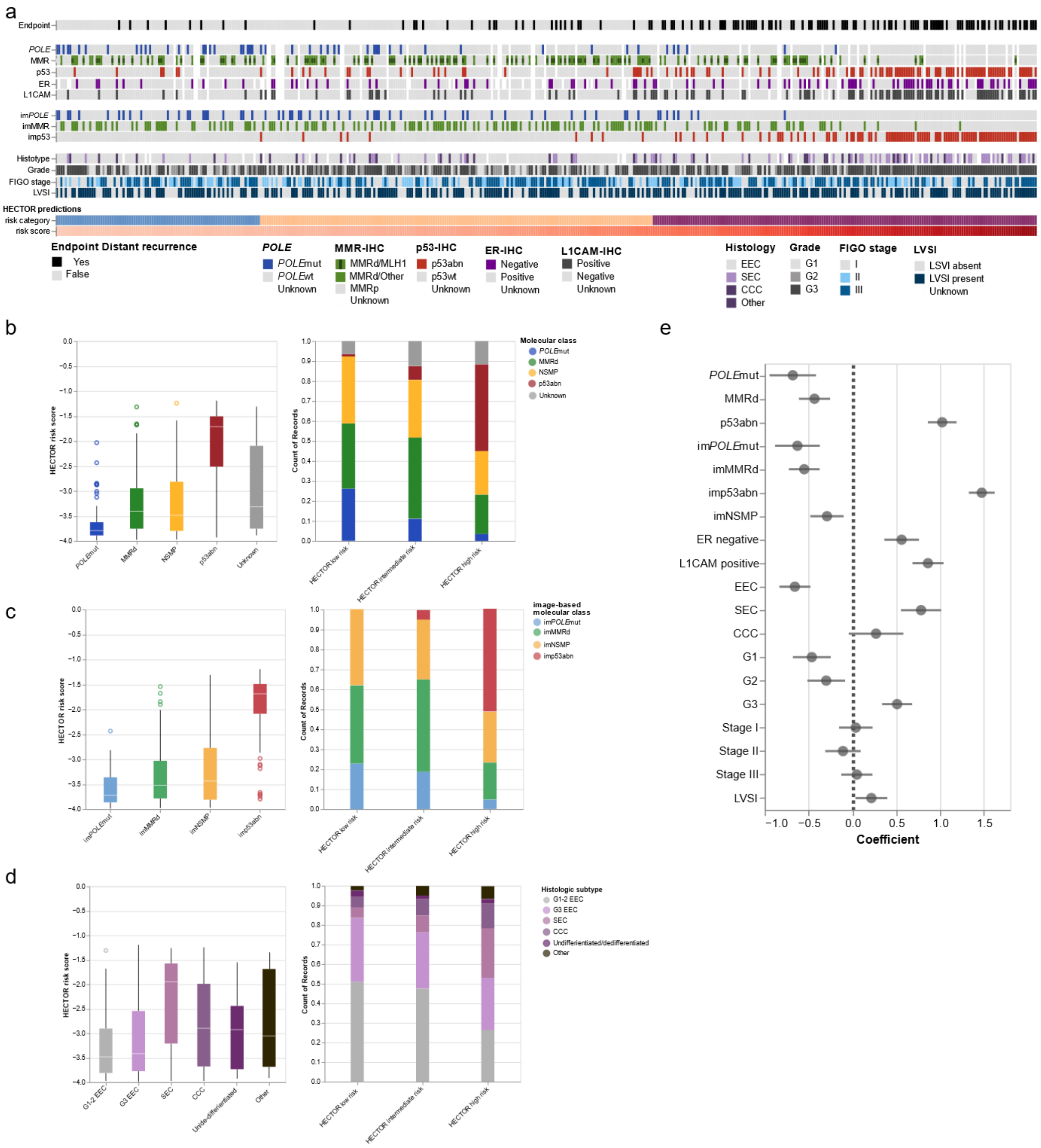

**Figure 26:** Association between HECTOR risk scores and clinicopathological data in the PORTEC-3 randomized trial (*n* = 442). a, Heatmap of prognostic factors for patients ordered by predicted risk scores. b, Association between the molecular class of endometrial cancer and HECTOR risk scores (left) as continuous (right) as categorical using cutoffs of the training set. c, Association between the image-based molecular class as predicted by im4MEC and HECTOR risk scores (left) as continuous (right) as categorical using cutoffs of the training set. d, Association between histologic subtypes and HECTOR risk scores (left) as continuous (right) as categorical using cutoffs of the training set. e, Association of the clinicopathological data and continuous HECTOR risk scores using single linear regression. *POLE*mut = *POLE* mutated; MMRd = mismatch repair deficient; p53abn = p53 abnormal; im = image-based; ER = estrogen receptor; EEC = endometrioid EC; SEC = serous; CCC = clear cell; G1-3 = grade 1-3.

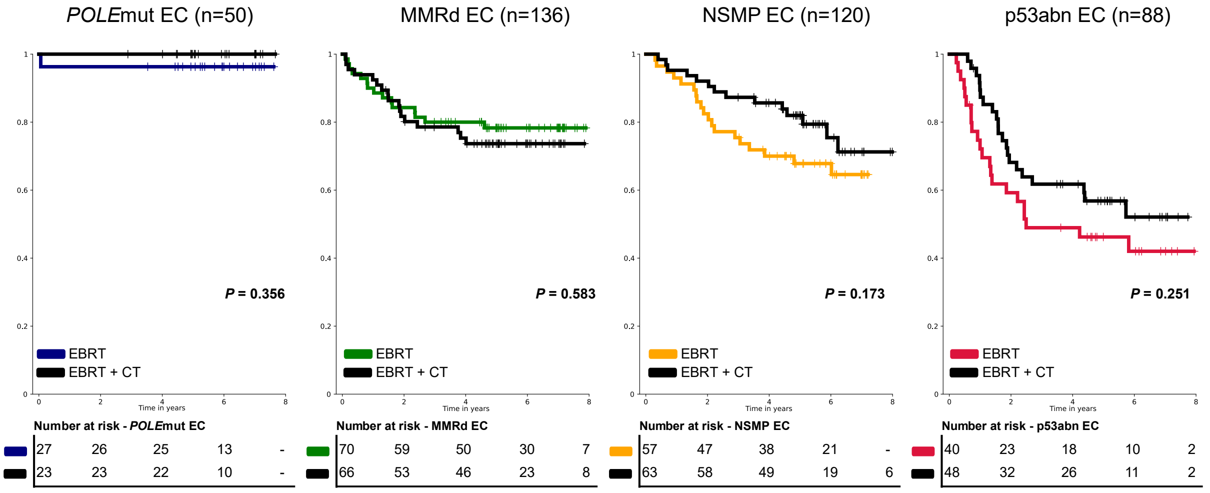


**Figure 27:** Eight-year distant recurrence-free probabilities of molecularly classified patients from the PORTEC-3 randomized trial by molecular class (*POLE*mut EC, MMRd EC, NSMP EC, p53abn EC). PORTEC-3 patients were randomized (1:1) between external beam radiotherapy (EBRT) and external beam radiotherapy and chemotherapy (CT). Log-rank test p-values are reported. *POLE*mut = *POLE* mutant; MMRd = mismatch repair deficient; NSMP = No specific molecular profile; p53abn = p53 abnormal; EC = endometrial cancer.

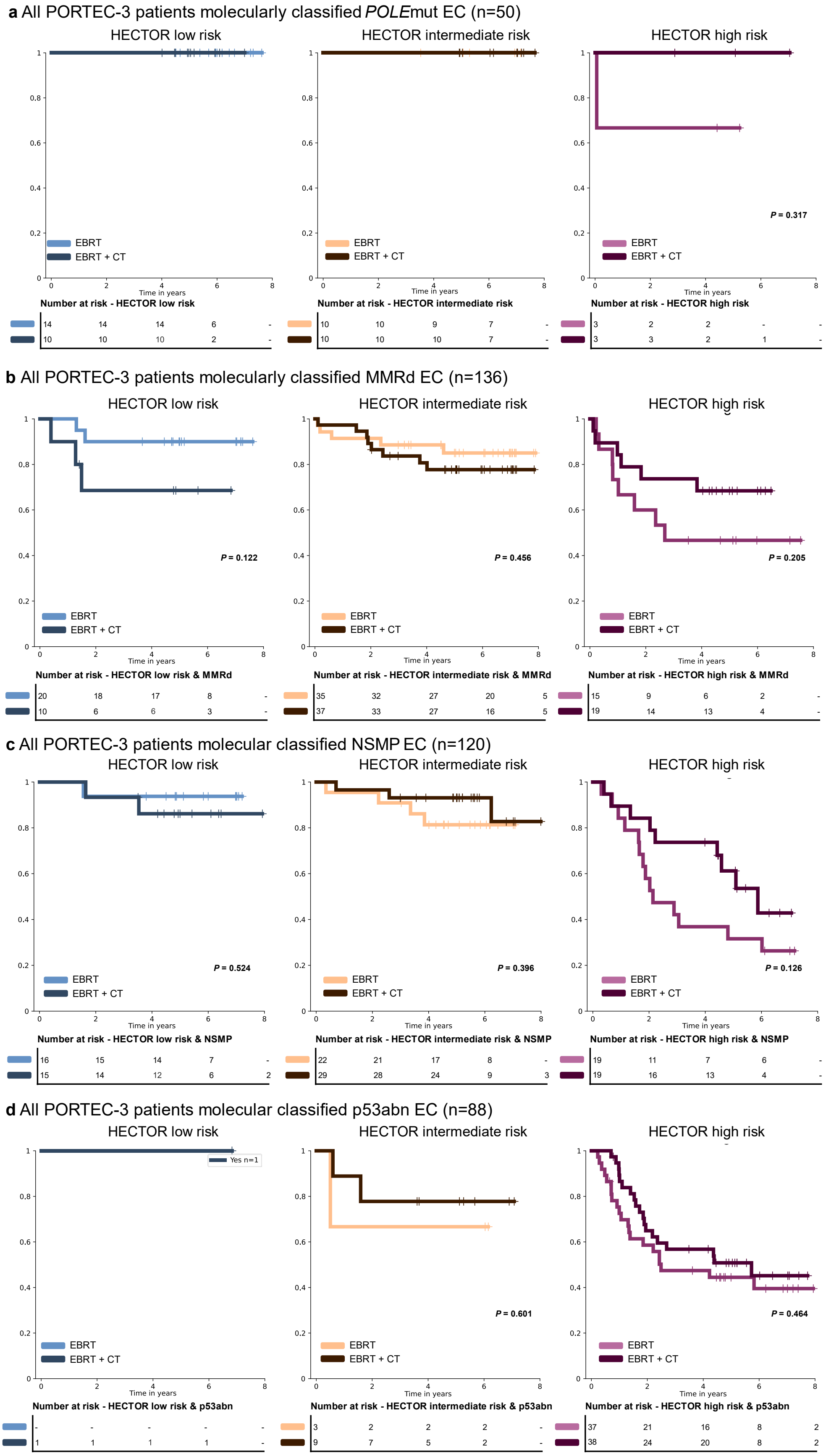


**Figure 28:** Eight-year distant recurrence-free probabilities of patients from the PORTEC-3 randomized trial by HECTOR risk group in the context of the true molecular classification of endometrial cancer

(EC), *POLE*mut, MMRd, NSMP or p53abn EC. PORTEC-3 patients were randomized (1:1) between external beam radiotherapy (EBRT) and external beam radiotherapy and chemotherapy (CT). Only patients with known molecular class were retained (*n* = 394); for reference Figure 29 shows the analysis by molecular class. a, All PORTEC-3 patients molecularly classified as *POLE*mut EC (*n* = 50) and HECTOR low (*n* = 24), intermediate (*n* = 20) and high risk group (*n* = 6), stratified by treatment arm. b, All PORTEC-3 patients molecularly classified as MMRd EC (*n* = 136) and HECTOR low (*n* = 30), intermediate (*n* = 72) and high risk group (*n* = 34), stratified by treatment arm. c, All PORTEC-3 patients molecularly classified as NSMP EC (*n* = 120) and HECTOR low (*n* = 31), intermediate (*n* = 51) and high risk group (*n* = 38), stratified by treatment arm. d, All PORTEC-3 patients molecularly classified as p53abn EC (*n* = 88) and HECTOR low (*n* = 1), intermediate (*n* = 12) and high risk group (*n* = 75), stratified by treatment arm. Log-rank test p-values are reported. *POLE*mut = *POLE* mutant; MMRd = mismatch repair deficient; NSMP = No specific molecular profile; p53abn = p53 abnormal.

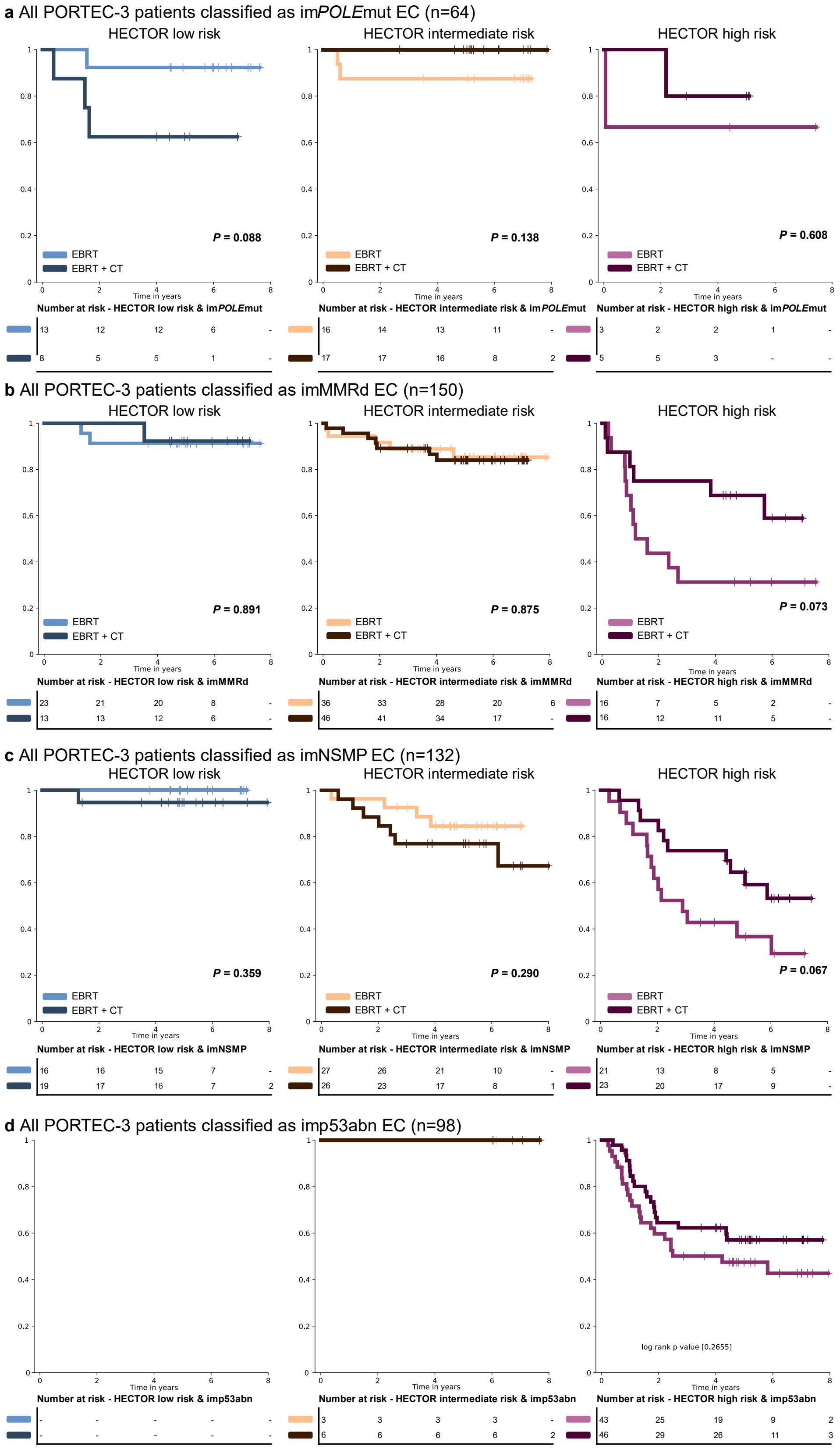


**Figure 29:** Eight-year distant recurrence-free probabilities of patients from the PORTEC-3 randomized trial by HECTOR risk group and stratified by the image-based (im) molecular classification arm of HECTOR, (as predicted by im4MEC[7]). PORTEC-3 patients were randomized (1:1) between external beam radiotherapy (EBRT) and external beam radiotherapy and chemotherapy (CT). a, The PORTEC-3

patients predicted as im*POLE*mut (*n* = 64) and by HECTOR low (*n* = 21), intermediate (*n* = 33) and high risk group (*n* = 8), stratified by treatment arm. b, The PORTEC-3 patients predicted as imMMRd (*n* = 150) and HECTOR low (*n* = 35), intermediate (*n* = 82) and high risk group (*n* = 32), stratified by treatment arm. c, The PORTEC-3 patients predicted as imNSMP (*n* = 132) and HECTOR low (*n* = 35), intermediate (*n* = 53) and high risk group (*n* = 44), stratified by treatment arm. d, The PORTEC-3 patients predicted as imp53abn (*n* = 98) and HECTOR low (*n* = 0), intermediate (*n* = 9) and high risk group (*n* = 89), stratified by treatment arm. Log-rank test p-values are reported. *POLE*mut = *POLE* mutant; MMRd = mismatch repair deficient; NSMP = No specific molecular profile; p53abn = p53 abnormal.
